# Multi-level Regulatory Roles of Lactate Metabolism Gene Network in Oral Cancer: Machine Learning Insights

**DOI:** 10.1101/2025.03.19.25324051

**Authors:** Xiaojing Shen, Zhonghao Liu

## Abstract

This study explores the multi-level regulatory roles of the lactate metabolism gene network in oral cancer development using machine learning models. Machine learning analysis shows that lactate-related gene expression profiles can effectively distinguish treatment groups from controls (AUC=0.933), with SLC16A1 and KIF2C being key features. Genes like PFKP, LDHA, LDHB, LDHC, and VCAN are highlighted for their biological relevance. PFKP, involved in glycolytic metabolic regulation, is significantly upregulated in oral cancer patients, suggesting its role in tumor metabolic adaptation. VCAN, ranking sixth in feature importance, is associated with increased disease risk and may become a therapeutic target. EP300 exhibits a complex relationship with oral cancer, potentially acting as a tumor suppressor. Enrichment analysis links these genes to the HIF-1 signaling pathway and metabolic reprogramming. These findings advance the understanding of lactate metabolism’s role in oral cancer and identify potential therapeutic targets.

## Introduction

Oral cavity and lip cancer is the 16th most common cancer (Bray et al., n.d.), Oral cancer remains a major global health challenge, with rising incidence in both developed and developing countries (Siegel et al., 2019) Due to its late diagnosis, effective treatment is limited and overall five-year survival is below 50 percent due to its poor outcome, delayed onset, aggressive local incendiaries, recurrence and metastasis(Popovici and Ozon, n.d.).The main risk factors included smoking and drinking (Hashibe et al., 2011; Rumgay, n.d.)HPV infection(Tan, 2023; Hübbers and Akgül, n.d.), Among them, squamous cell carcinoma (OSCC) is the most common form, accounting for more than 90% of all oral cancers (Bagan et al., n.d.).The global burden of oral cancer highlights the need for better understanding and new treatments, especially in high-risk populations with chronic habits. Therefore, there is an urgent need to explore new potential biomarkers and features for early diagnosis of ORCA to facilitate targeted and personalized treatment strategies.

Post-translational modifications (PTMs) play a crucial role in regulating protein function and cellular processes, affecting various aspects of disease progression, including cancer. PTMs involve the addition or removal of specific chemical groups from amino acid residues of proteins (Li et al., 2023), T These processes can alter protein activity, localization, stability, and interactions (Czuba et al., 2018). PTMs are critical in tumorigenesis because they regulate key signaling pathways and cellular functions such as proliferation, apoptosis, and metastasis (Sanchez-Vega et al., 2018; Wu et al., 2019; Pan et al., 2020; Senga and Grose, 2021; Fouad and Aanei, n.d.; Hanahan and Weinberg, n.d.), It also plays an important role in regulating the tumor microenvironment, such as remodeling the extracellular matrix (ECM) to treat cancer invasion and chronic inflammation (Leeming et al., 2011; Chang and Ding, 2018; Brown Chandler et al., 2019).In recent years, there has been increasing interest in identifying specific PTMs as therapeutic targets, and many PTMs have been implicated in the pathophysiology of cancer, including oral cancer. Among them, lactylation has emerged as an interesting and novel PTM with increasing relevance to cancer biology.

Lactylation is a newly discovered PTM involving the attachment of lactate molecules to lysine residues of proteins (Zhang et al., 2019). This modification has been shown to affect cellular metabolism (Lv et al., 2023) and gene expression.(Rho et al., 2023). The discovery of lactylation expands our understanding of how metabolic byproducts, such as lactate, affect cellular processes beyond energy production. Lactate, a key metabolite produced during glycolysis, has long been associated with the Warburg effect in cancer cells, increasing glucose metabolism even in the presence of oxygen (Koppenol et al., 2011; Fukushi et al., 2022). Lactate acylation in cancer cells is thought to contribute to tumor progression by promoting adaptations in cellular metabolism and transcriptional regulation, and recent studies have highlighted the involvement of lactate in regulating key oncogenic pathways, making it a potential target for therapeutic intervention (Chen et al., 2024), The role of lactylation in promoting tumor growth and metastasis emphasizes its potential as a therapeutic target for cancer. In oral cancer, lactylation has only recently been explored, but earlier studies suggest that this modification may be a key factor in oral cancer progression. Alterations in cellular metabolism by lactate in oral squamous cell carcinoma (OSCC) may support enhanced proliferation and resistance to apoptosis, as in other cancers. However, the specific mechanisms by which lactylation promotes the pathophysiology of oral cancer remain unclear. The objectives of this study were to investigate the role of lactylation in oral cancer, to explore its potential as a diagnostic or prognostic biomarker, and to evaluate its therapeutic potential. This article aims to review the emerging field of lactylation and its potential application in the treatment of oral cancer, so as to lay the foundation for future research and clinical trials in this field.

Global cancer statistics indicate that oral cancer ranks as the sixteenth most common malignancy, with an increasingly significant disease burden (Bray et al., 2023).

Despite considerable advances in modern oncological therapeutics, the five-year survival rate for oral cancer patients remains below 50%, primarily attributable to late-stage diagnosis, aggressive local tissue invasion, high recurrence rates, and metastatic propensity (Popovici & Ozon, 2022; Siegel et al., 2019).

Histopathologically, over 90% of oral cancer cases are classified as squamous cell carcinoma (OSCC), strongly associated with behavioral risk factors including tobacco consumption and chronic alcohol abuse (Hashibe et al., 2011; Bagan et al., 2021). The evolving epidemiological landscape of this disease has intensified research efforts toward identifying stage-specific biomarkers and molecular-targeted therapeutic approaches to disrupt the pernicious cycle of delayed diagnosis and treatment resistance.

Post-translational modifications (PTMs) exert pivotal regulatory functions in carcinogenesis progression. Through covalent chemical alterations including phosphorylation, ubiquitination, and acetylation, PTMs precisely modulate protein functionality while influencing structural integrity, subcellular localization, and interaction networks (Li et al., 2023; Czuba et al., 2018). These coordinated modifications constitute an intricate regulatory system participating in diverse biological processes ranging from signal transduction to chromatin remodeling. During cancer progression, PTM dysregulation triggers abnormalities in critical cellular pathways, driving malignant transformation (Sanchez-Vega et al., 2018). Notably, PTMs impact not only cell-autonomous behaviors but also alter the tumor microenvironment through extracellular matrix remodeling, facilitating tumor invasion and sustaining chronic inflammatory states (Leeming et al., 2011; Chang & Ding, 2018). Within oral cancer research, therapeutic strategies targeting PTM-specific vulnerabilities are garnering increasing attention. Among these, lactylation has recently emerged as a newly identified PTM subtype whose role in cancer biology is undergoing extensive investigation (Brown Chandler et al., 2019; Wu et al., 2019). Lactylation represents a metabolic-epigenetic interface, involving lactate conjugation to lysine residues on histones and other proteins. This modification serves dual biological functions: modulating chromatin topology to influence transcriptional programs and acting as a real-time sensor of glycolytic activity (Zhang et al., 2019; Rho et al., 2023). By connecting aerobic glycolysis (the Warburg effect) with oncogenic signaling, lactylation may enable tumors to coordinate energy production with malignant phenotype acquisition (Lv et al., 2023; Chen et al., 2024). In OSCC, early evidence suggests lactylation contributes to metabolic optimization and apoptotic resistance, though mechanistic details regarding key regulatory genes and their clinical relevance remain poorly characterized.

Computational analysis of lactylation-associated gene signatures offers a strategic approach to address these gaps. Advanced machine learning algorithms can extract discriminative patterns from multi-omics datasets while minimizing confounding signals, enabling both disease classification and mechanistic exploration. This study applies integrative bioinformatics to oral cancer transcriptomics, focusing on lactylation-related gene expression patterns. The methodology aims to identify prognostic biomarkers and therapeutic targets through systematic evaluation of lactylation’s functional impact, potentially bridging molecular insights with clinical applications in OSCC management.

## Method

### Lactate gene acquisition

The identification of lactylation-related genes for this study was conducted through a systematic integration of literature curation and multi-database mining. Initially, we systematically interrogated three authoritative databases—UniProt, Human Protein Atlas, and PhosphoSitePlus—to identify genes with experimentally validated roles in lactate modification. Priority was given to genes implicated in immune regulation and oncogenesis, including key regulators such as EP300 (histone acetyltransferase), LDHA (glycolytic enzyme), and SIRT1 (deacetylase). This database-driven approach was further augmented by recent clinical findings: Meanwhile, Cheng et al. reported that 16 lactylation-related genes were able to predict the prognosis of liver cancer (Zhengdong et al., 2024). Similarly, Yang et al. found that six lactylation-related genes were significantly associated with gastric cancer prognosis, suggesting that lactylation-related genes are associated with tumor prognosis (Yang et al., 2023).

Through the synthesis of these complementary approaches, we compiled a final set of 45 lactylation-associated genes for subsequent analyses:

AARS1, AARS2, ARID3A, CCNA2, CREBBP, DDX39A, EFNA3, EHMT2, EP300, ESCO2, FABP5, G6PD, H2AX, HBB, HDAC1, HDAC2, HDAC3, HDAC8, HMGA1, KAT5, KAT7, KAT8, KIF2C, LDHA, LDHB, LDHC, MKI67, NUP50, PFKP, PKM2, PLOD2, RACGAP1, RFC4, SIRT1, SIRT2, SIRT3, SIRT6, SLC16A4, SLC16A7, SMARCA4, STC3, STMN1, TKT, VCAN.

These candidate genes were subsequently subjected to machine learning-based predictive modeling and Mendelian randomization analysis to rigorously evaluate their potential causal relationships with disease pathogenesis.

### pQTL data acquisition

pQTL refers to the study of the correlation between genetic variation and gene expression by treating protein expression as a phenotype. We analyzed genetic and phenotypic information on 35,559 Icelanders and preprocessed the downloaded data. Data cleaning and standardization steps included the removal of missing values, batch effect correction, and Z-score normalization to improve data comparability and accuracy of the analysis. Finally, 1,558 preliminary pQTL associations were identified. The selected pQTL loci provided the basis for subsequent Mendelian randomization analyses.

### eQTL data acquisition

Cis-eQTL summary statistics were obtained from the eQTLGen Consortium (https://eqtlgen.org/cis-eqtls.html), specifically the dataset “2019-12-11-cis-eQTLsFDR-ProbeLevel-CohortInfoRemoved-BonferroniAdded.txt.gz”. This dataset includes genome-wide significant cis-eQTL associations (FDR < 0.05) across 37 blood cell types. SNP allele frequency information was merged from the supplementary file “2018-07-18_SNP_AF_for_AlleleB_combined_allele_counts_and_MAF_pos_added.txt.gz” to calculate effect allele frequencies (EAF). We use R packages (TwoSampleMR, dplyr), beta coefficients and standard errors were derived from Z-scores and sample size, with allele frequency adjustments to ensure accurate effect estimates. Retained SNPs with genome-wide significance (P<5×10^-8^ ) for downstream analysis. Performed clumping (clump_kb = 10,000, clump_r2 = 0.001) using the European reference panel to ensure independence of instrumental variables (IVs). Removed SNPs with low instrument strength (F-statistic<20) using R-based calculations to avoid bias in Mendelian randomization.

### Intersection of eQTL, pQTL, and lactatated genes

To show the intersection between pQTL, eQTL, and lactylation genes, Venn diagrams were plotted using the VennDiagram package in the R language. We transformed the above list of pQTL, eQTL, and lactylation genes after data cleaning and screening into vectors in R. The gene ID in each list is unique, ensuring the accuracy of intersection analysis.

### Outcome data

In this study, we used multiple SNP data sets as outcome data. We selected sources from FinnGen Release 11 (C3_ORALCAVITY_EXALLC and CD2_INSITU_ORALC AVITY_EXALLC) and GWAS published on November 4, 2024, Dataset on oral cancer in the Catalog database (GCST012237, GCST012238, GCST012239, ieu-b-4961). The vroom package was used to load the outcome data, and key information such as S NPs, effect sizes (BETA), standard errors (SE), and p-values were extracted. Subsequently, the outcome data were formatted with the format_data function in the TwoSam pleMR package, conforming to the data structure required for MR Analysis.

### Mendelian randomization

In this study, we used the TwoSampleMR package, version 0.6.9, for Mendelian randomization (MR) analyses. Instrumental variables were selected based on SNPs significantly associated with lactylation gene expression in the eQTL data, and a screening criterion with an F-statistic greater than 10 was used to avoid weak instrumental variable bias. Data processing and reconciliation were done with the harmonise_data function, ensuring consistency of effect alleles and effect sizes. The primary analysis was performed by inverse variance weighting (IVW), and sensitivity analysis was performed by combining weighted median and MR-Egger regression. Heterogeneity was assessed by Cochran’s Q statistic, and horizontal pleiotropy was tested by MR-Egger intercept. The final results were expressed by the odds ratio (OR) and its 95% confidence interval (CI). All analyses were done in the R language environment using packages such as TwoSampleMR, ieugwasr, MRInstruments, and dplyr. Analyses followed the 20-item checklist of STROBE-MR (Reporting standards for Observational Studies with Mendelian Randomization) to ensure scientific validity and reproducibility of the study.

### GEO Data acquisition and pre-processing

In this study, we used the keywords “oral cancer” to search the GEO database to identify qualified datasets. We set specific search criteria: Item Type was set to Series, Study type was set to Analysis by Array Expression, and species were limited to humans. After careful screening, we finally selected GSE13601, GSE23558, and GSE30784 datasets and downloaded the corresponding platform files. For each dataset, we used R scripts to transform gene probe IDs into gene symbols, which were subsequently normalized. In addition, we used vroom and data.table packages to efficiently read and integrate multiple GEO datasets, which ensured the consistency of gene expression data between different datasets. To remove batch effects, we used the ComBat method in the sva package, which effectively corrects for batch effects from different datasets so that gene expression data between datasets are compared on the same scale. The final batch effect corrected data were exported and saved as standardized data files for subsequent differential expression analysis.

### Differential expression analysis

The Wilcoxon rank-sum test was performed using the wilcox.test() function, and significant genes were screened according to the criterion of p-value < 0.05. To control false positive results due to multiple testing, p-values were corrected by the Benjamini-Hochberg (BH) adjustment method, which ensured that the false discovery rate (FDR) was controlled within a reasonable range. Boxplot was used for visualization to visually show the expression difference of each target gene between the control and experimental groups.

### Machine Learning predict potential targets

The lactate-modified gene expression analysis framework employs a three-stage analytical design to achieve comprehensive interpretation from raw data to biological insights. The workflow sequentially performs: 1) classification modeling of lactylation-related genes, 2) dual-validation feature importance assessment, and 3) SHAP-based interaction analysis, establishing a complete analytical pipeline from data processing to mechanistic interpretation.

For classification model construction, XGBoost algorithm was selected for its demonstrated superiority in handling high-dimensional biological data. To enhance model robustness, we implemented cross-validation strategy for parameter optimization and objectively evaluated model performance using ROC curve analysis with AUC quantification. Notably, the input features were strictly confined to lactylation-related gene sets, thereby effectively minimizing interference from non-specific signals.

Feature contribution assessment adopted a dual-validation approach: The XGBoost’s inherent feature importance algorithm quantified gene contributions during decision tree construction, while SHAP analysis provided game theory-based quantification of directional impacts and magnitude effects on model outputs. Additionally, Principal Component Analysis (PCA) was applied to visualize sample distribution patterns in lactylation-related gene expression space, systematically evaluating the collective discriminative capacity of these genes across different physiological states.

## Result

### Intersection features of lactatated genes in eQTL and pQTL

Our comprehensive analysis revealed limited convergence between lactylation-associated genes and quantitative trait loci (QTL) genes. Specifically, only 29 genes (0.2% of total analyzed genes) demonstrated dual associations as both expression QTL (eQTL) and lactylation-related genes (Figure 1A). Similarly, protein QTL (pQTL) genes showed minimal overlap with lactylation targets, with merely four genes (0.3% of total pQTL genes) exhibiting concurrent associations (Figure 2B).

**Figure 1.**
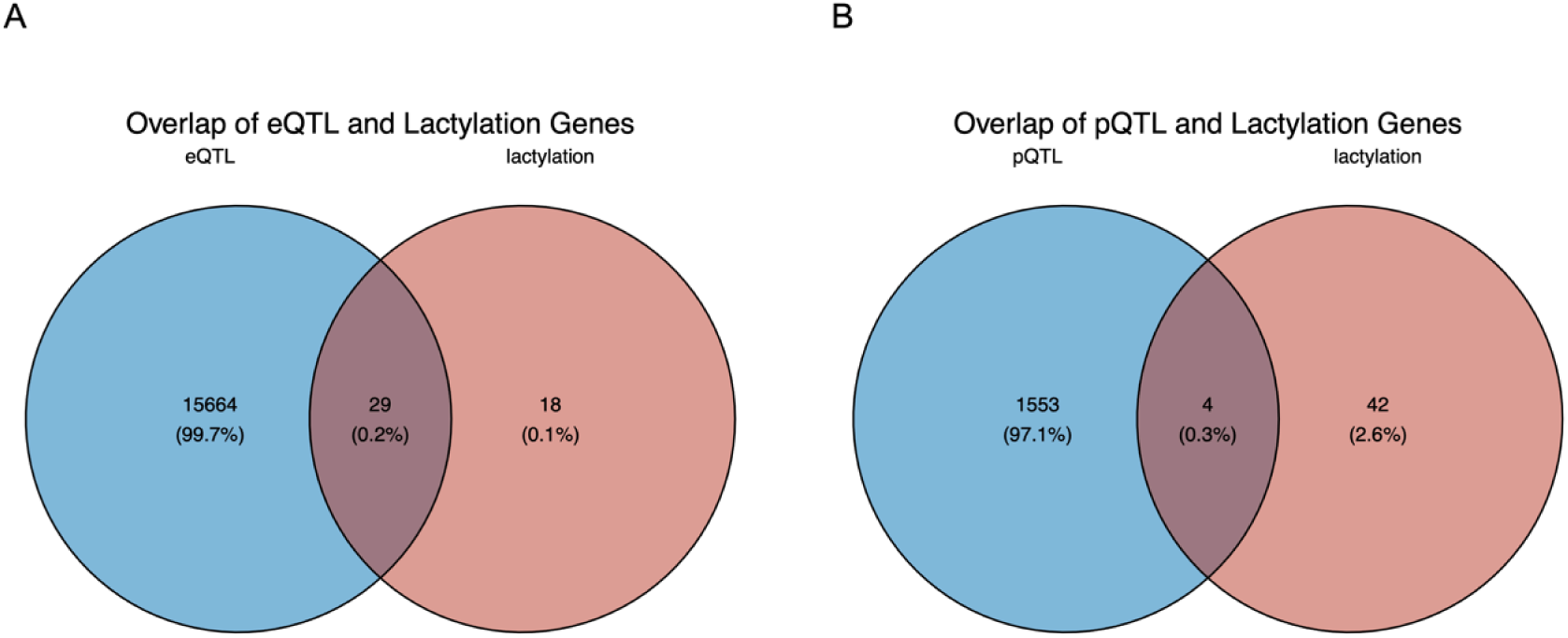
**Intersection of eQTL and pQTL genes with lactatation genes**

**Figure 2.**
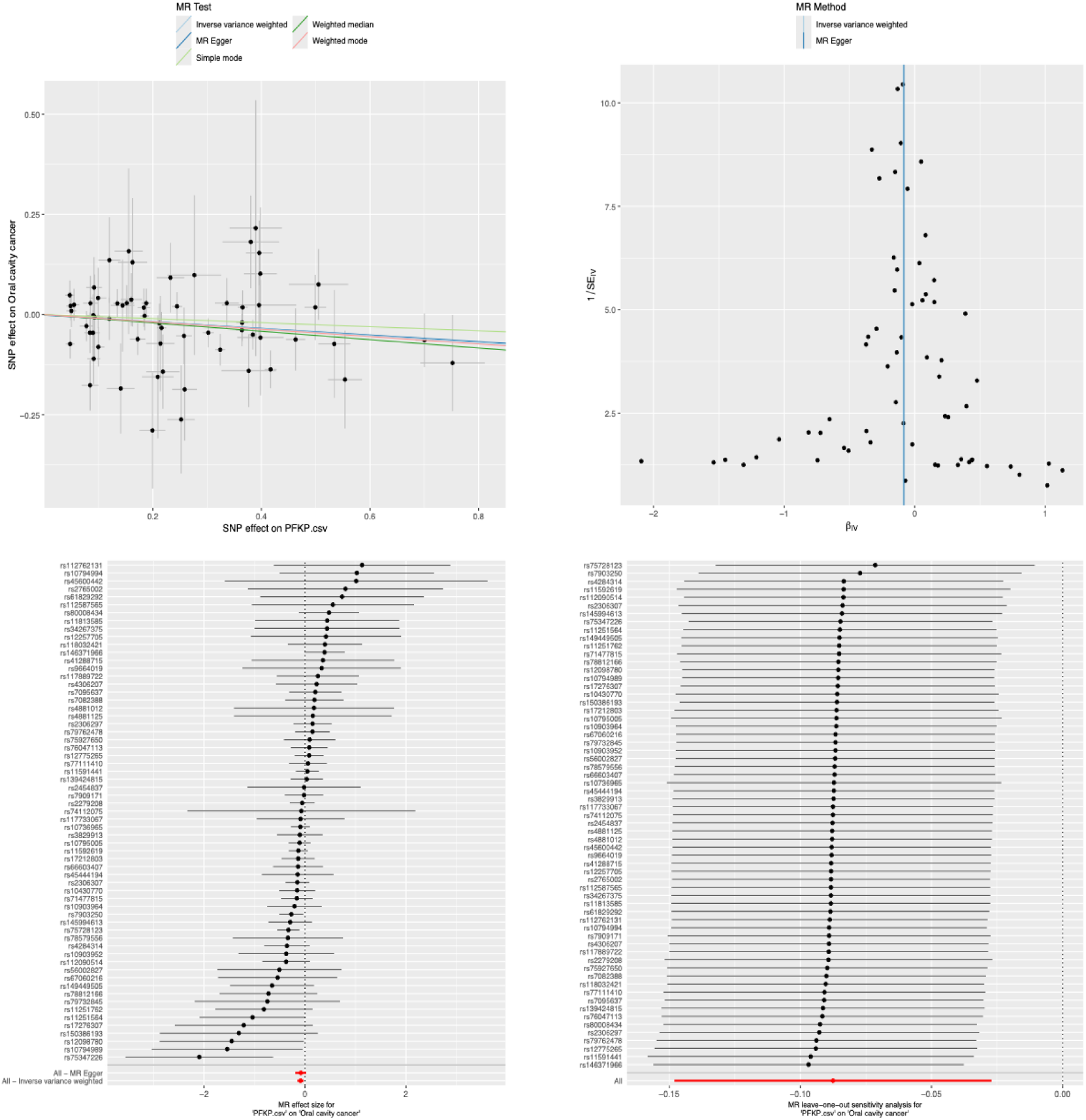
**SNP effect analysis and MR Method validation plots in genomic association studies**

This striking disparity in gene set overlap suggests distinct functional specialization, where eQTL/pQTL genes and lactylation-modified genes likely participate in divergent biological pathways and regulatory mechanisms. Notably, the observed partial convergence, though statistically modest, implies potential crosstalk between lactylation modifications and QTL-mediated regulation. These findings position protein lactylation as a potentially unique regulatory layer that may complement traditional QTL mechanisms in governing gene expression and protein function.

### Analysis of lactate gene expression and eqtlMR in oral cancer

This study employed a bidirectional Mendelian randomization (MR) approach to investigate causal relationships between lactylation-associated genes and oral carcinogenesis, leveraging integrated data from the FinnGen R12 biobank and GWAS Catalog. Genetic instruments derived from expression quantitative trait loci (eQTLs) were analyzed through five complementary MR techniques: MR-Egger regression, weighted median, inverse variance weighting (IVW), and simple/weighted mode analyses, ensuring methodological robustness.

Primary analysis of the FinnGen_R12_C3_ORALCAVITY_EXALLC cohort identified PFKP as a protective locusand VCAN as a risk-associated gene. Replication in the FinnGen_R12_CD2_INSITU_ORALCAVITY_EXALLC dataset confirmed these associations while revealing additional protective effects for EP300 (Supplementary Table S1).

Cross-validation across four GWAS Catalog cohorts reinforced the pathogenic roles of PEKP, VCAN, and EP300. Metabolic regulators LDHA, LDHC, SIRT1, and chromatin remodeler SMARCA4 exhibited consistent pro-carcinogenic effects, whereas SIRT3 and TKT demonstrated tumor-suppressive properties (all IVW p < 0.05). Proteome-wide MR (pQTL) analyses suggested a marginal association between LDHB and oral cancer, though statistical significance thresholds were not met.

### Differential expression analysis of genes involved in lactatation

After batch effect correction by applying the ComBat method (bottom left panel), the gene expression levels of all items tended to be consistent and the distribution between data points more coincided. This indicates that batch effects have been successfully removed and gene expression data from different projects have become more uniform. We further evaluated the effectiveness of batch effect adjustment by principal component analysis (PCA). As shown in (upper right panel), before batch effect correction, the samples of the three sets of data show clear separation on the first principal component (PC1), GSE13601 (blue) and GSE23558 (red) form different clusters, and GSE30784 (gray) is independently distributed on the second principal component (PC2). This separation reflects technical variation due to batch effects. After correction for batch effects (lower right panel), the samples of the three sets of data are more mixed and the degree of cluster overlap between items is greater. This indicates that batch effects have been effectively removed and that comparability between datasets has been significantly improved.

These results suggest that batch effect correction using the ComBat method significantly reduces technical variation between datasets, thereby improving the reliability of subsequent analyses.

To evaluate the expression differences of candidate lactatation-related genes between oral cancer patients and normal individuals, we performed differential expression analysis of 10 target genes using GEO data. Analysis was performed using the Wilcoxon rank sum test to compare differences in gene expression levels between the normal group (Control) and the cancer group (Treat). The results of the analysis are summarized as follows: LDHA, LDHB, LDHC, PFKP and VCAN genes were significantly up-regulated in patients with oral cancer, especially LDHA, LDHC and VCAN genes (p values were 3.65 ×10^-16, 6.95 ×10^-6 and 1.75 ×10^-15, respectively). In addition, EP300 was significantly down-regulated in oral cancer patients (p = 0.0274). However, the expression differences of TKT, SIRT1 and SIRT3 genes between the two groups did not reach statistical significance. To show the differences in the expression of these genes more intuitively, we plotted boxplots , where blue represents normal individuals and orange represents oral cancer patients. The figure shows that the expression levels of LDHA, LDHB, LDHC, PFKP, and VCAN genes were significantly higher in oral cancer patients than in normal individuals, while the EP300 gene showed the opposite pattern. These results suggest that lactatation-related genes may play an important role in the development and progression of oral cancer, and provide important clues for further research on the molecular mechanisms and potential therapeutic targets of oral cancer.

**Figure 3.**
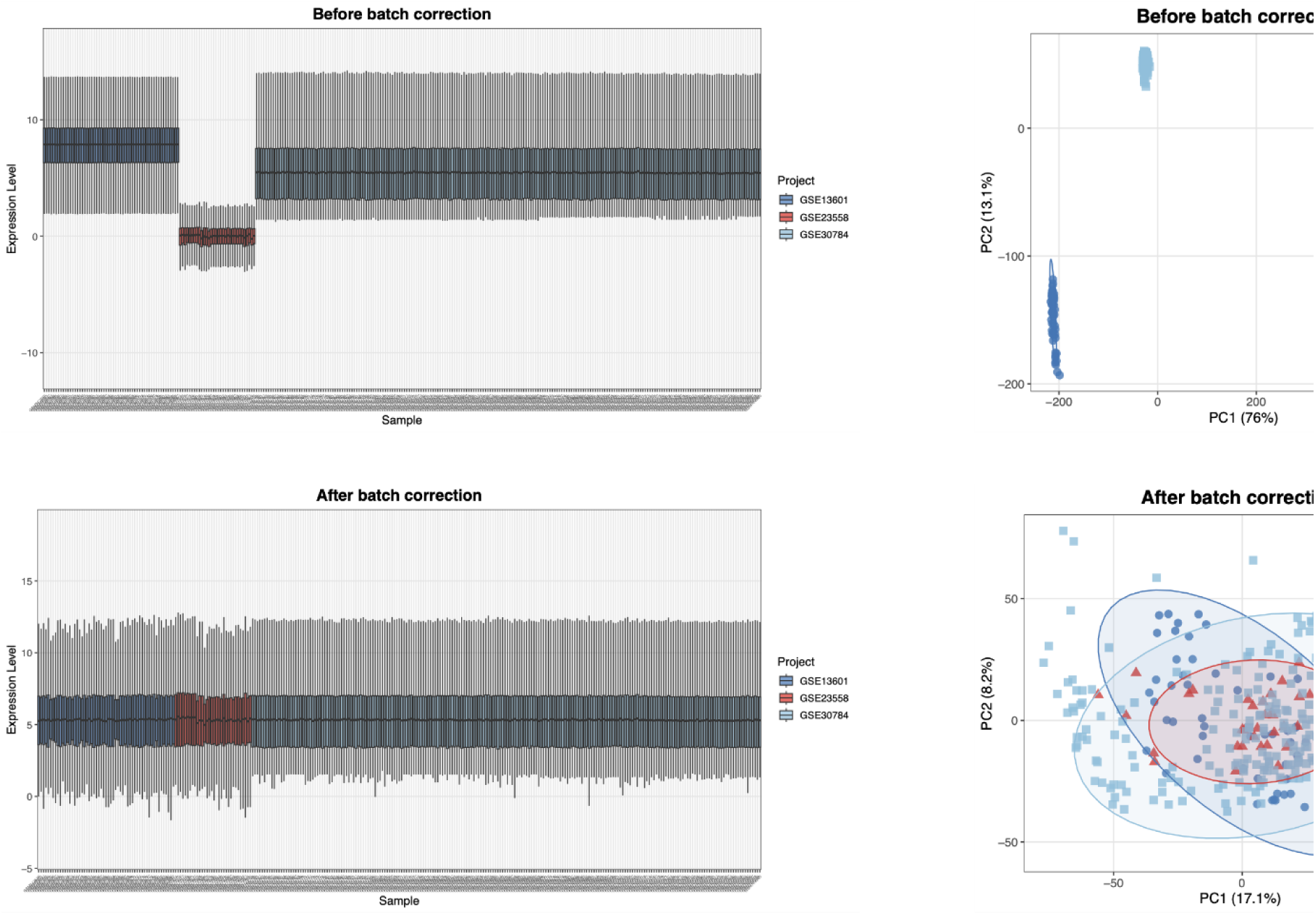
**GEO data correction comparison**

### Protein-Protein Interaction (PPI) and Pathway Analysis

In this study, we conducted protein-protein interaction (PPI) network analysis and pathway enrichment analysis on intersecting genes. The PPI network revealed that these genes form multiple densely interconnected modules within cells, with several hub genes exhibiting higher degrees of connectivity, suggesting their potential roles as key regulators. These hub genes were primarily associated with lactate transmembrane transporters and various dehydrogenase activities. Notably, the monocarboxylate transporter (MCT) family plays a critical role in lactate transmembrane transport, and its interaction with lactate dehydrogenase (LDH) may regulate the metabolic directionality of lactate.

DAVID pathway enrichment analysis further demonstrated that KEGG-enriched genes were predominantly clustered in the HIF-1 signaling pathway, as well as glycolysis/gluconeogenesis pathways - biological processes closely associated with carcinogenesis. The HIF-1 signaling pathway mediates metabolic reprogramming under hypoxic conditions by promoting glycolysis and lactate production, thereby influencing tumor progression.

Gene Ontology (GO) analysis revealed significant associations of these genes with energy metabolism processes, including pyruvate catabolic processes and transmembrane transport. At the molecular functional level, these genes showed predominant associations with deacetylase activities targeting specific histone residues: H3K18, H3K56, H4K16, H3K14, H3K9, and H3K4. These findings elucidate the potential roles of intersecting genes in cellular physiological functions and provide valuable insights for subsequent investigations into their mechanistic contributions.

**Figure 4.**
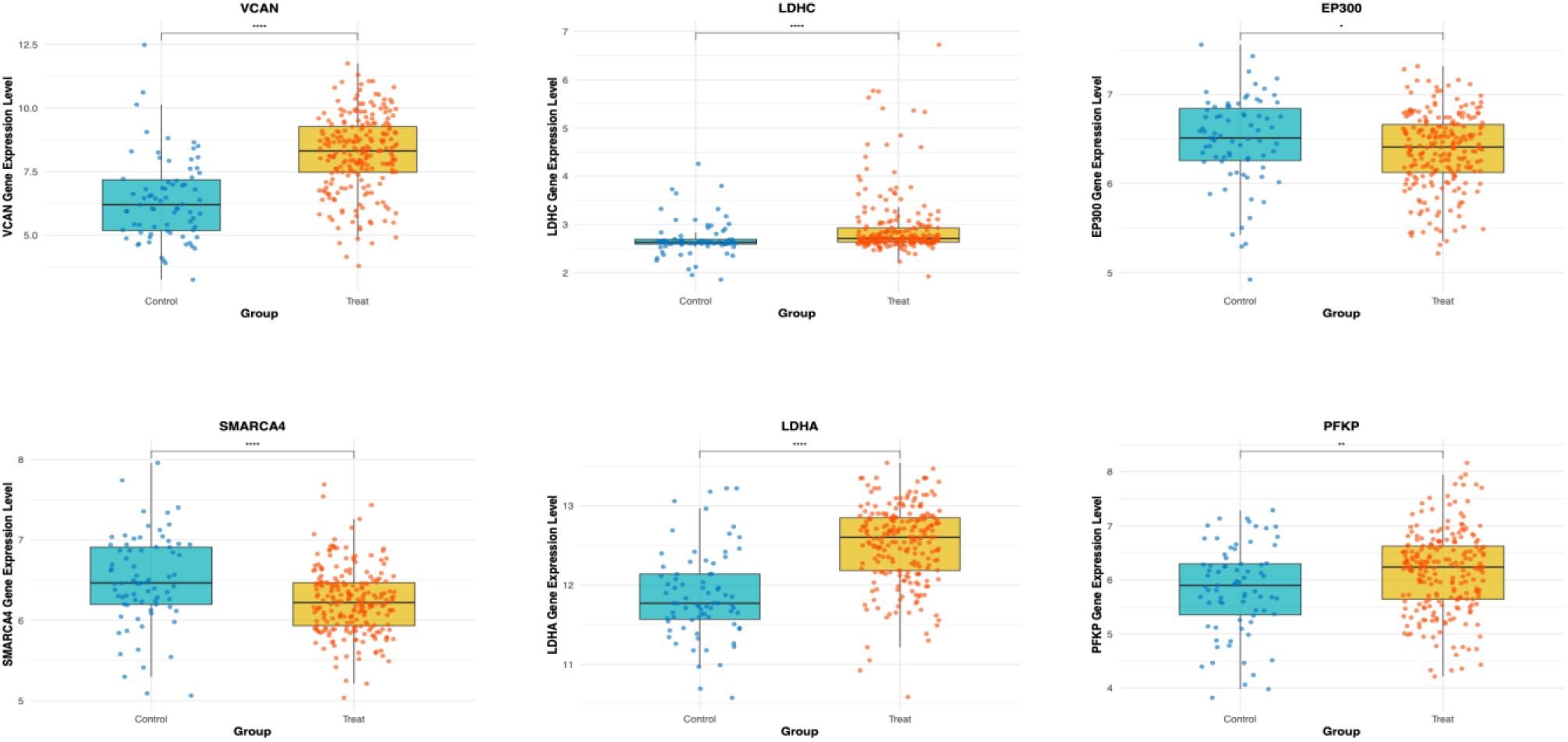
**Box plot of core gene expression**

### Machine Learning-Based Feature Selection and Predictive Analysis

The lactate-related gene expression profile effectively distinguished the treatment group from the control group. The model’s AUC of 0.933 from ROC curve analysis confirms that these gene expression patterns contain key biological information. The model achieved a high true positive rate with a low false positive rate, indicating that lactate gene-based classification is highly specific and sensitive. Gene contribution analysis revealed a hierarchy of key lactate genes. SLC16A1 and KIF2C were ranked highest by XGBoost’s feature importance, with scores of 0.24 and 0.23 respectively. SHAP values confirmed SLC16A1’s central role and highlighted FABP5, RFC4, and PLOD2 as important contributors to the classification.

SHAP analysis showed complex associations between gene expression and phenotypes. High SLC16A1 expression correlated with the treatment group (positive SHAP value), while SLC16A7 and HBB showed the opposite pattern, being associated with the control group. This differential regulation may reflect the lactate pathway’s complex regulation in disease states. PCA showed that lactate gene expression data could partially distinguish the two groups in two-dimensional space. The first and second principal components explained 23.44% and 10.56% of the variance respectively. Treatment samples mostly occupied the positive PC1 region, while control samples concentrated in the negative PC1 region. Despite some overlap, the overall distribution aligns with the classification model’s high performance.

Through multidimensional analysis, SLC16A1, KIF2C, FABP5, and RFC4 were identified as core markers for distinguishing disease states. These gene expression differences provide new insights into lactate’s potential role in disease pathogenesis and lay the foundation for biomarker screening and molecular mechanism analysis.

**Figure 5.**
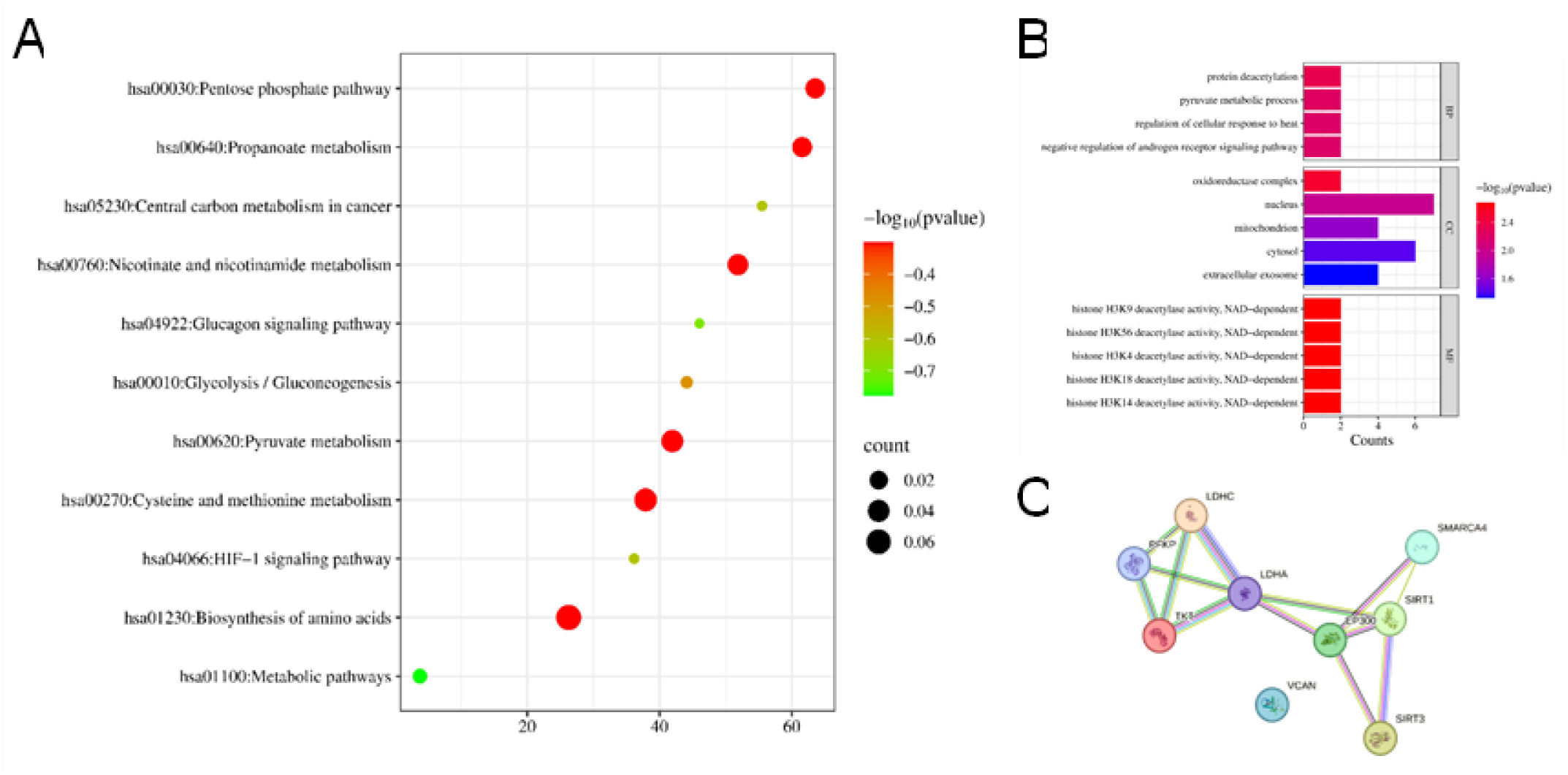
KEGG and GO Annotation

**Figure 6.**
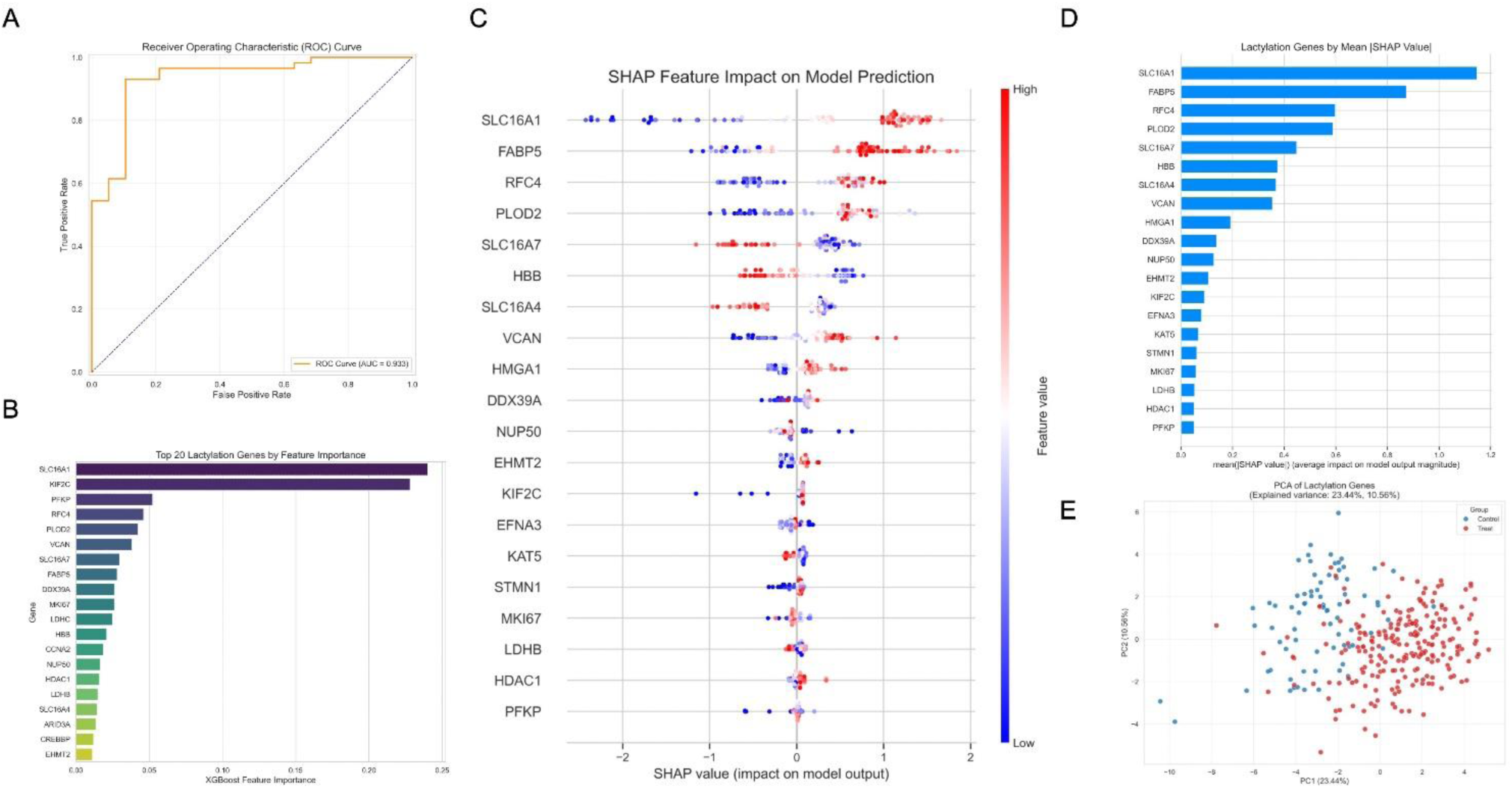
Machine learning-based feature importance analysis of lactylation-related genes in oral cancer. (A) ROC curve of the predictive model, showing an AUC of 0.933. (B) Top 20 lactylation-related genes ranked by feature importance in the XGBoost model. (C) SHAP summary plot indicating the impact of each gene on model predictions. (D) Mean absolute SHAP values of lactylation genes, highlighting their contribution to the model. (E) Principal component analysis (PCA) of lactylation-related genes in oral cancer and control samples, illustrating the separation between the two groups.

## Discussion

Oral cancer is a serious malignancy that poses a significant threat to patient survival and quality of life. Despite several advances in diagnosis and treatment in recent years, current treatments still have limitations and do not fully meet clinical needs. Therefore, the discovery of new therapeutic targets and the development of more effective therapies are essential to improve the prognosis of patients with oral cancer.

This study systematically elucidated the multi-level regulatory roles of the lactate metabolism gene network in the occurrence and development of oral cancer by integrating machine learning models with Mendelian randomization analysis.

Machine learning analysis showed that the expression profiles of lactate-related genes could effectively distinguish the treatment group from the control group (AUC=0.933), with transport-related genes such as SLC16A1 (MCT1) and KIF2C dominating in feature importance analysis. Notably, EP300, TKT, SIRT1, LDHA, and SIRT3, which were significantly associated with oral cancer risk in the MR analysis, did not rank in the top 20 in the ML model’s feature ranking, while PFKP, LDHB, LDHC, and VCAN showed biological relevance in both analysis methods.

In recent years, it has been found that PFKP (phosphofructokinase-1) plays an important role in a variety of cancers, especially in tumor metabolism and progression. For example, in lung adenocarcinoma (LUAD) and ovarian cancer (OVCA), high expression of PFKP is significantly associated with tumor invasion and metastasis(Faubert et al., 2020). The expression level of PFKP is positively correlated with tumor mutational burden (TMB) and microsatellite instability (MSI), suggesting that it may affect the immune microenvironment of tumors(Ling et al., 2024).

Our MR analysis revealed a significant inverse association between PFKP expression and oral cancer risk, suggesting its potential protective role in carcinogenesis. In machine learning frameworks, PFKP emerged as a key discriminative feature, securing a prominent position in the XGBoost feature ranking (importance score: 0.052). However, SHAP value analysis revealed a nuanced pattern - while PFKP contributes to model differentiation, its individual predictive impact remains moderate compared to top-tier features like SLC16A1.As a lactatation-related gene, PFKP participates in metabolic regulation in the glycolytic pathway. Its negative correlation may reflect that PFKP plays a protective role in oral carcinogenesis by regulating the metabolic state of cells and reducing the energy supply of tumor cells. However, the results of differential expression analysis showed that the expression of PFKP gene was significantly upregulated in oral cancer patients, suggesting that PFKP may be involved in metabolic reprogramming in tumor cells.

Lactate modification may promote the high expression of PFKP gene in the tumor microenvironment to support the rapid proliferation of tumor cells. Cancer cells typically undergo metabolic reprogramming and rely on enhanced glycolytic pathways for energy supply, which may be one reason for PFKP upregulation(Faubert et al., 2020). Thus, although MR Analysis suggests a protective role for PFKP, the results of differential expression analysis suggest that it functions as part of metabolic adaptation in tumor cells.

In addition to PFKP, the upregulation of genes such as LDHA, LDHC, and VCAN also suggested an important role of lactatation in tumor metabolism. In particular, LDHA and LDHC, as key enzymes in lactate metabolism, may play a key role in the metabolic adaptation of tumor cells by their lactatation modification. By lactatation modification, cancer cells are able to regulate their metabolic pathways to meet the energy demands required for rapid proliferation.

VCAN is highly expressed in various types of cancer, such as gastric cancer, hepatocellular carcinoma, and oral squamous cell carcinoma, and its high expression is associated with poor prognosis in patients. It mediates the movement of tumor cells by forming large molecular complexes with CD44 and hyaluronic acid, thereby promoting the invasion and metastasis of tumor cells.In the machine learning analysis, VCAN ranks sixth in the XGBoost feature importance. The SHAP plot shows that the expression distribution of VCAN is mainly concentrated in the positive SHAP value area and appears in red, indicating that high expression of VCAN is associated with an increased disease risk predicted by the model. This result is consistent with our Mendelian randomization (MR) analysis results, jointly supporting the possible role of VCAN in promoting the development of oral cancer. Given the multi-level regulatory role of VCAN in oral cancer, it may become a potential therapeutic target. By targeting VCAN, it is possible to disrupt the composition and function of the tumor microenvironment, thereby inhibiting tumor invasion and metastasis.

Meanwhile, combined with the prediction results of machine learning models, it can more accurately screen out patient groups that may benefit from VCAN-targeted therapy, improving treatment outcomes and reducing side effects.

At the same time, the down-regulation of EP300 gene may be related to the regulation of lactatation modification, suggesting that EP300 may inhibit tumorigenesis by regulating lactatation and other modifications.

The expression and mutation of EP300 are closely related to the occurrence and development of tumors. However, its relationship with tumors is complex, and it has dual effects, that is, it has tumor suppression or tumor promotion in different tumor types or different stages of tumors(Mees et al., 2010; Asaduzzaman et al., 2018; Zhu et al., 2020; Kim et al., 2022). In some cases, EP300 regulates gene expression through lactatation modification and may promote tumor cell proliferation and migration. For example, in gastric cancer, EP300 activates the YAP-TEAD complex through lactatation modification, thereby promoting the malignant proliferation of tumor cells(Ju et al., 2024). However, in other studies, the lactatation modification of EP300 may play the opposite role. For example, in melanoma, the binding level of EP300 in the YTHDF2 promoter region is positively correlated with the level of histone lactate, and its activity is regulated by glycolytic inhibitors(Yu et al., 2021).

In the present study, we found a significant inverse association between the EP300 gene and oral cancer risk and a significant downregulation of the EP300 gene in patients with oral cancer. This suggests that EP300 may play a tumor suppressor role in the development of oral cancer. However, we remain skeptical about the results showing that high EP300 expression in oral squamous cell carcinoma (OSCC) significantly promotes tumor cell proliferation and migration. We speculate that, on the one hand, downregulation of EP300 may inhibit tumor formation and progression at an early stage, which is consistent with the results of our MR Analysis. On the other hand, EP300 may be hijacked by tumor cells in some cases to promote the malignant progression of tumors by enhancing cell proliferation and migration signaling pathways.

Our study focused on VCAN, LDHC, LDHA, PFKP, TKT, EP300, SIRT1, SIRT3, SMARCA4 and other genes and performed enrichment analysis. The results showed that these genes were changed to different degrees in oral cancer and were closely related to lactatation modification. In our study, these changes in lactication genes were closely related to the HIF-1 signaling pathway, which is activated under hypoxic conditions and regulates the expression of glycolysis-related genes, thereby promoting metabolic reprogramming of tumors. In addition, SIRT1 and SIRT3, as “eraser” of lactate, play important roles in glycolysis and cell metabolism, further affecting the progression of oral cancer(Du et al., 2024). Through enrichment analysis, we found that the changes in these genes were not only involved in glycolytic and gluconeogenic pathways, but also closely related to the activation of HIF-1 signaling pathway, which further demonstrated the reliability and importance of our findings.

Future research should further investigate the specific molecular mechanisms and validate the clinical applicability of these findings to advance the development of effective therapies for oral cancer.

## Conclusion

This study systematically revealed the multidimensional regulatory mechanisms of lactate metabolism-related genes in the occurrence and development of oral cancer by integrating machine learning models with Mendelian randomization (MR) analysis.The findings revealed that lactate-related genes, such as PFKP, VCAN, LDHA, LDHC, and EP300, play significant roles in the development and progression of oral cancer. PFKP exhibited a potential protective role in carcinogenesis despite being upregulated in patients, possibly due to its involvement in metabolic adaptation within tumor cells. VCAN was identified as a potential therapeutic target that could disrupt the tumor microenvironment and inhibit cancer progression. EP300 demonstrated a complex dual role, acting as a tumor suppressor in some contexts while potentially being exploited by tumor cells to promote malignancy in others.

## Data Availability

All data produced are available online at GWAS catalog (https://www.ebi.ac.uk/gwas/) and Gene Expression Omnibus (https://www.ncbi.nlm.nih.gov/geo/)

## Reference

Asaduzzaman, M., Constantinou, S., Min, H., Gallon, J., Lin, M.-L., Singh, P., et al. (2018). Correction to: Tumour suppressor EP300, a modulator of paclitaxel resistance and stemness, is downregulated in metaplastic breast cancer. Breast Cancer Res Treat 167, 605–606. doi: 10.1007/s10549-017-4633-6

Bagan, J., Sarrion, G., and Jimenez, Y. (n.d.). Oral cancer: Clinical features.

Bray, F., Laversanne, M., Sung, H., Me, J. F., Siegel, R. L., Soerjomataram, I., et al. (n.d.). Global cancer statistics 2022: GLOBOCAN estimates of incidence and mortality worldwide for 36 cancers in 185 countries.

Brown Chandler, K., E. Costello, C., and Rahimi, N. (2019). Glycosylation in the Tumor Microenvironment: Implications for Tumor Angiogenesis and Metastasis. Cells 8, 544. doi: 10.3390/cells8060544

Chang, S. C., and Ding, J. L. (2018). Ubiquitination and SUMOylation in the chronic inflammatory tumor microenvironment. Biochimica et Biophysica Acta (BBA) - Reviews on Cancer 1870, 165–175. doi: 10.1016/j.bbcan.2018.08.002

Chen, H., Li, Y., Li, H., Chen, X., Fu, H., Mao, D., et al. (2024). NBS1 lactylation is required for efficient DNA repair and chemotherapy resistance. Nature 631, 663–669. doi: 10.1038/s41586-024-07620-9

Czuba, L. C., Hillgren, K. M., and Swaan, P. W. (2018). Post-translational modifications of transporters. Pharmacology & Therapeutics 192, 88–99. doi: 10.1016/j.pharmthera.2018.06.013

Du, R., Gao, Y., Yan, C., Ren, X., Qi, S., Liu, G., et al. (2024). Sirtuin 1/sirtuin 3 are robust lysine delactylases and sirtuin 1-mediated delactylation regulates glycolysis. iScience 27, 110911. doi: 10.1016/j.isci.2024.110911

Faubert, B., Solmonson, A., and DeBerardinis, R. J. (2020). Metabolic reprogramming and cancer progression. Science 368, eaaw5473. doi: 10.1126/science.aaw5473

Fouad, Y. A., and Aanei, C. (n.d.). Revisiting the hallmarks of cancer.

Fukushi, A., Kim, H.-D., Chang, Y.-C., and Kim, C.-H. (2022). Revisited Metabolic Control and Reprogramming Cancers by Means of the Warburg Effect in Tumor Cells. IJMS 23, 10037. doi: 10.3390/ijms231710037

Hanahan, D., and Weinberg, R. A. (n.d.). The Hallmarks of Cancer Review.

Hashibe, M., Brennan, P., Chuang, S., Boccia, S., Castellsague, X., Chen, C., et al. (2011). Interaction between tobacco and alcohol use and the risk of head and neck cancer: pooled analysis in the INHANCE consortium.

Hübbers, C. U., and Akgül, B. (n.d.). HPV and cancer of the oral cavity.

Ju, J., Zhang, H., Lin, M., Yan, Z., An, L., Cao, Z., et al. (2024). The alanyl-tRNA synthetase AARS1 moonlights as a lactyltransferase to promote YAP signaling in gastric cancer. Journal of Clinical Investigation 134, e174587. doi: 10.1172/JCI174587

Kim, K.-B., Kabra, A., Kim, D.-W., Xue, Y., Huang, Y., Hou, P.-C., et al. (2022). KIX domain determines a selective tumor-promoting role for EP300 and its vulnerability in small cell lung cancer. Sci. Adv. 8, eabl4618. doi: 10.1126/sciadv.abl4618

Koppenol, W. H., Bounds, P. L., and Dang, C. V. (2011). Otto Warburg’s contributions to current concepts of cancer metabolism. Nat Rev Cancer 11, 325–337. doi: 10.1038/nrc3038

Leeming, D. J., Bay-Jensen, A. C., Vassiliadis, E., Larsen, M. R., Henriksen, K., and Karsdal, M. A. (2011). Post-translational modifications of the extracellular matrix are key events in cancer progression: Opportunities for biochemical marker development. Biomarkers 16, 193–205. doi: 10.3109/1354750X.2011.557440

Li, Y., Li, B., Yang, K., Zhu, L., Tang, H., Huang, Y., et al. (2025). PER3 suppresses tumor metastasis of oral squamous cell carcinoma by promoting HIF-1α degradation. Translational Oncology 52, 102258. doi: 10.1016/j.tranon.2024.102258

Li, Y., Zhang, R., and Hei, H. (2023). Advances in post-translational modifications of proteins and cancer immunotherapy. Front. Immunol. 14, 1229397. doi: 10.3389/fimmu.2023.1229397

Ling, X., Zhang, L., Fang, C., Liang, H., and Ma, J. (2024). A comprehensive prognostic and immunological implications of PFKP in pan-cancer. Cancer Cell Int 24, 310. doi: 10.1186/s12935-024-03497-w

Lv, X., Lv, Y., and Dai, X. (2023). Lactate, histone lactylation and cancer hallmarks. Expert Rev. Mol. Med. 25, e7. doi: 10.1017/erm.2022.42

Mees, S. T., Mardin, W. A., Wendel, C., Baeumer, N., Willscher, E., Senninger, N., et al. (2010). EP300—A miRNA-regulated metastasis suppressor gene in ductal adenocarcinomas of the pancreas. Intl Journal of Cancer 126, 114–124. doi: 10.1002/ijc.24695

Pan, S., Brentnall, T. A., and Chen, R. (2020). Proteome alterations in pancreatic ductal adenocarcinoma. Cancer Letters 469, 429–436. doi: 10.1016/j.canlet.2019.11.020

Popovici, V., and Ozon, E. A. (n.d.). Oral Cancer: Prophylaxis, Etiopathogenesis and Treatment.

Qiu, L., Gao, Q., Liao, Y., Li, X., and Li, C. (2025). Targeted inhibition of the PTEN/PI3K/AKT pathway by YSV induces cell cycle arrest and apoptosis in oral squamous cell carcinoma. J Transl Med 23, 145. doi: 10.1186/s12967-025-06169-z

Rho, H., Terry, A. R., Chronis, C., and Hay, N. (2023). Hexokinase 2-mediated gene expression via histone lactylation is required for hepatic stellate cell activation and liver fibrosis. Cell Metabolism 35, 1406–1423.e8. doi: 10.1016/j.cmet.2023.06.013

Rumgay, H. (n.d.). Global burden of oral cancer in 2022 attributable to smokeless tobacco and areca nut consumption: a population attributable fraction analysis.

Sanchez-Vega, F., Mina, M., Armenia, J., Chatila, W. K., Luna, A., La, K. C., et al. (2018). Oncogenic Signaling Pathways in The Cancer Genome Atlas. Cell 173, 321–337.e10. doi: 10.1016/j.cell.2018.03.035

Senga, S. S., and Grose, R. P. (2021). Hallmarks of cancer—the new testament. Open Biol. 11, 200358. doi: 10.1098/rsob.200358

Siegel, R. L., Miller, K. D., and Jemal, A. (2019). Cancer statistics, 2019. CA CANCER J CLIN.

Tan, Y. (2023). Oral squamous cell carcinomas: state of the field and emerging directions. International Journal of Oral Science.

The Multiple Tissue Human Expression Resource (MuTHER) Consortium, Grundberg, E., Small, K. S., Hedman, Å. K., Nica, A. C., Buil, A., et al. (2012). Mapping cis- and trans-regulatory effects across multiple tissues in twins. Nat Genet 44, 1084–1089. doi: 10.1038/ng.2394

Tuo, L., Xiang, J., Pan, X., Gao, Q., Zhang, G., Yang, Y., et al. (2018). PCK1 Downregulation Promotes TXNRD1 Expression and Hepatoma Cell Growth via the Nrf2/Keap1 Pathway. Front. Oncol. 8, 611. doi: 10.3389/fonc.2018.00611

Wang, Z., and Dong, C. (2019). Gluconeogenesis in Cancer: Function and Regulation of PEPCK, FBPase, and G6Pase. Trends in Cancer 5, 30–45. doi: 10.1016/j.trecan.2018.11.003

Wu, Y., Zeng, J., Zhang, F., Zhu, Z., Qi, T., Zheng, Z., et al. (2018). Integrative analysis of omics summary data reveals putative mechanisms underlying complex traits. Nat Commun 9, 918. doi: 10.1038/s41467-018-03371-0

Wu, Z., Huang, R., and Yuan, L. (2019). Crosstalk of intracellular post-translational modifications in cancer. Archives of Biochemistry and Biophysics 676, 108138. doi: 10.1016/j.abb.2019.108138

Yang, H., Zou, X., Yang, S., Zhang, A., Li, N., and Ma, Z. (2023). Identification of lactylation related model to predict prognostic, tumor infiltrating immunocytes and response of immunotherapy in gastric cancer. Front. Immunol. 14, 1149989. doi: 10.3389/fimmu.2023.1149989

Yu, J., Chai, P., Xie, M., Ge, S., Ruan, J., Fan, X., et al. (2021). Histone lactylation drives oncogenesis by facilitating m6A reader protein YTHDF2 expression in ocular melanoma. Genome Biol 22, 85. doi: 10.1186/s13059-021-02308-z

Zhang, D., Tang, Z., Huang, H., Zhou, G., Cui, C., Weng, Y., et al. (2019). Metabolic regulation of gene expression by histone lactylation. Nature 574, 575–580. doi: 10.1038/s41586-019-1678-1

Zhengdong, A., Xiaoying, X., Shuhui, F., Rui, L., Zehui, T., Guanbin, S., et al. (2024). Identification of fatty acids synthesis and metabolism-related gene signature and prediction of prognostic model in hepatocellular carcinoma. Cancer Cell Int 24, 130. doi: 10.1186/s12935-024-03306-4

Zhu, G., Pei, L., Li, Y., and Gou, X. (2020). EP300 mutation is associated with tumor mutation burden and promotes antitumor immunity in bladder cancer patients. Aging 12, 2132–2141. doi: 10.18632/aging.102728

