## Supplementary figures and images for "Multi-level Regulatory Roles of Lactate Metabolism Gene Network in Oral Cancer: Machine Learning Insights"

### Differential Expression Analysis with GEO.jpg

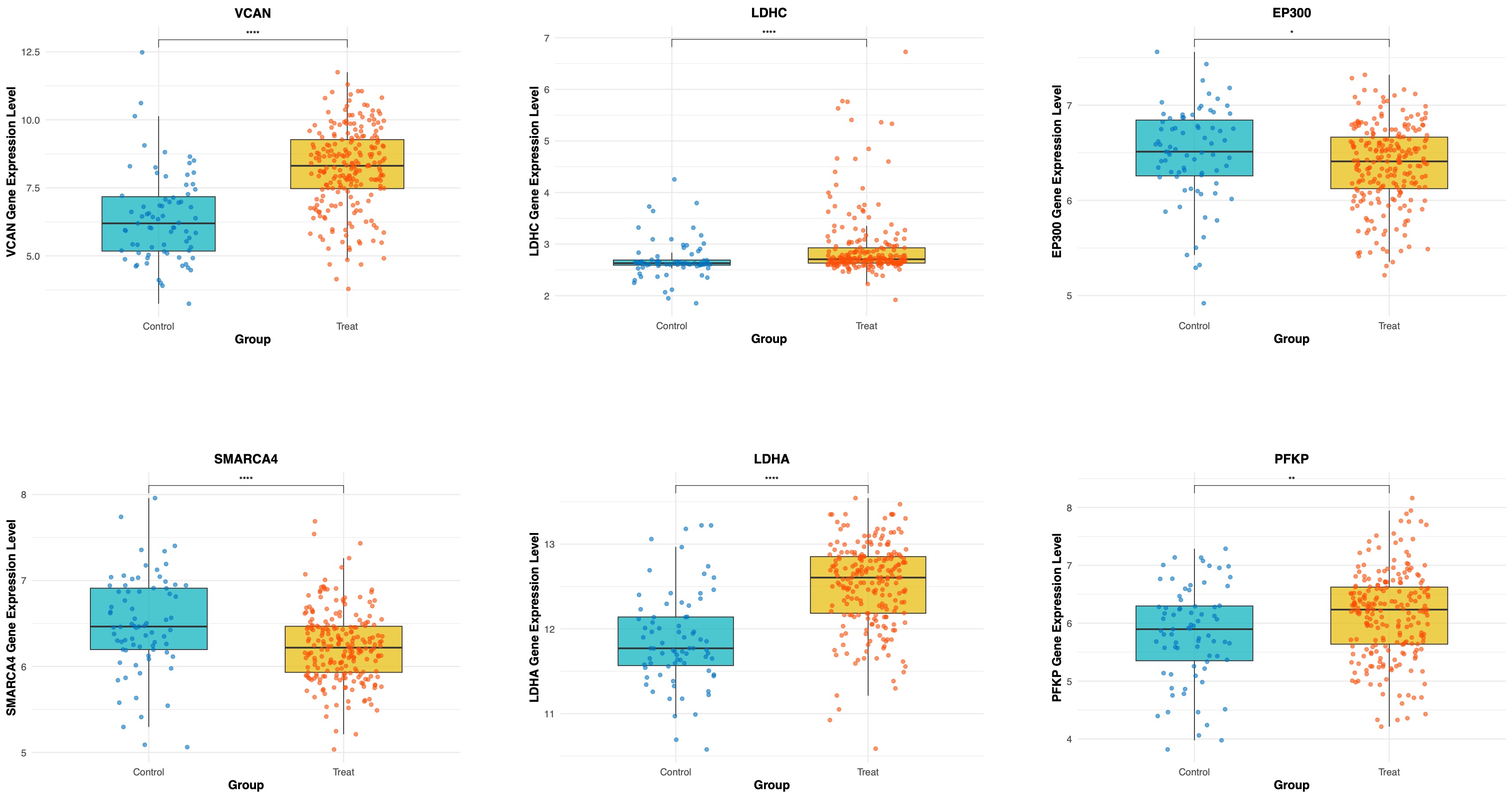

### eQTL_lactylation.pdf

Overlap of eQTL and Lactylation Genes

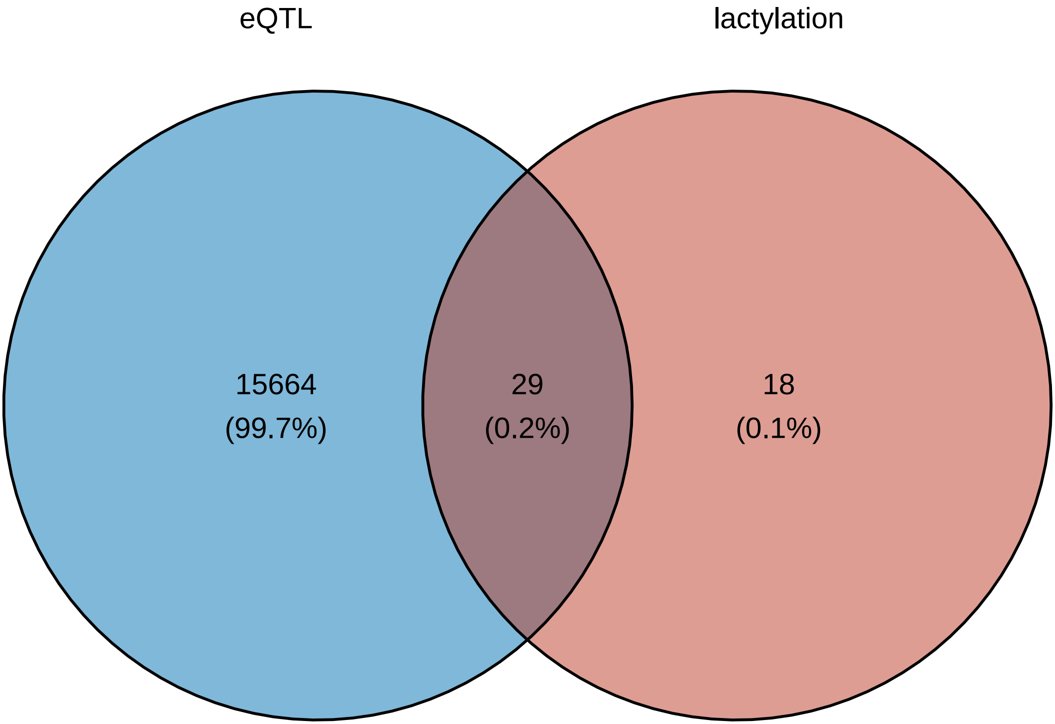

### forest.pdf

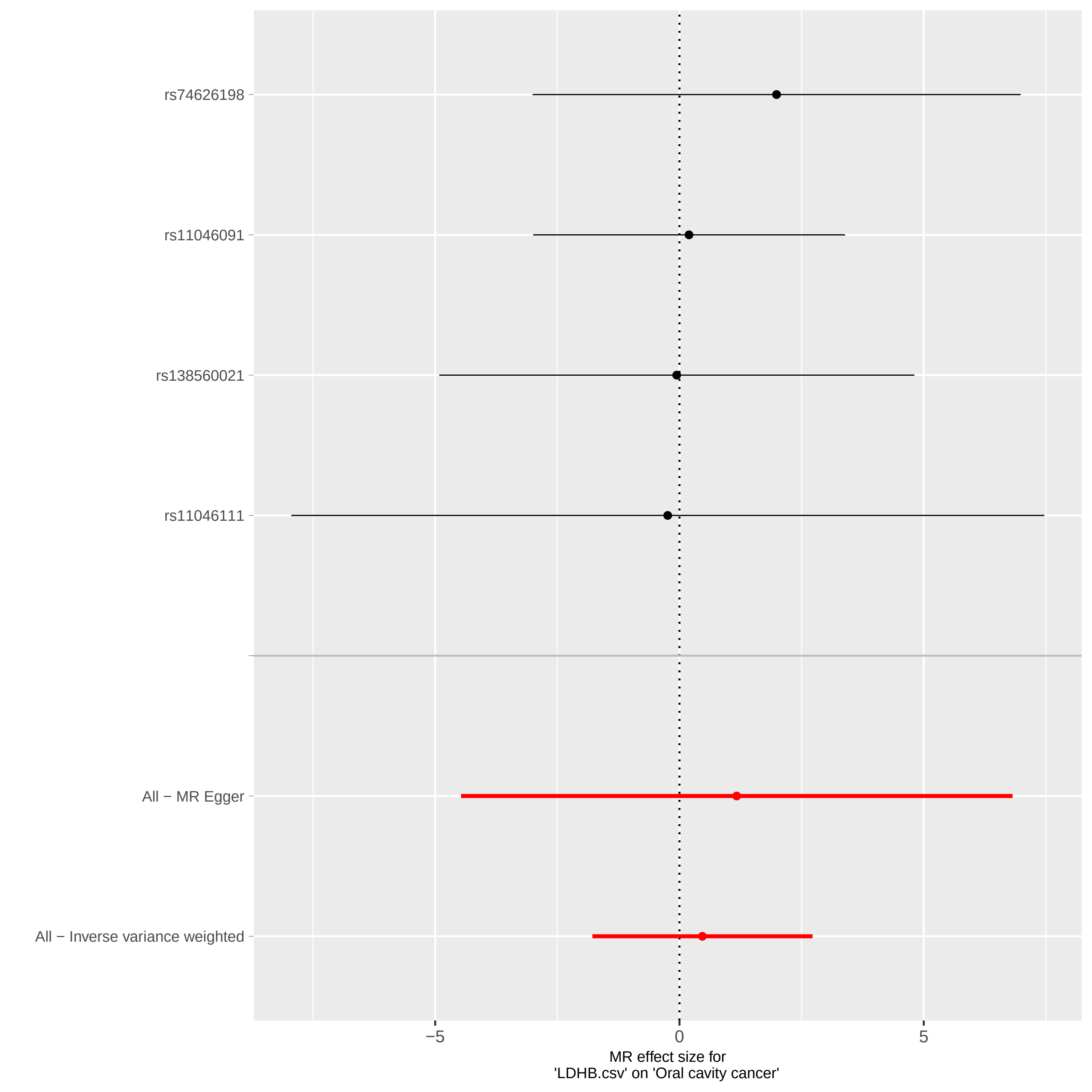

### forest.pdf

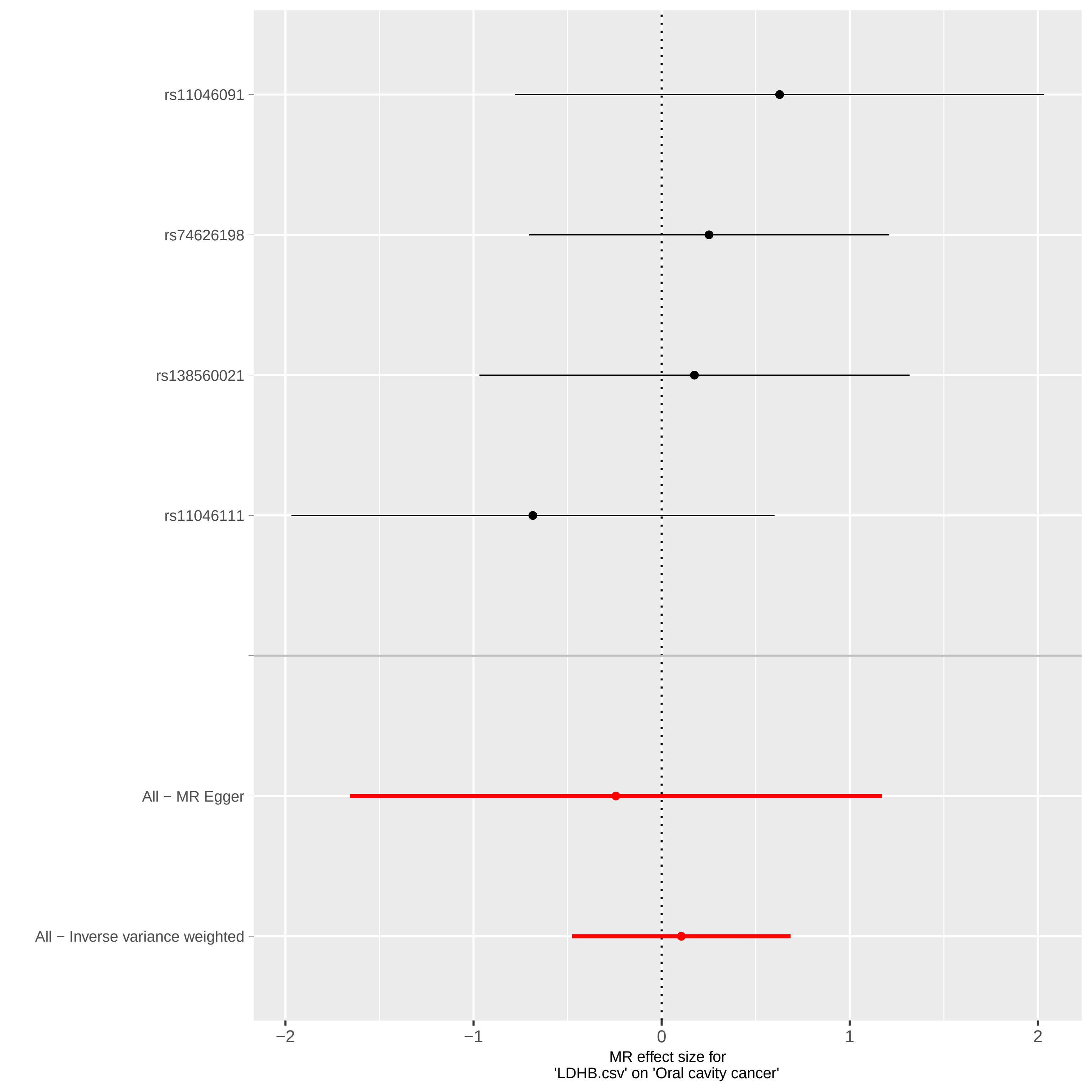

### forest.pdf

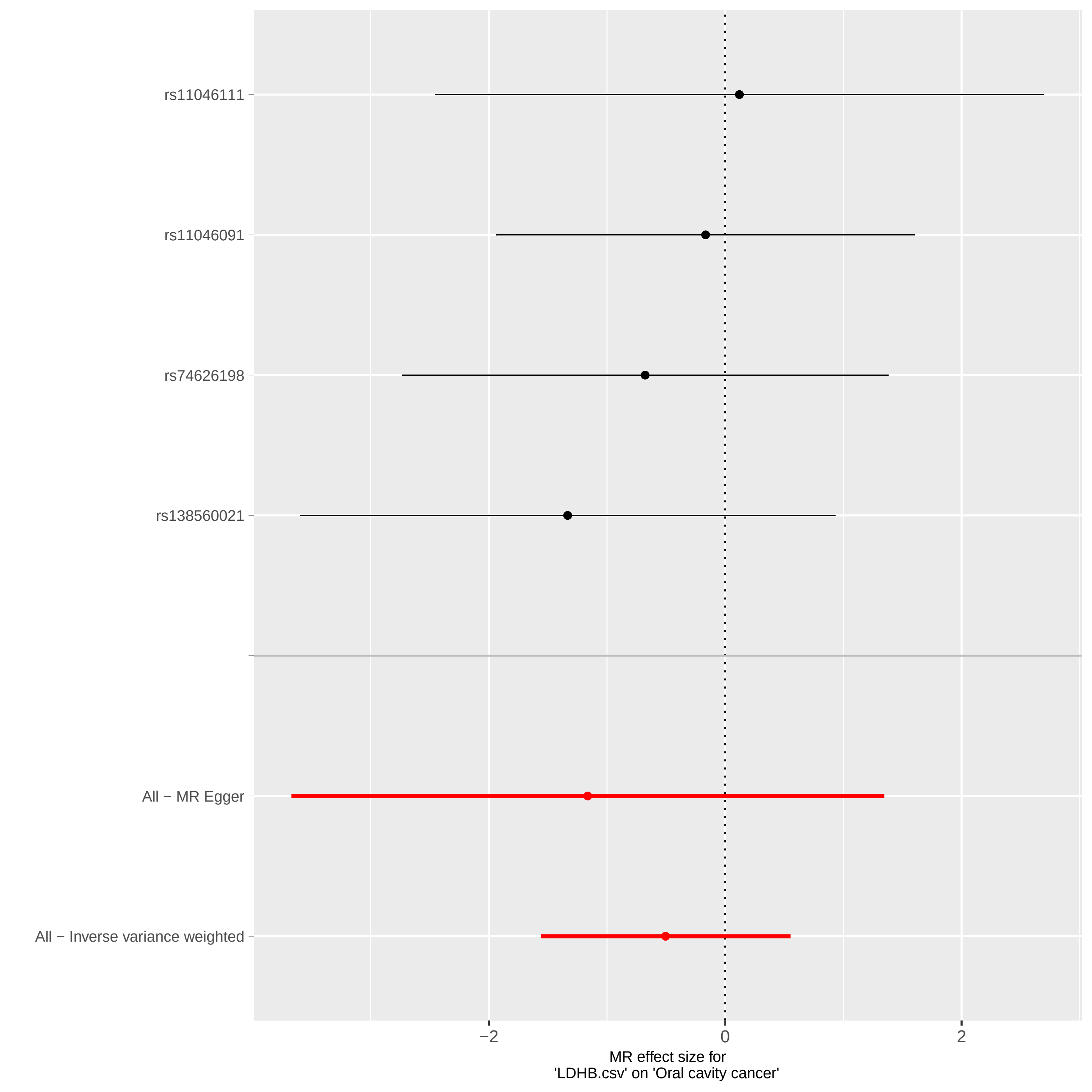

### forest.pdf

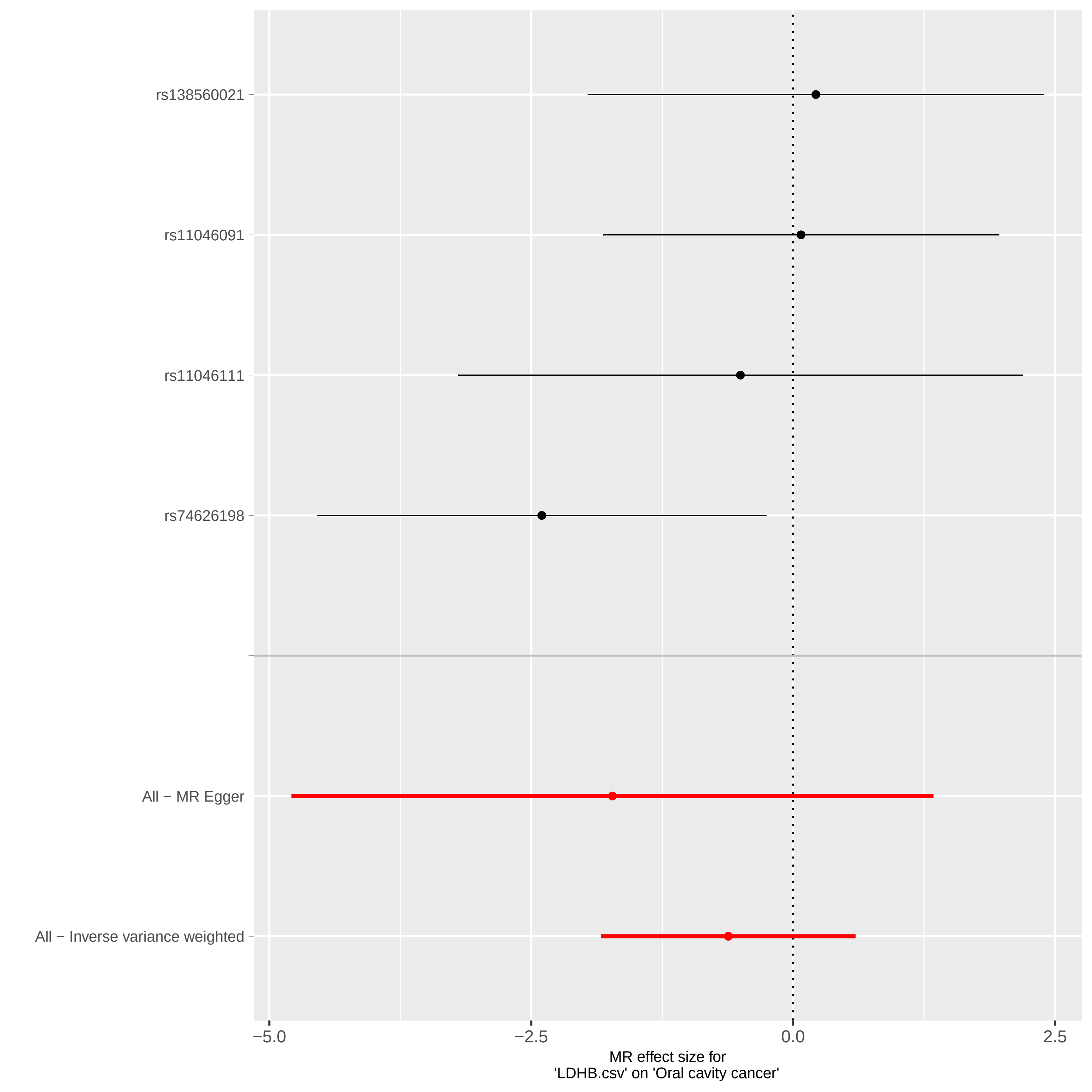

### forest.pdf

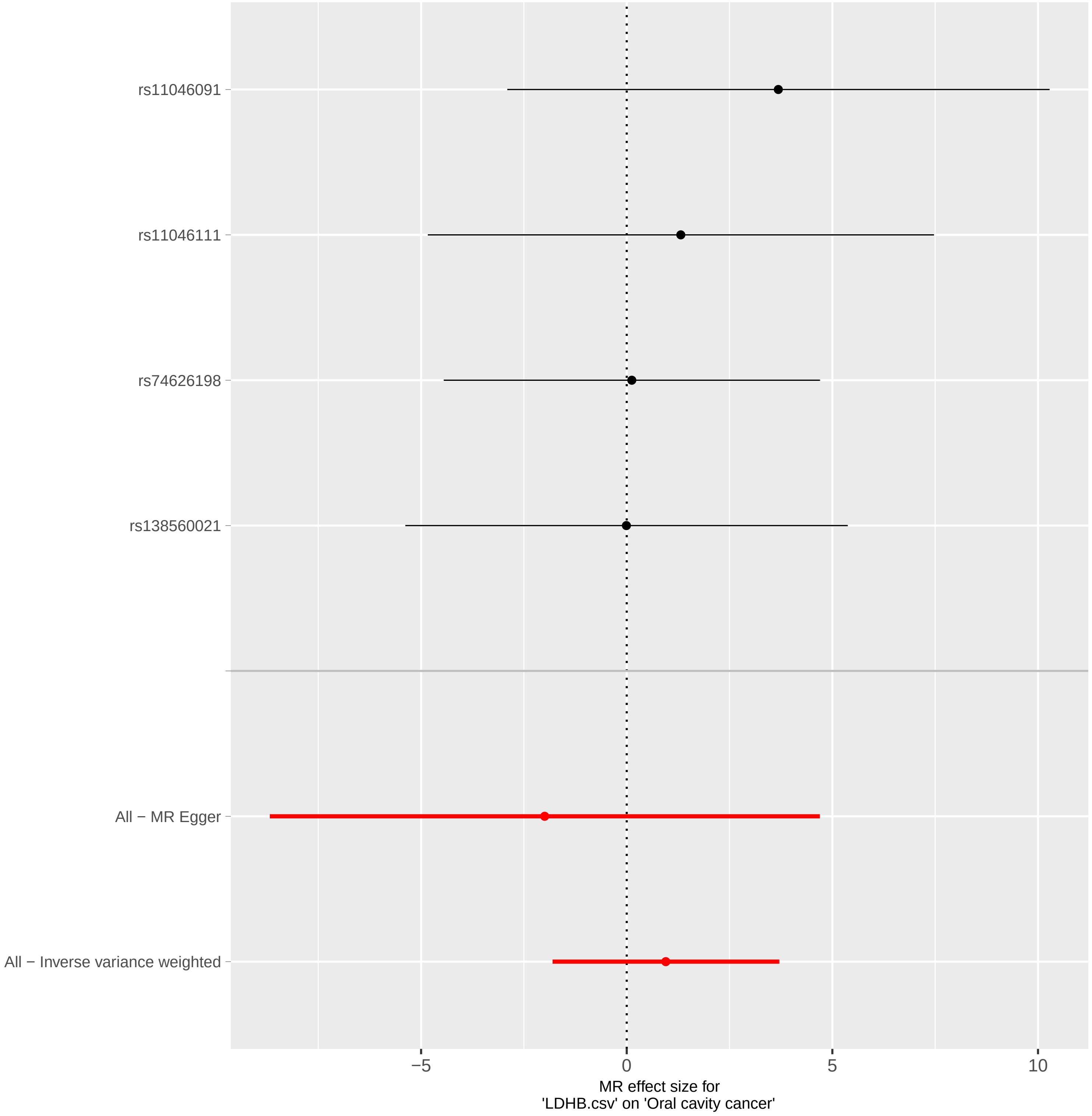

### forest.pdf

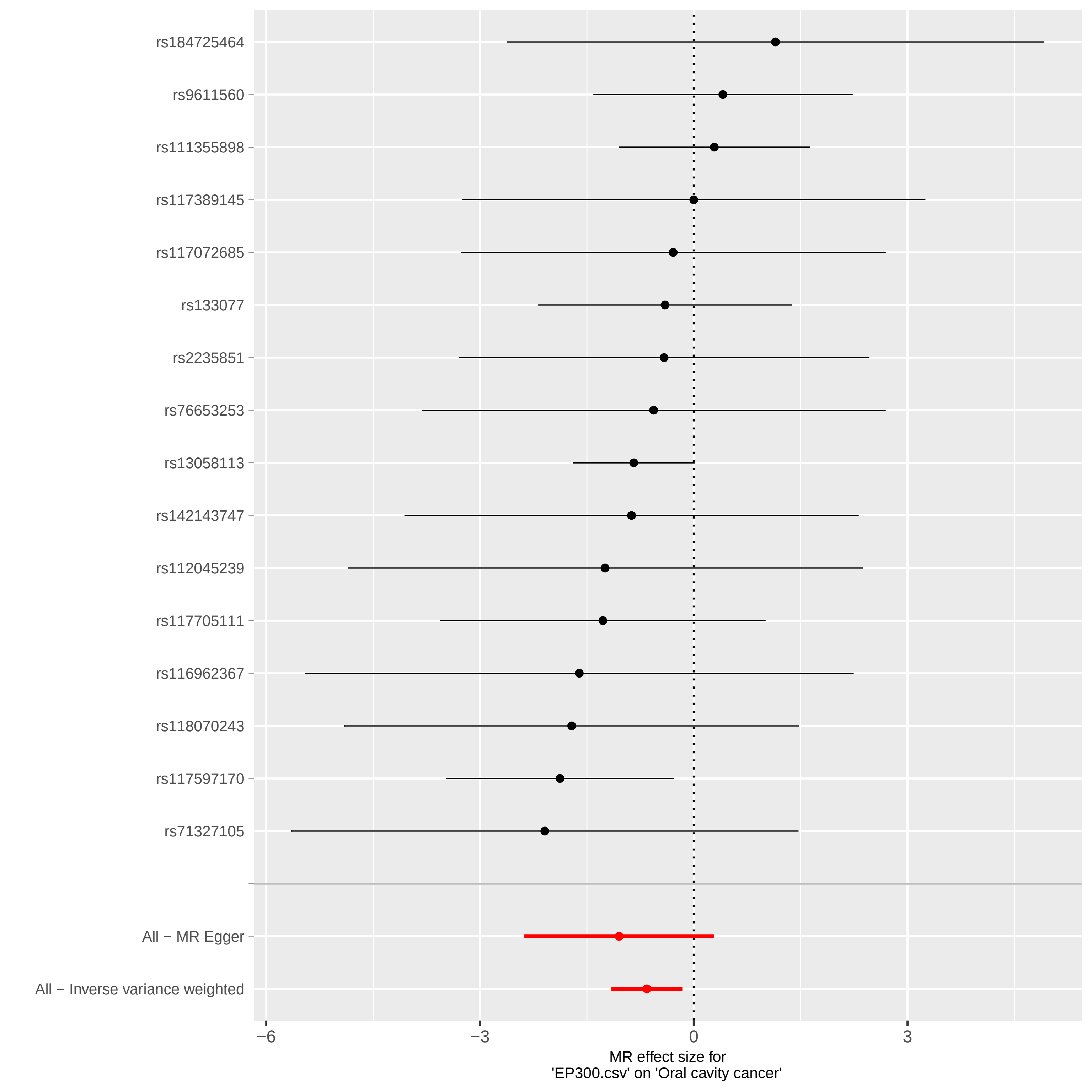

### forest.pdf

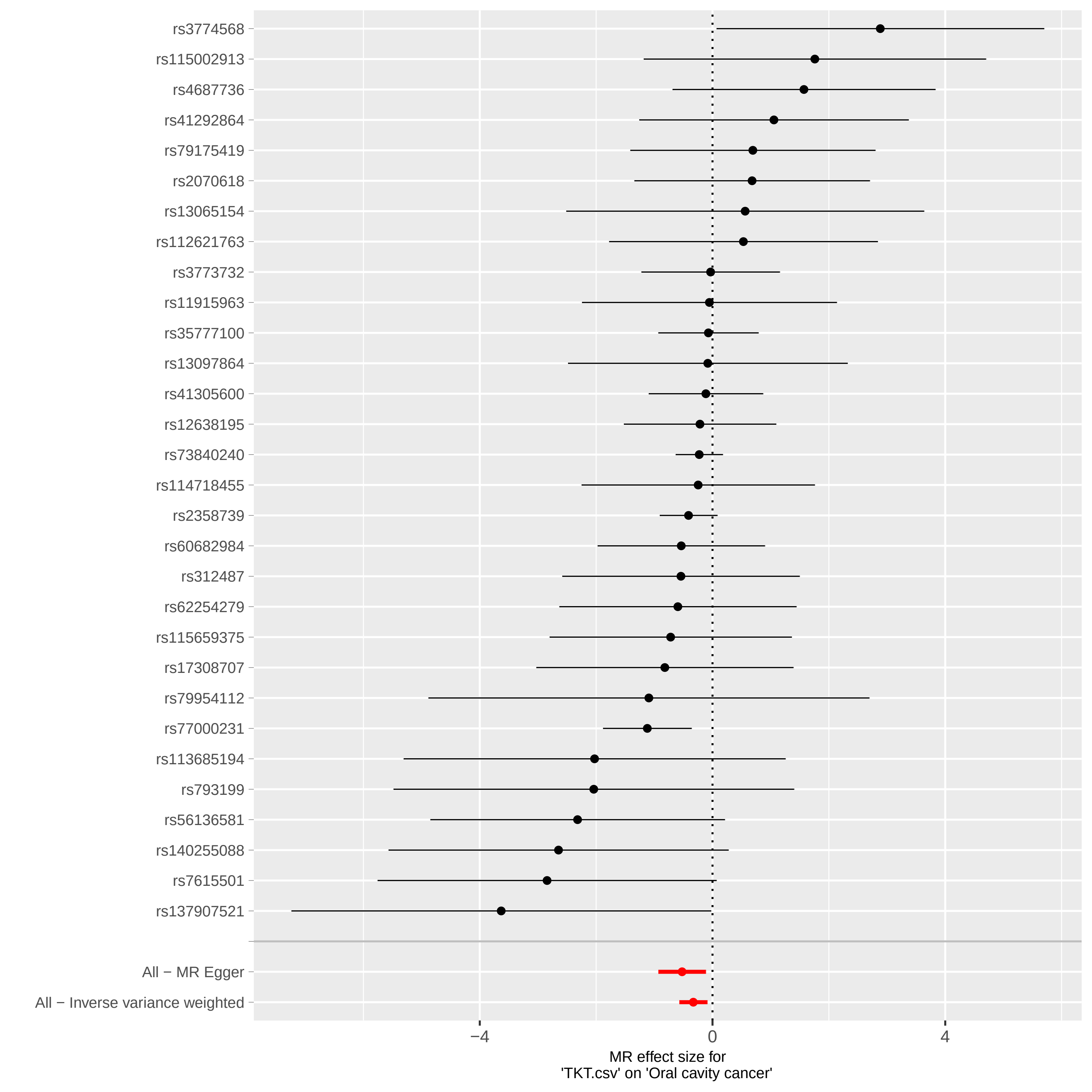

### forest.pdf

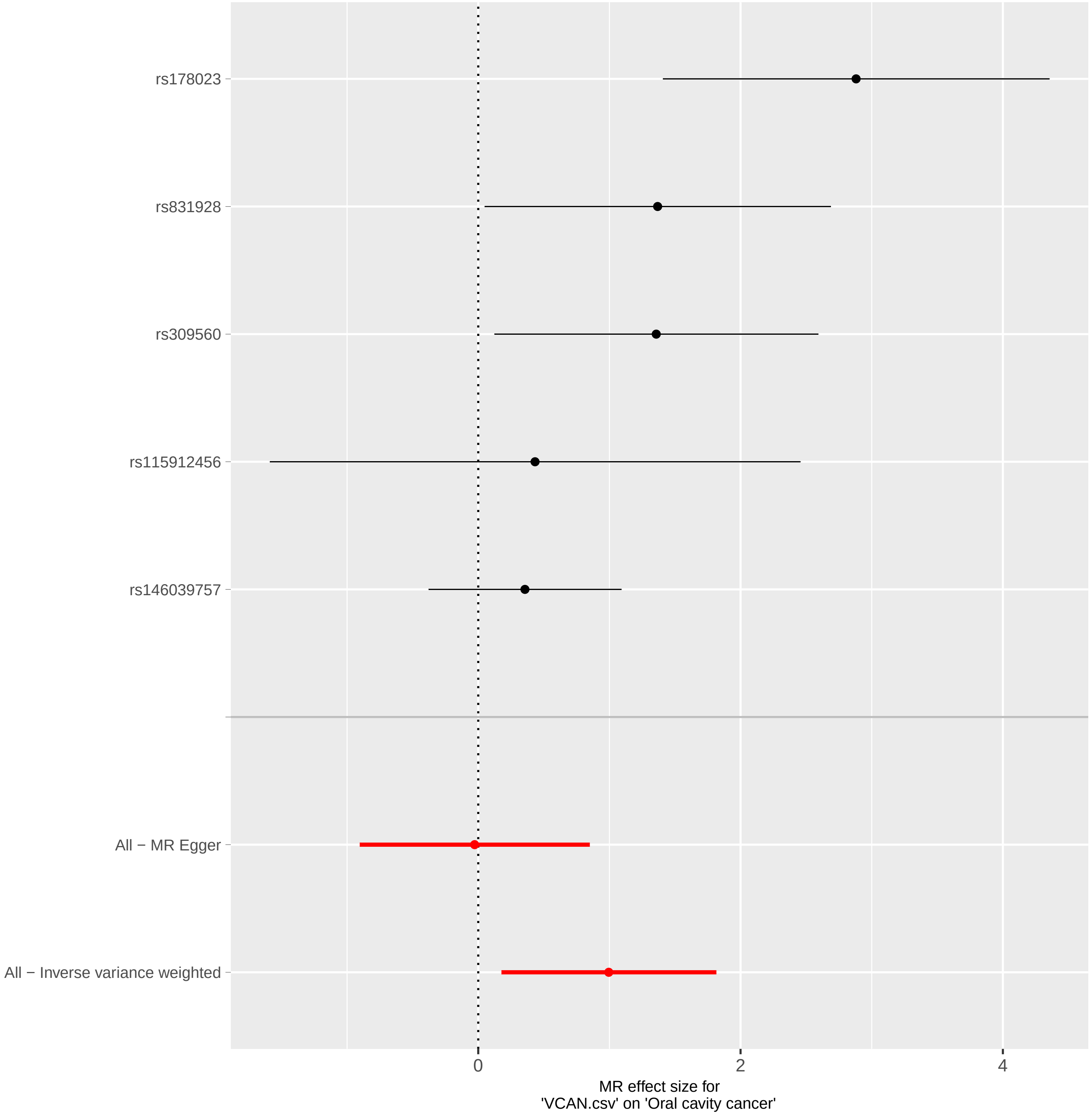

### forest.pdf

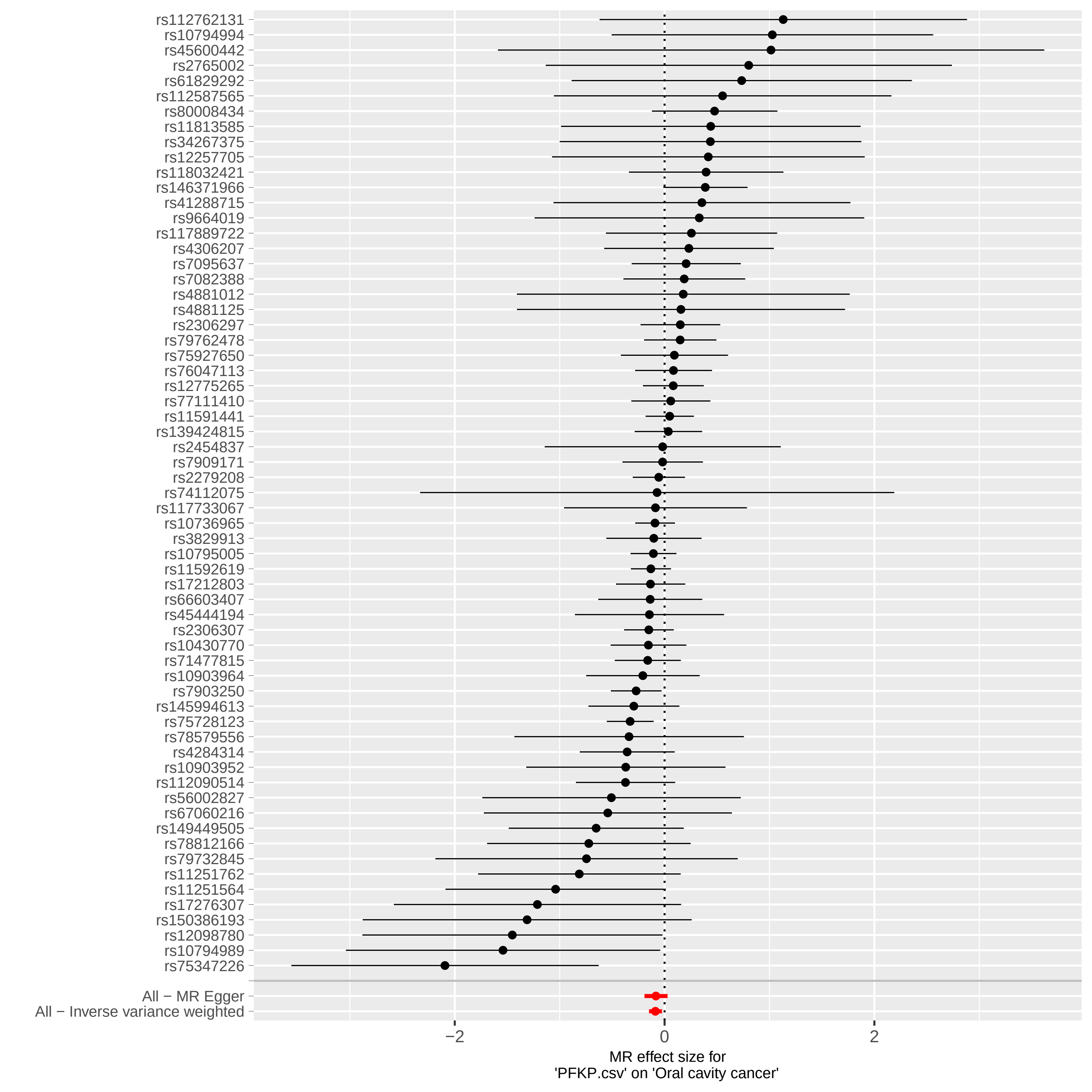

### forest.pdf

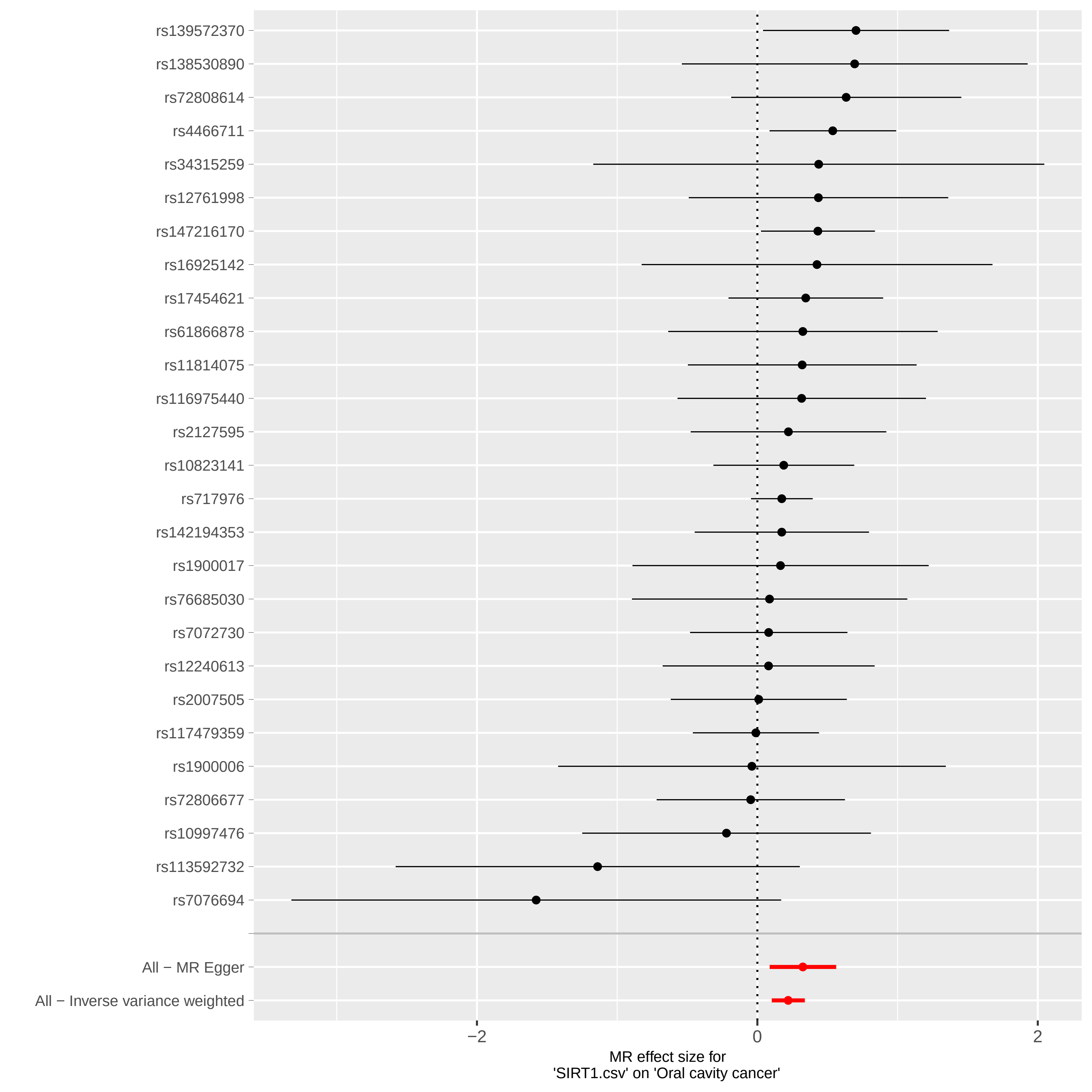

### forest.pdf

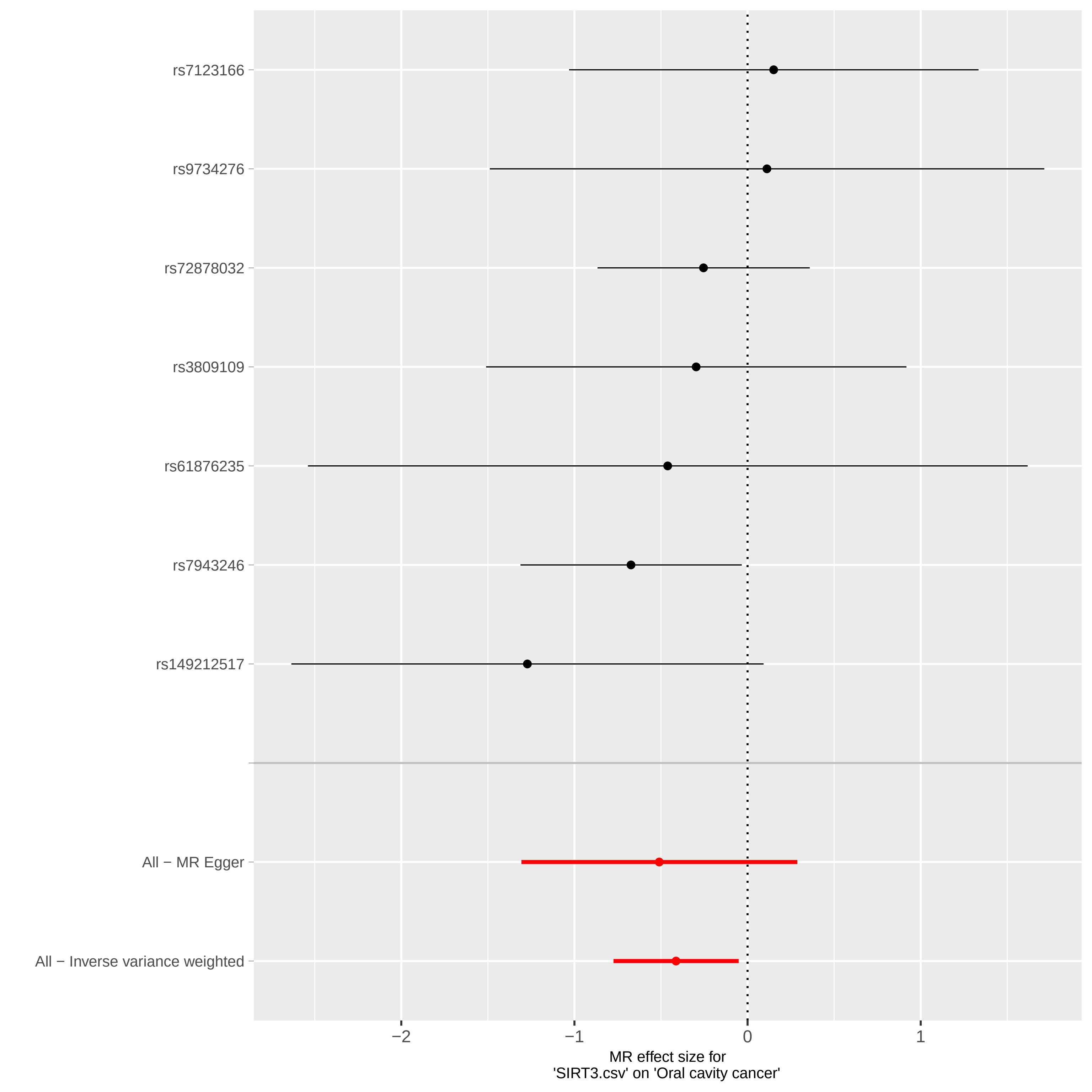

### forest.pdf

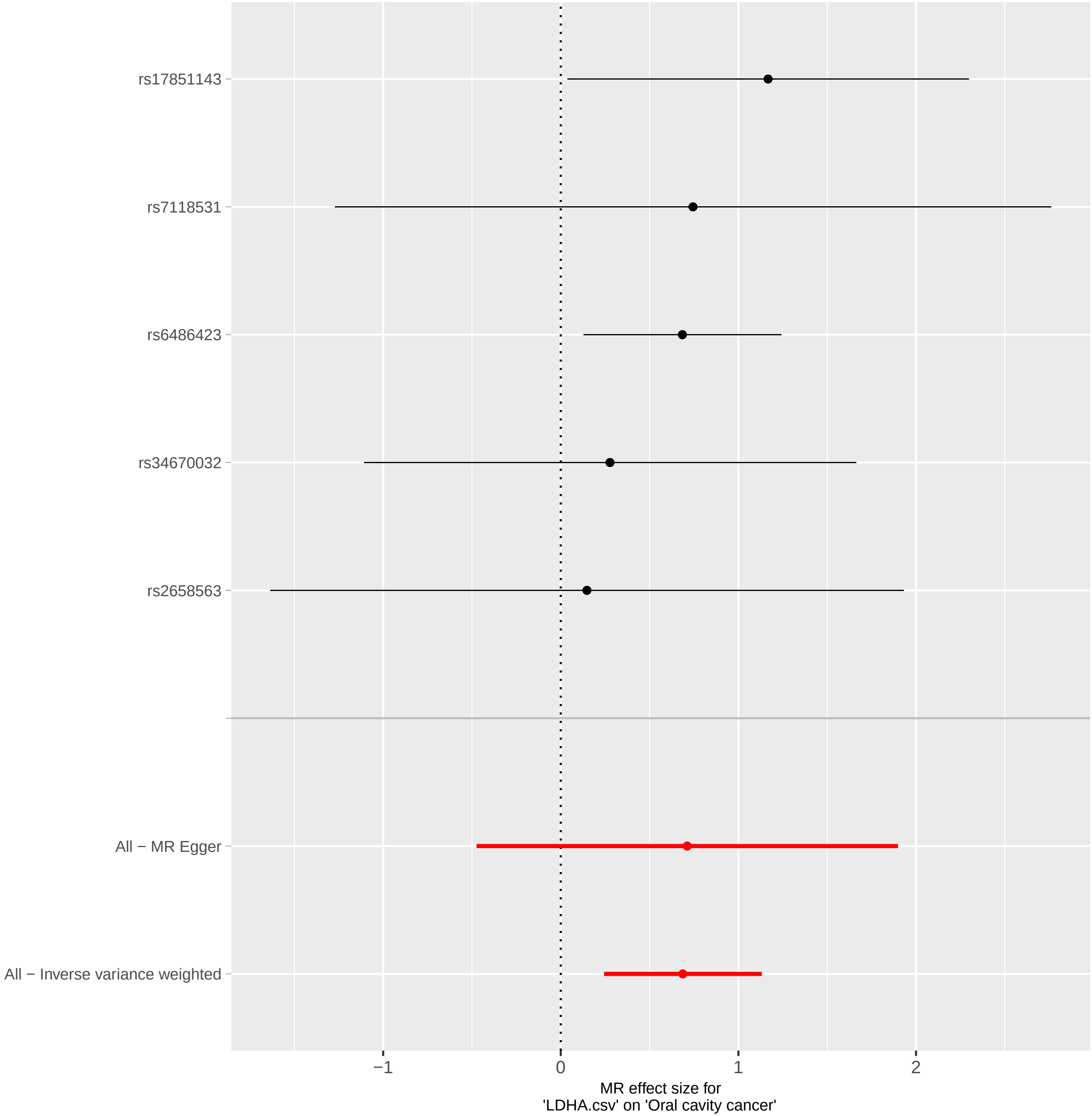

### forest.pdf

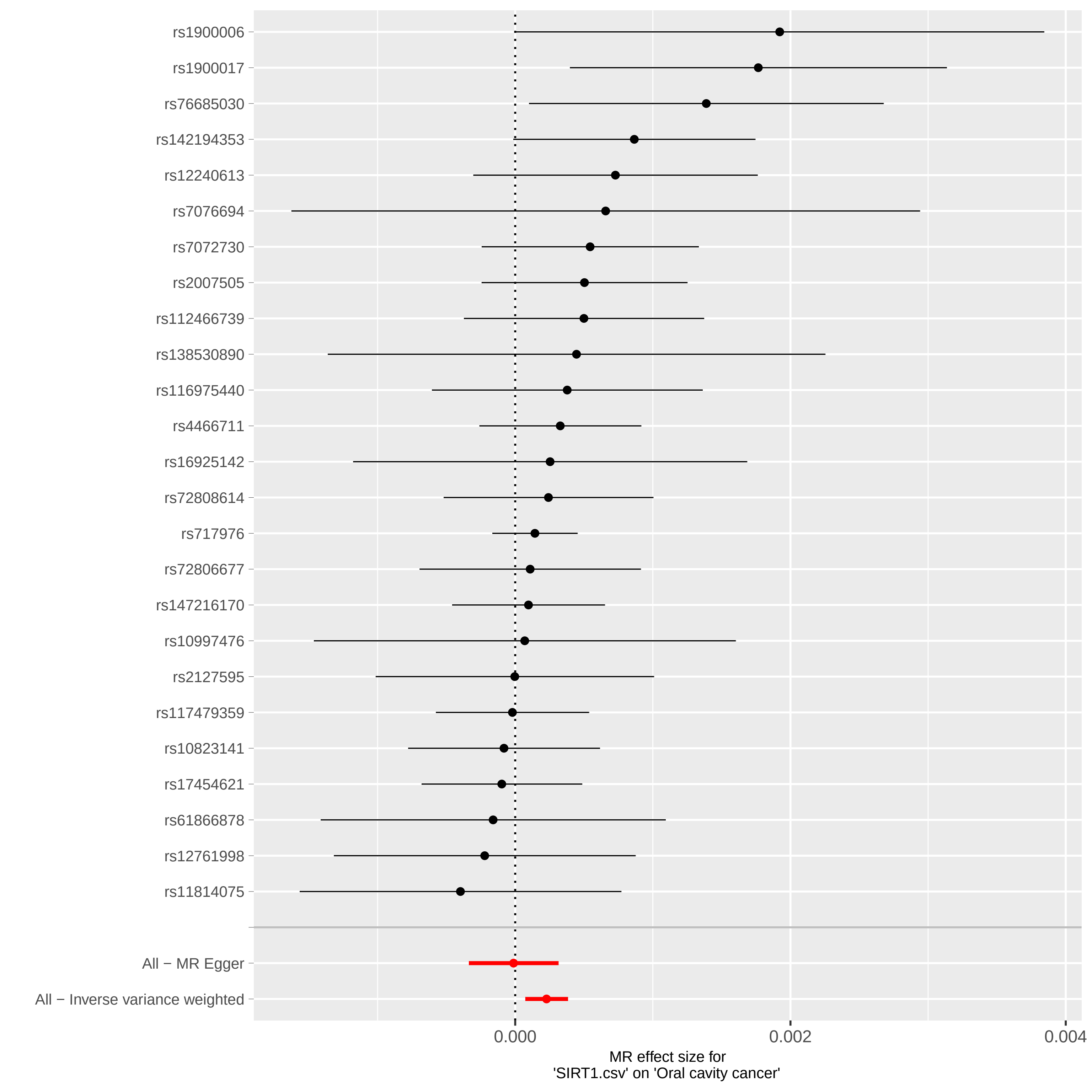

### forest.pdf

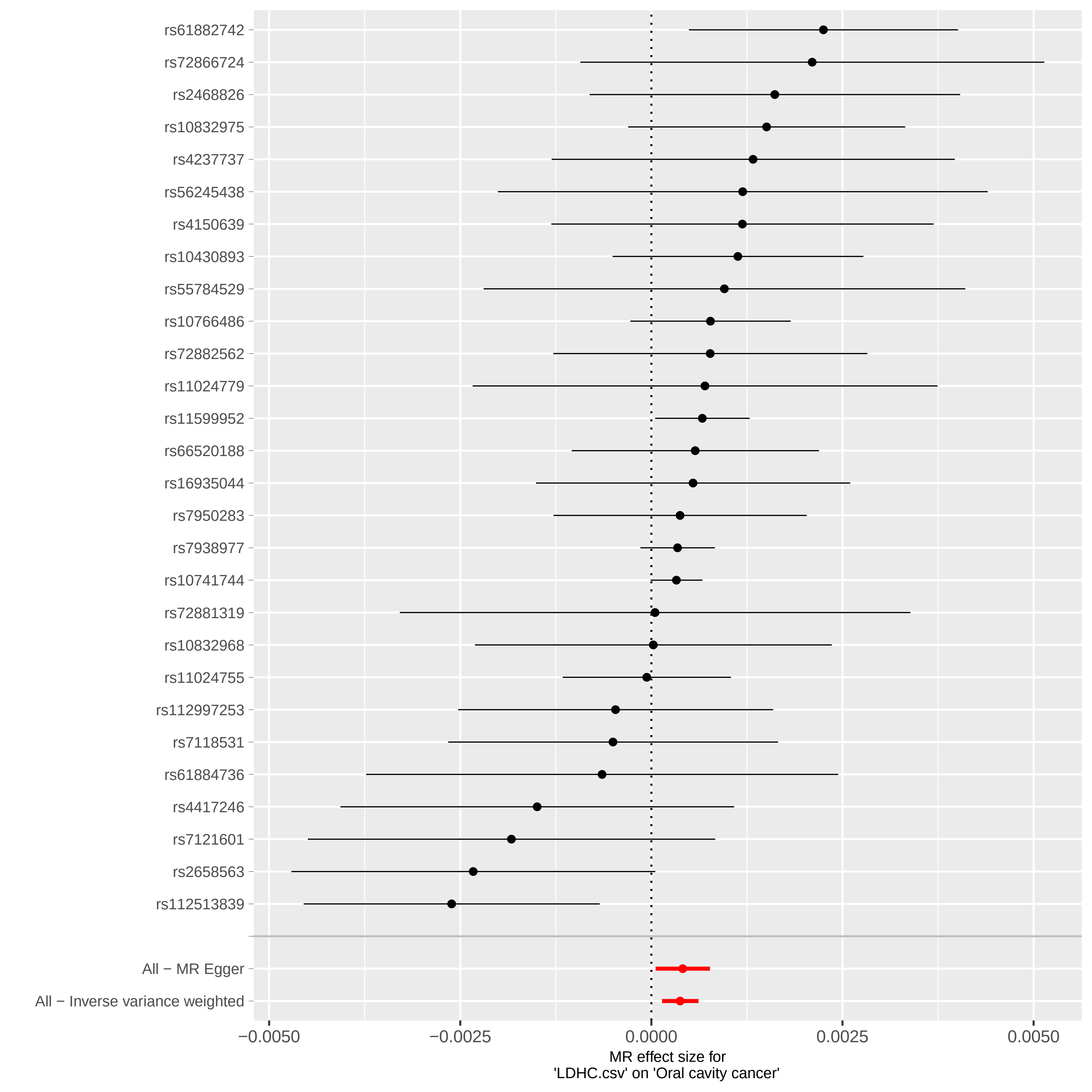

### forest.pdf

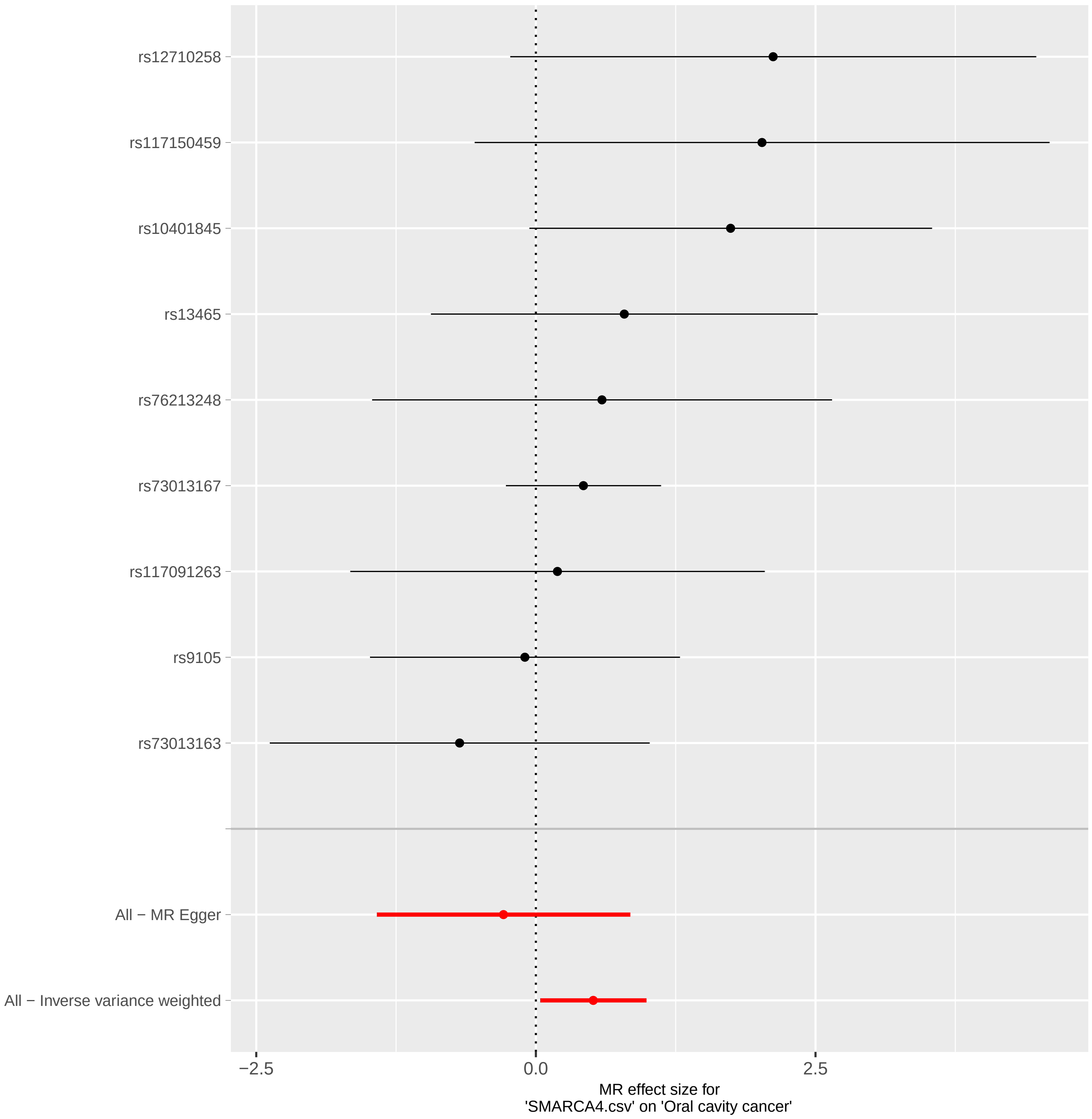

### forest.pdf

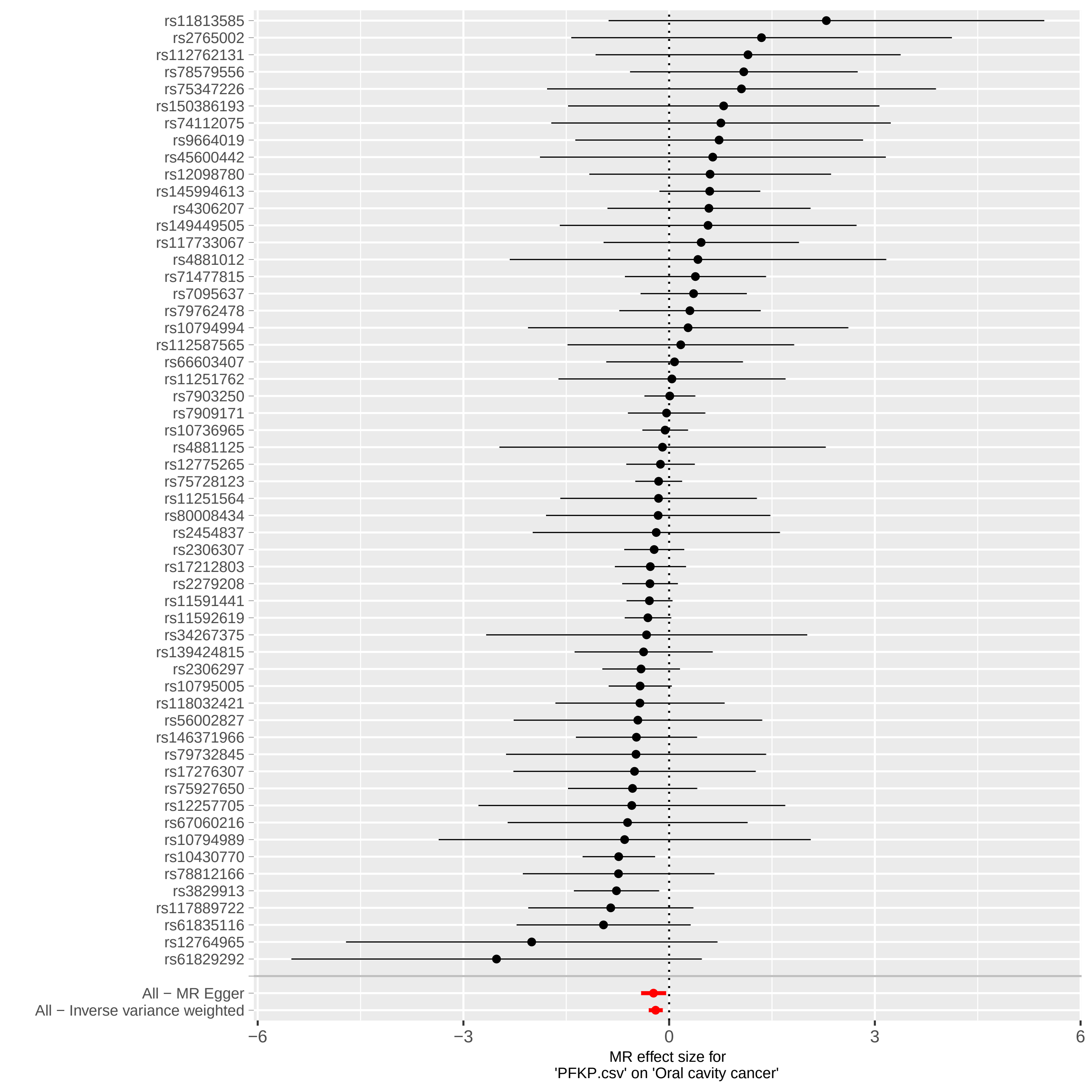

### forest.pdf

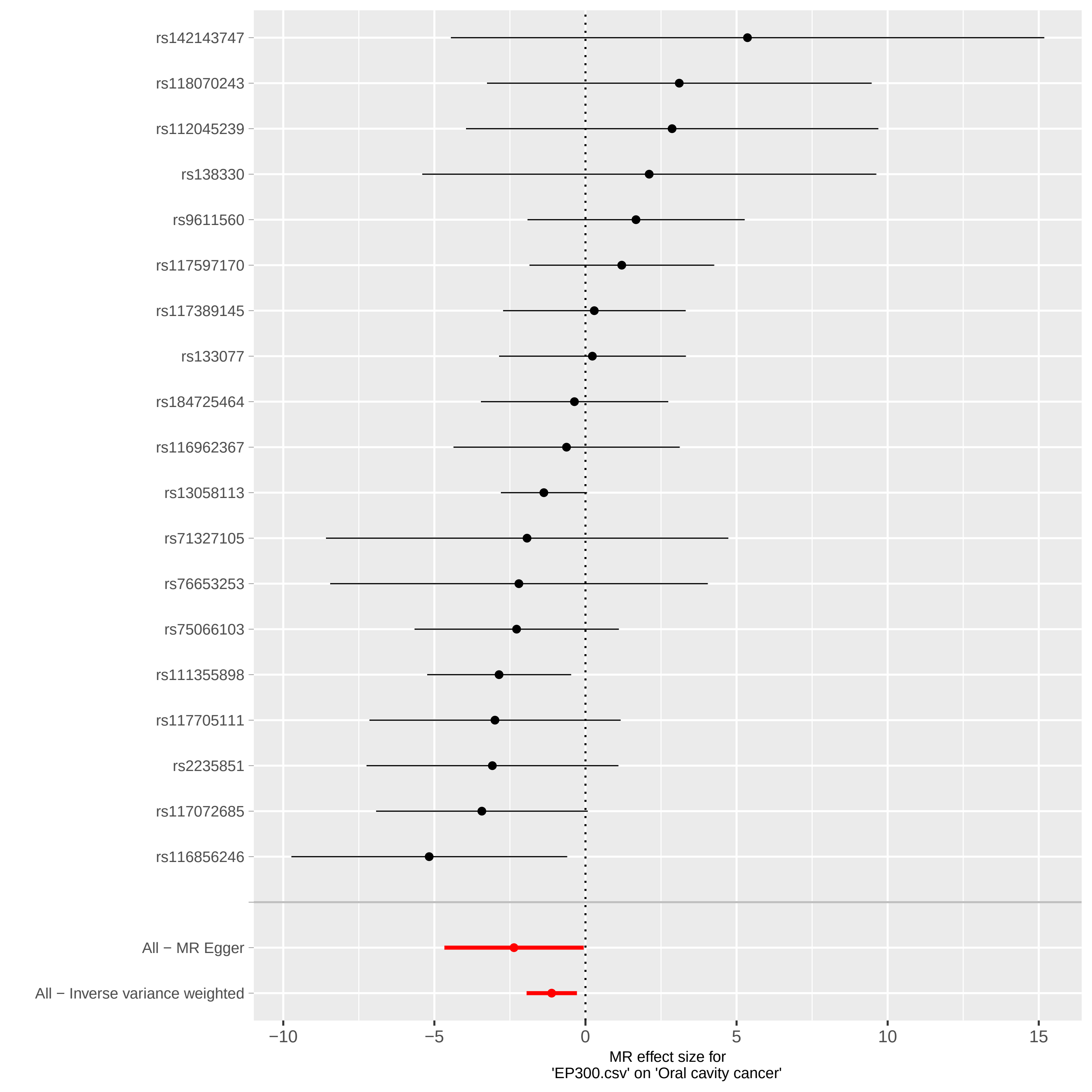

### forest.pdf

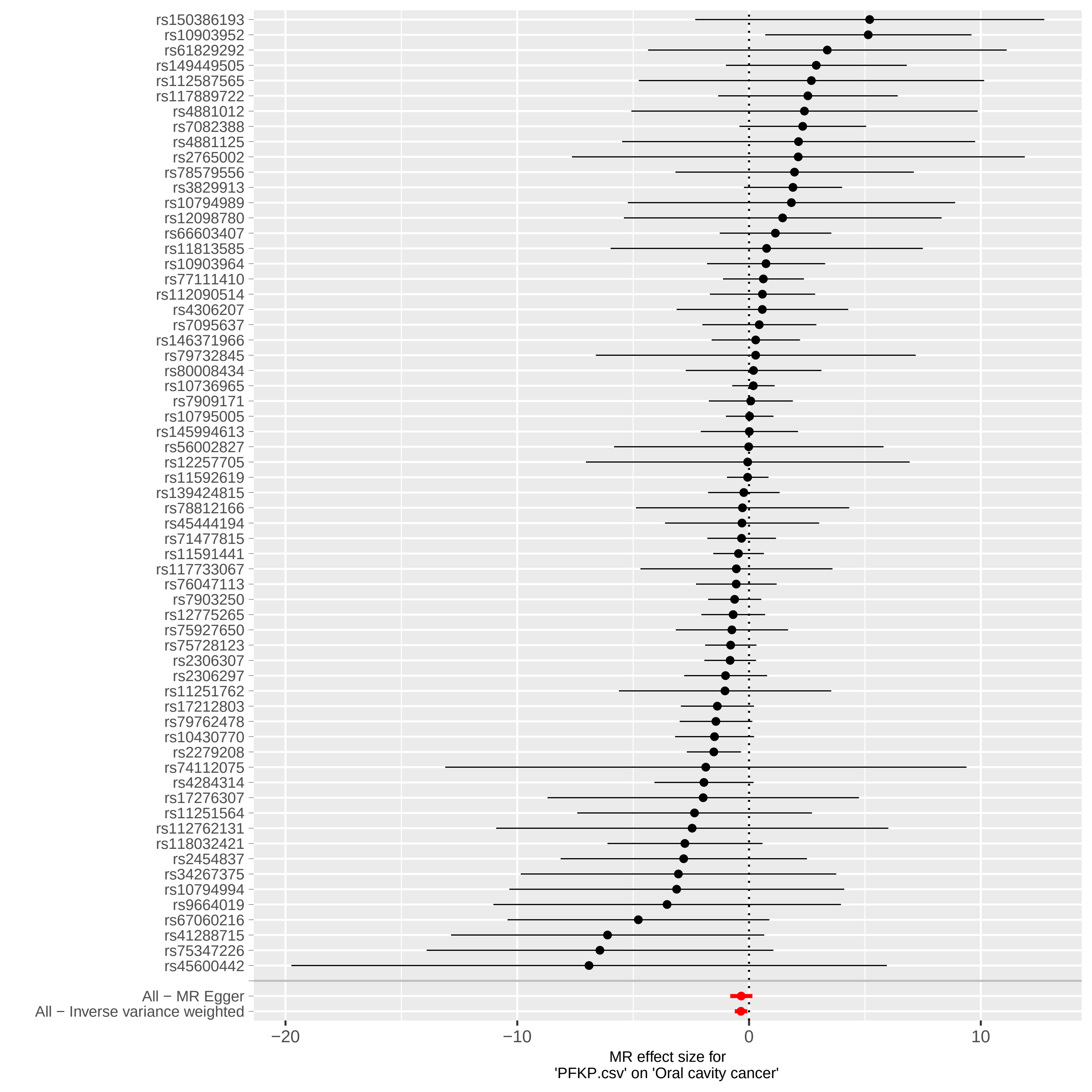

### funnelplot.pdf

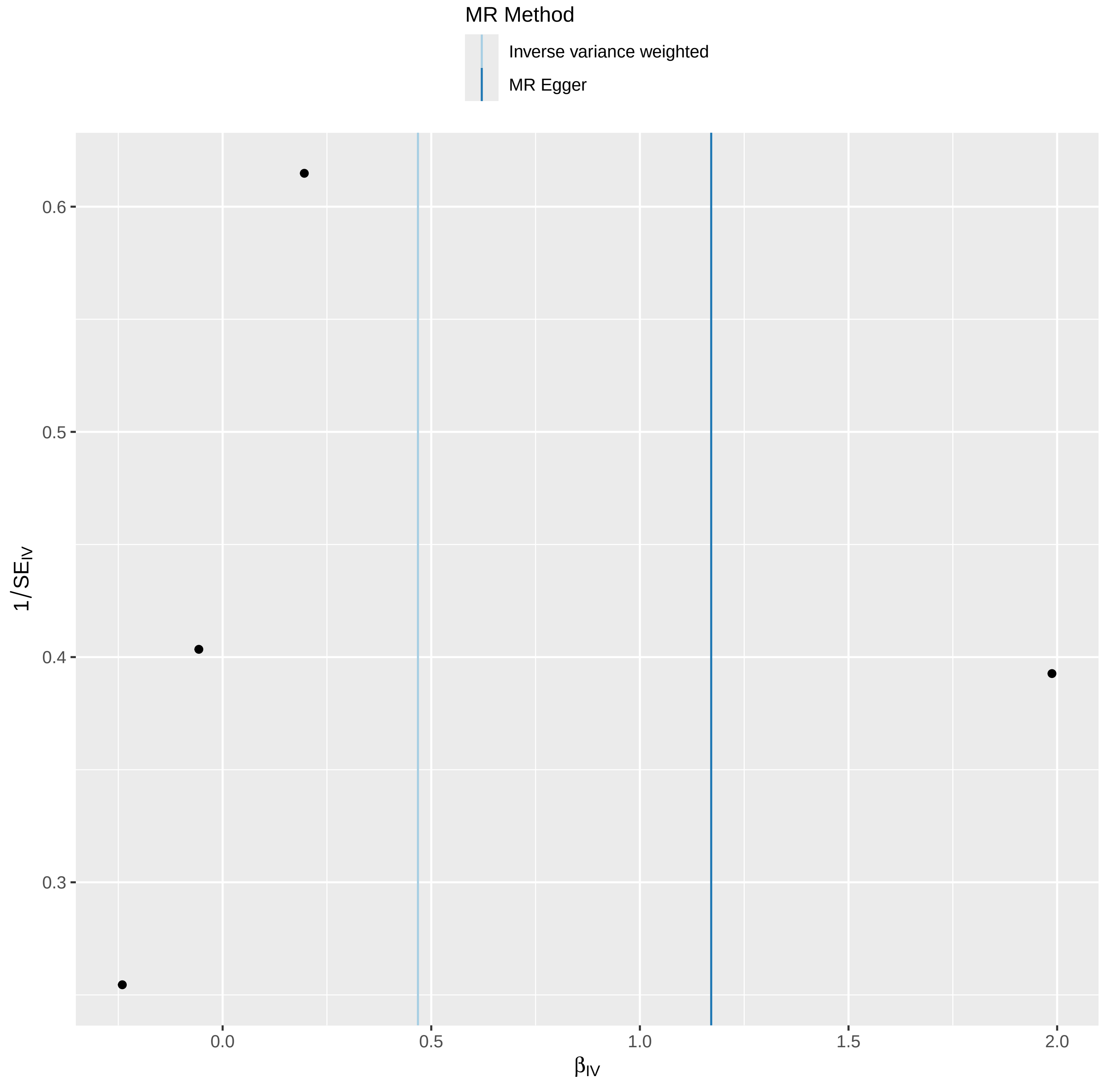

### funnelplot.pdf

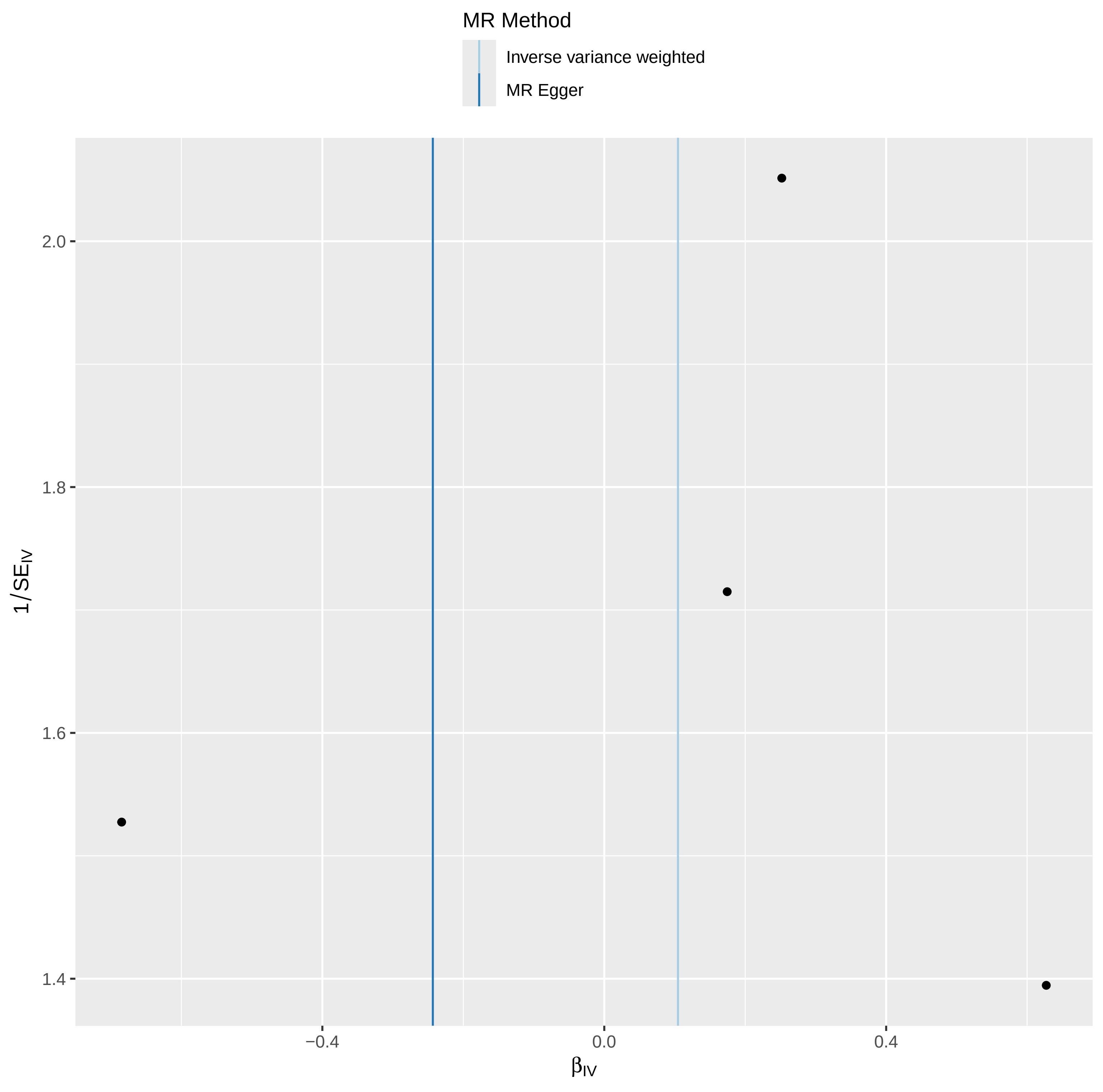

### funnelplot.pdf

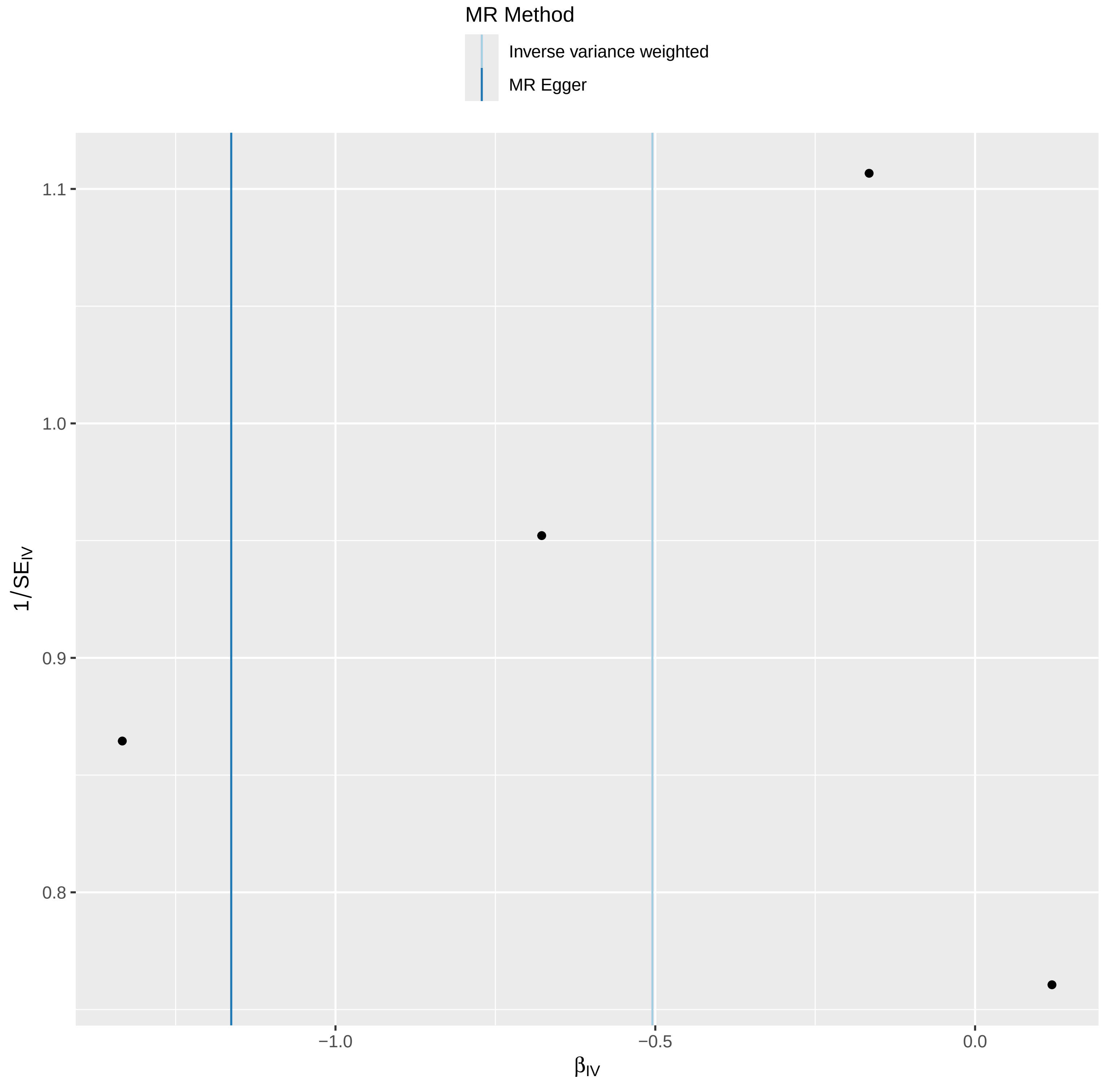

### funnelplot.pdf

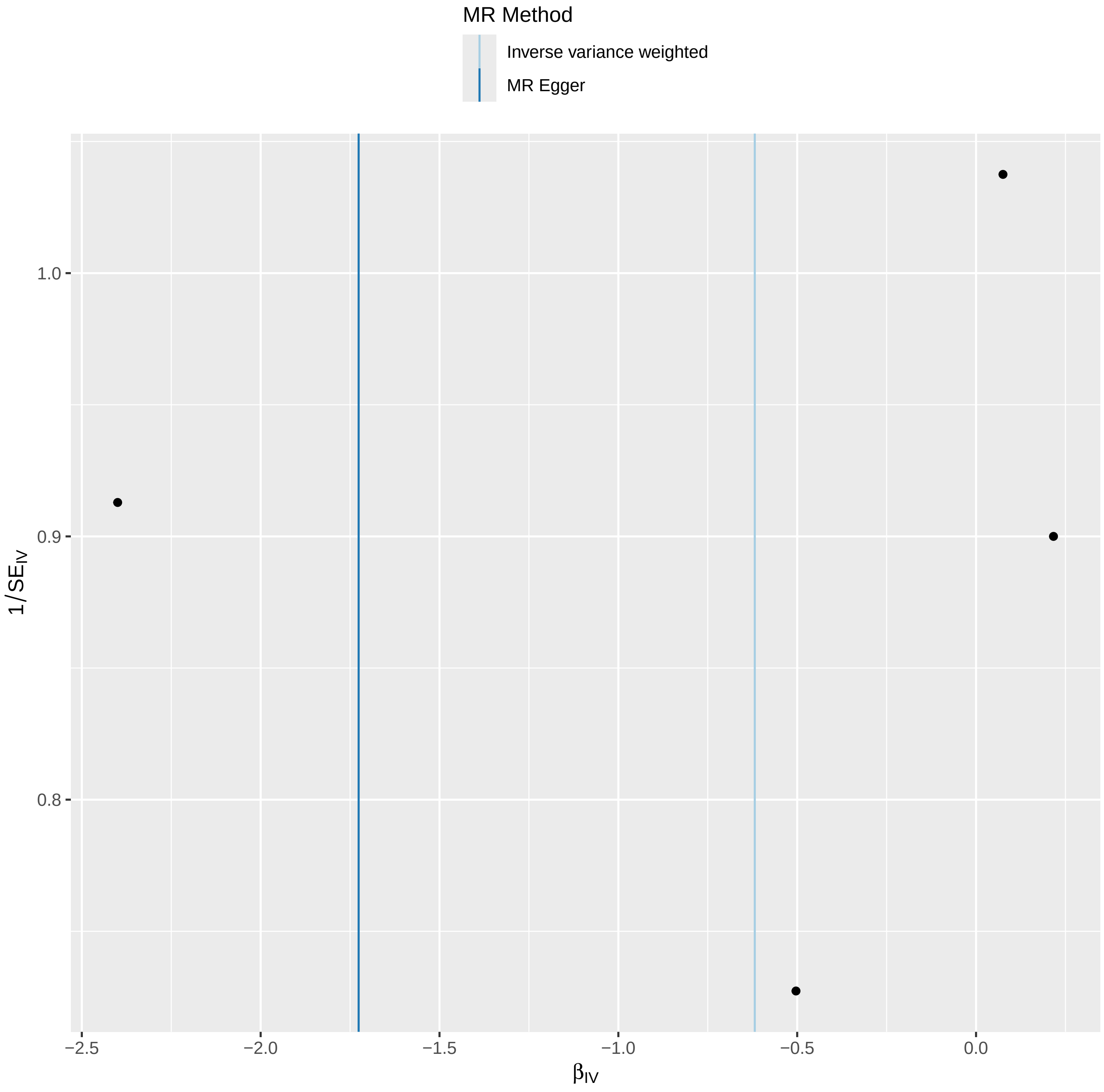

### funnelplot.pdf

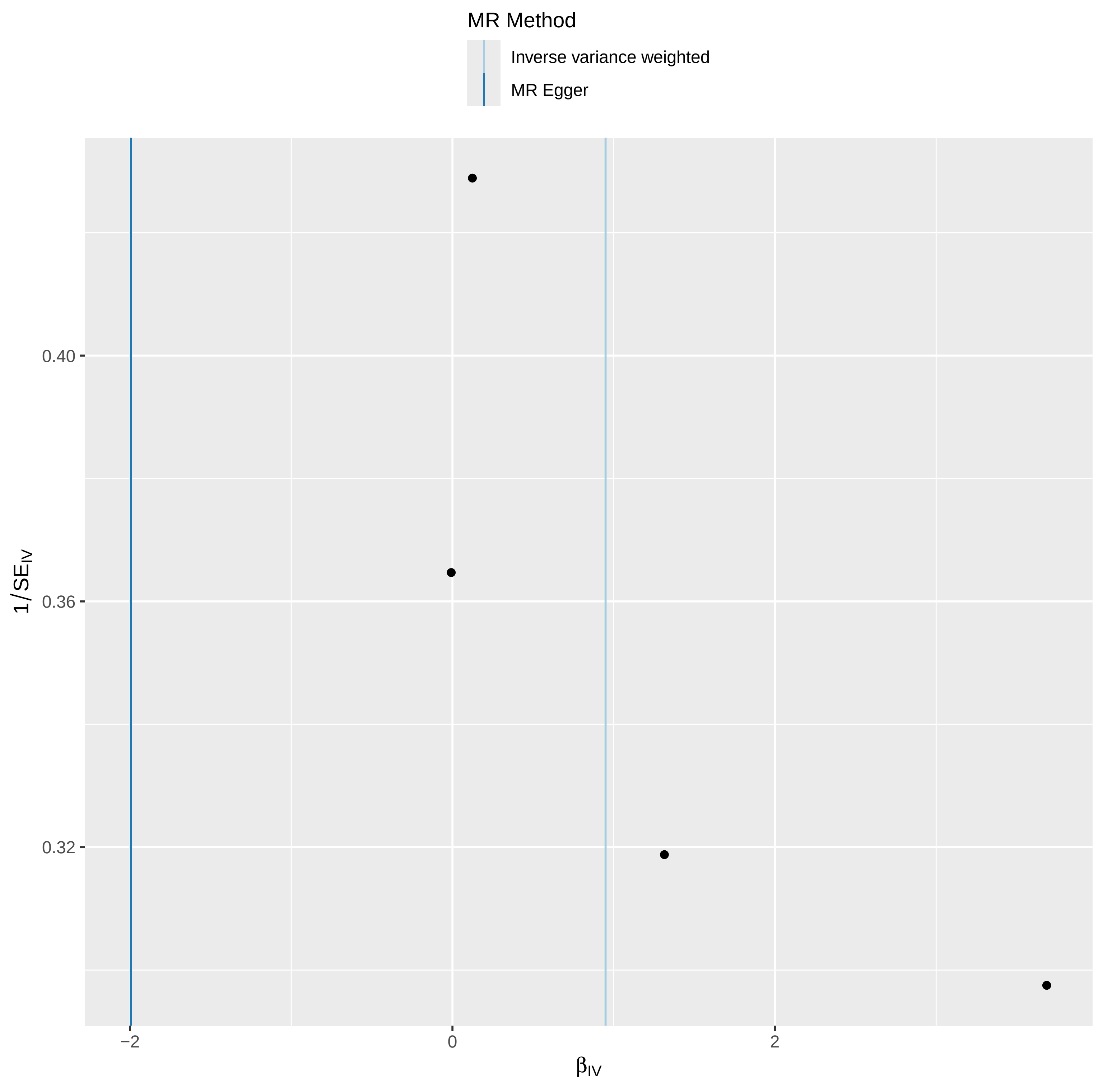

### funnelplot.pdf

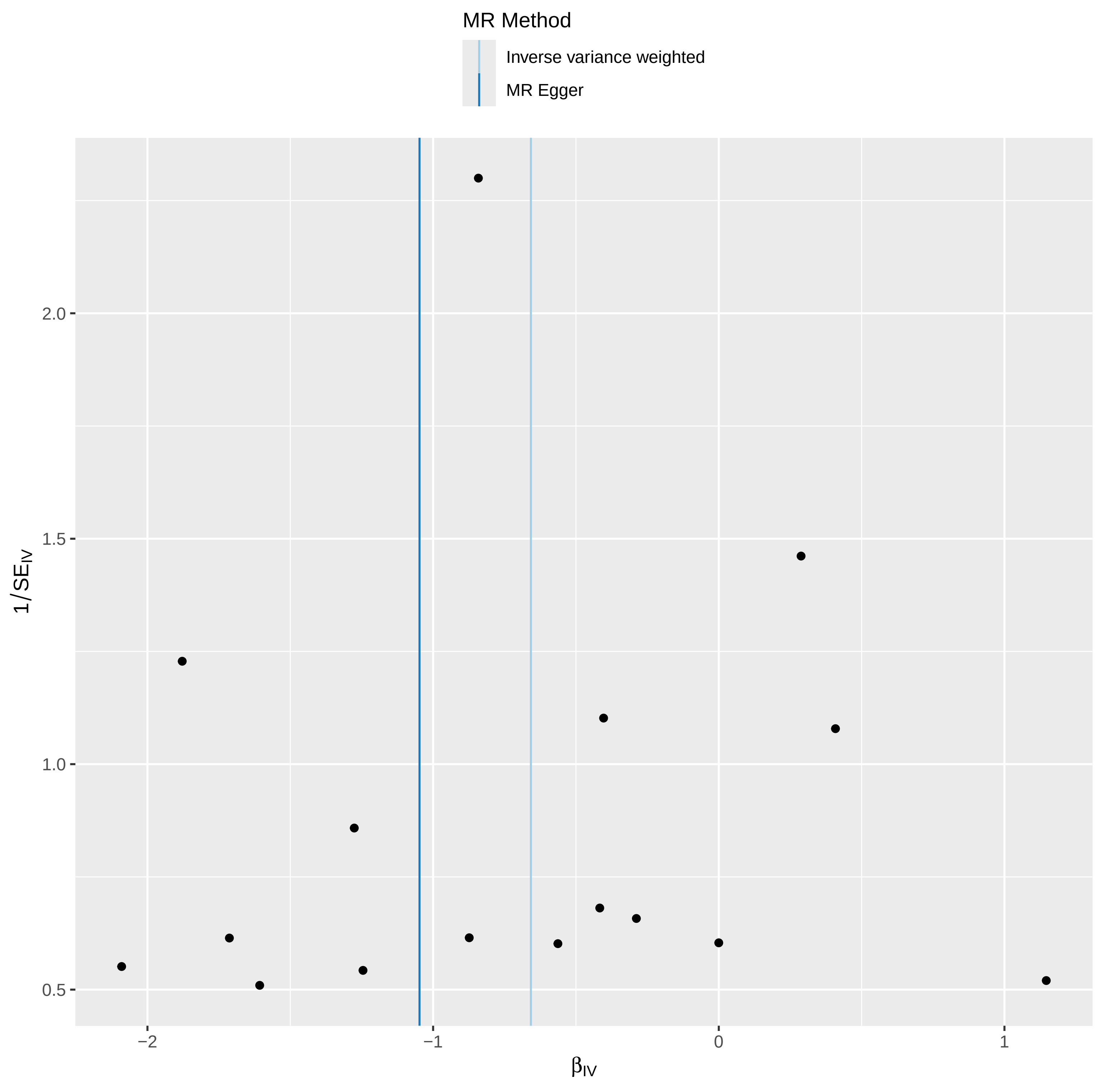

### funnelplot.pdf

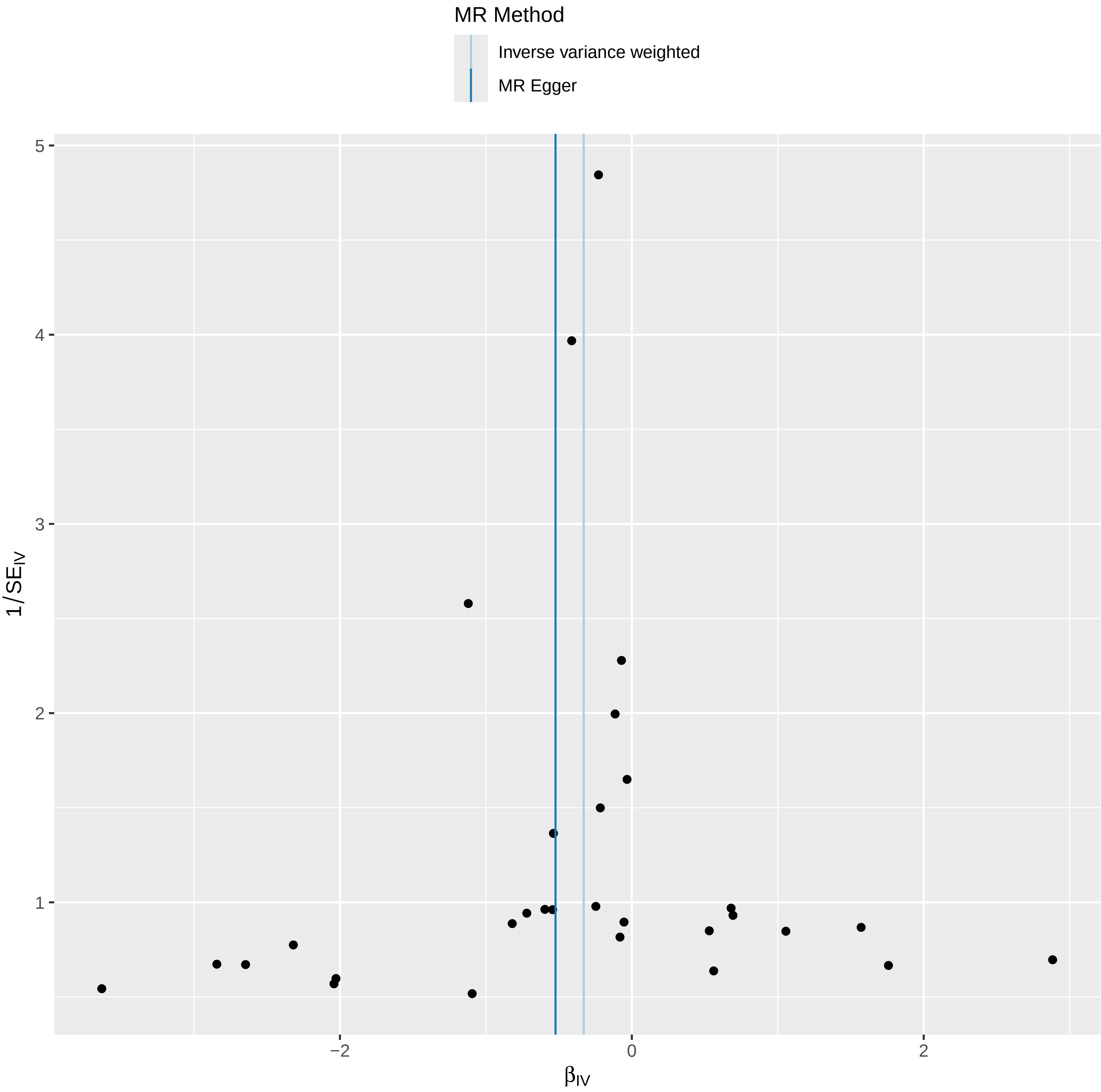

### funnelplot.pdf

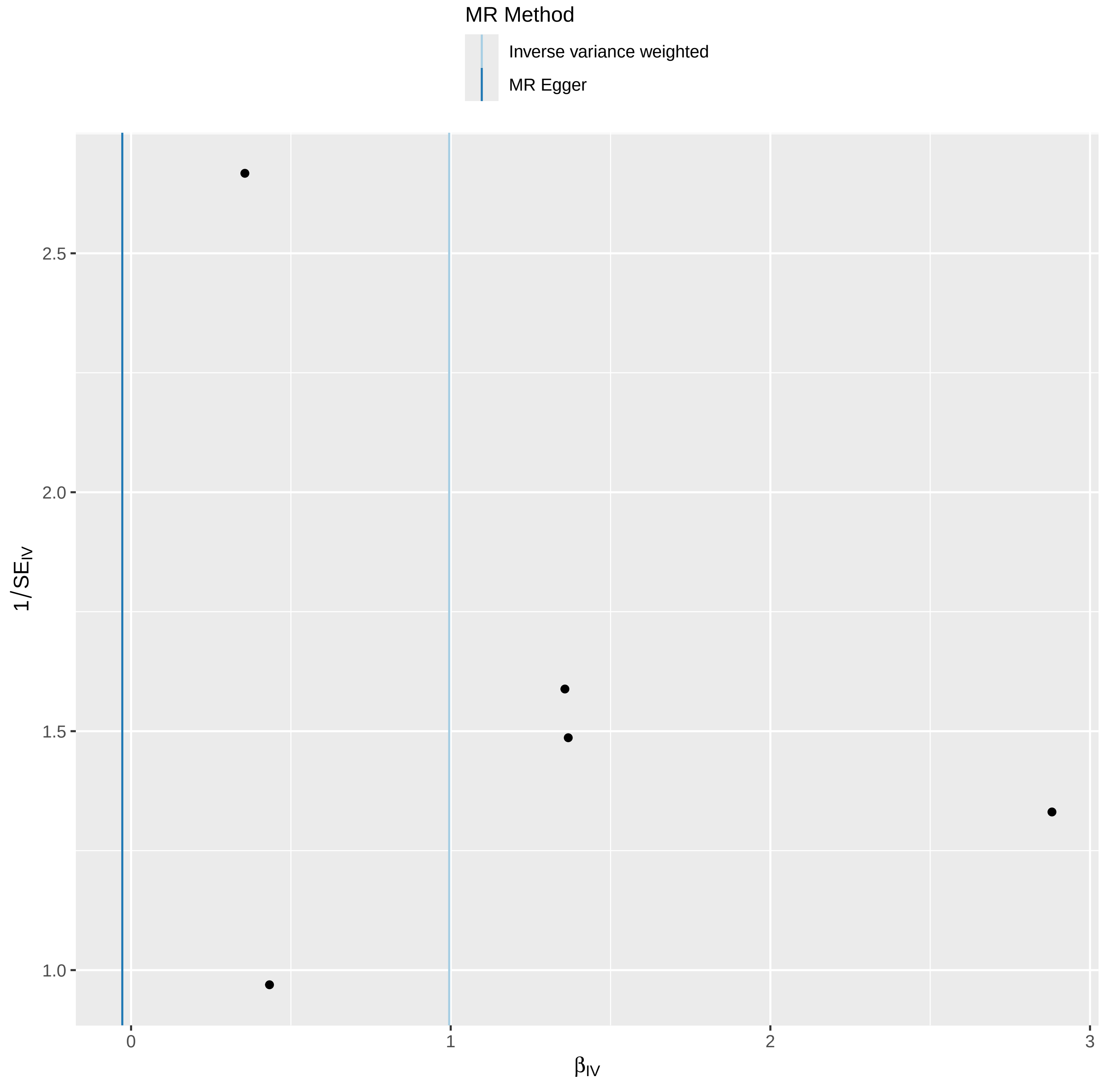

### funnelplot.pdf

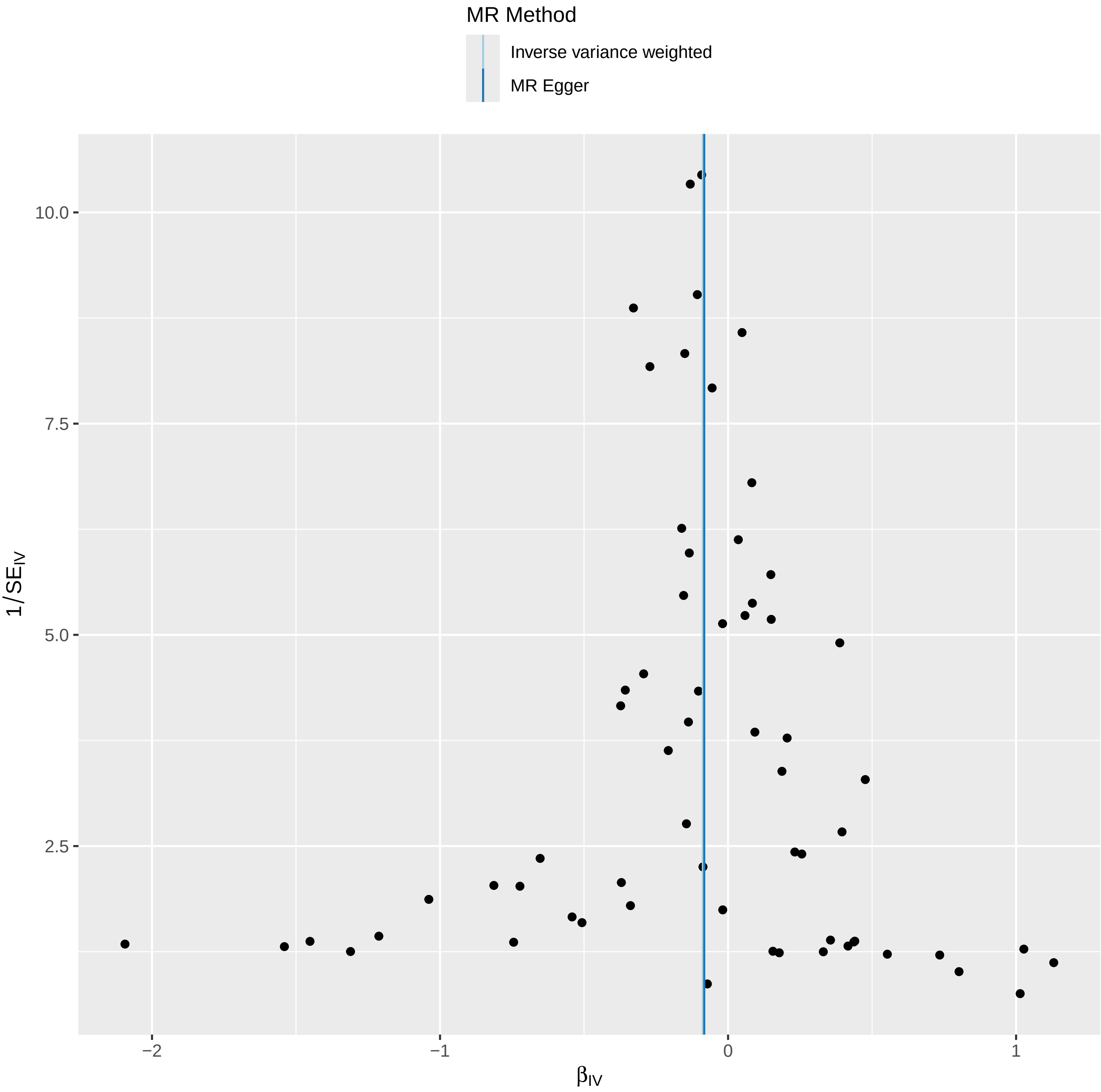

### funnelplot.pdf

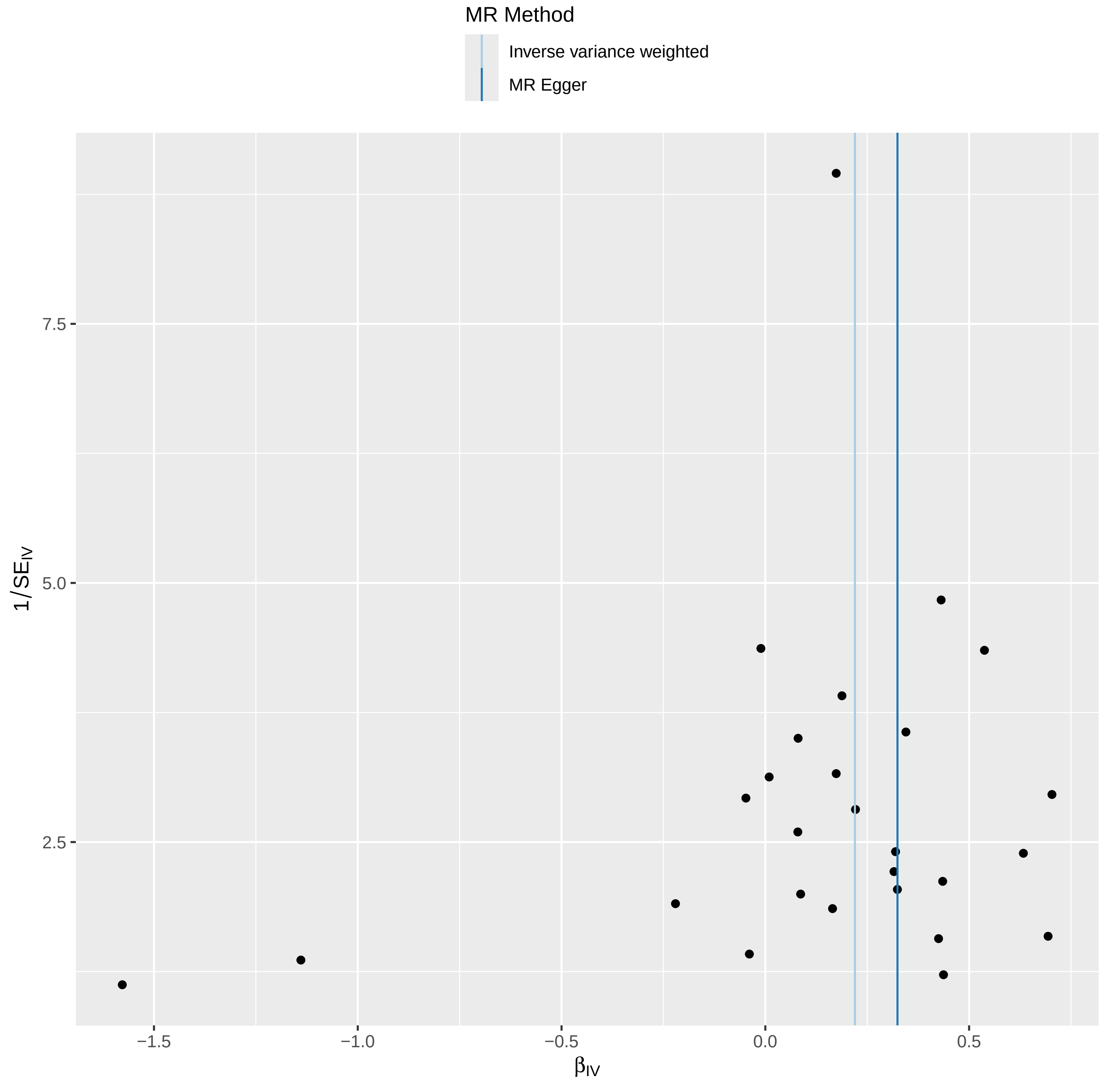

### funnelplot.pdf

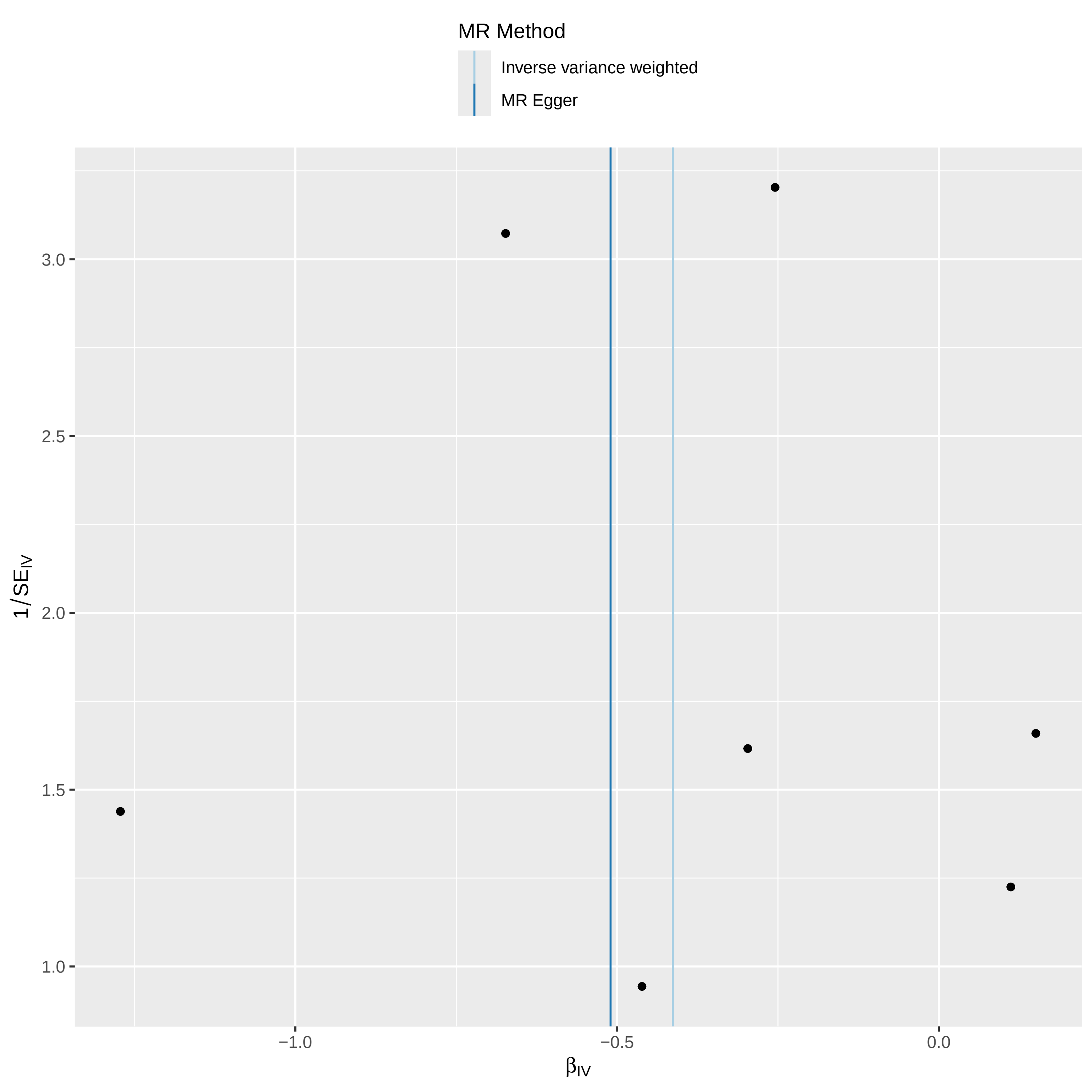

### funnelplot.pdf

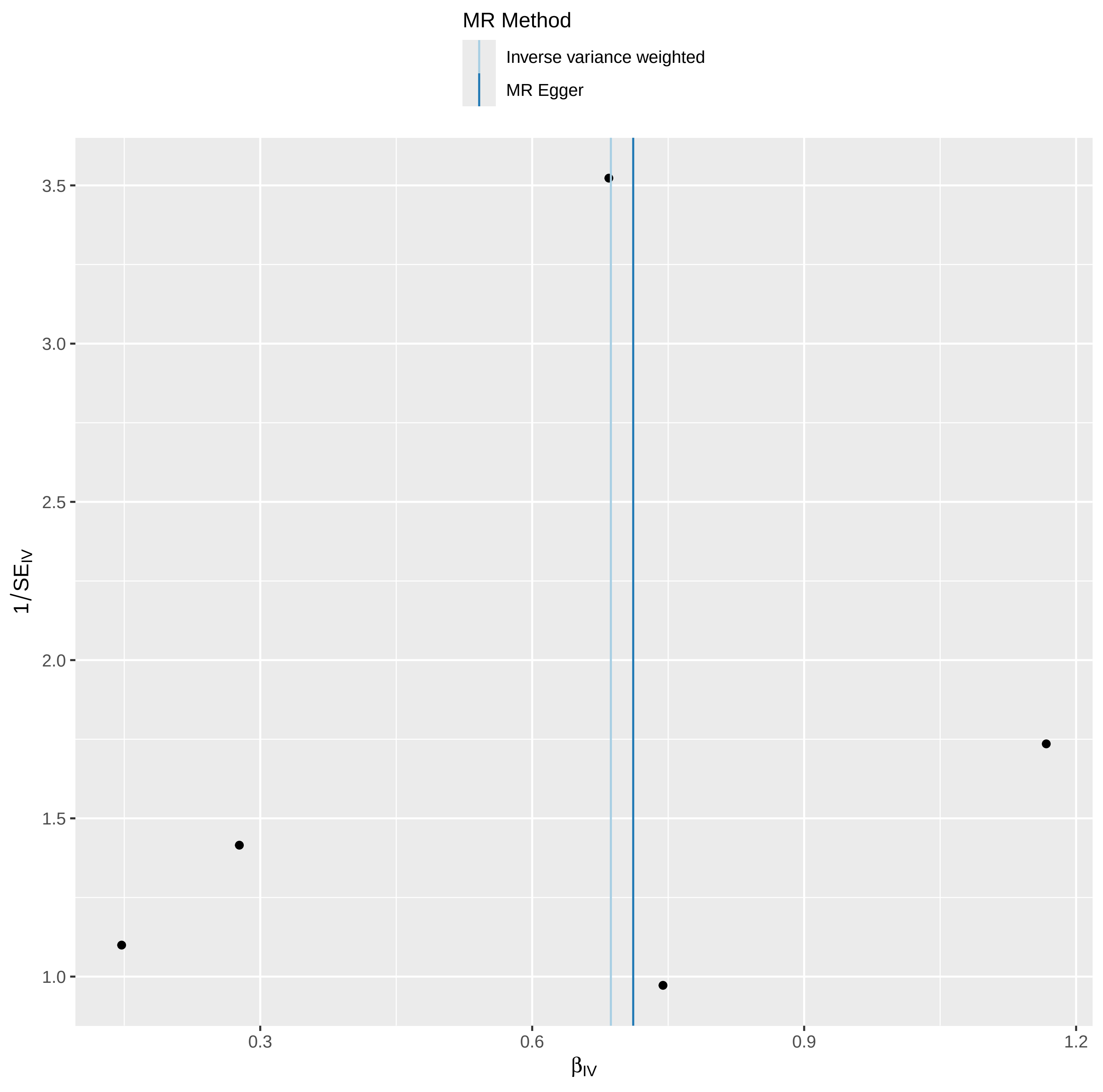

### funnelplot.pdf

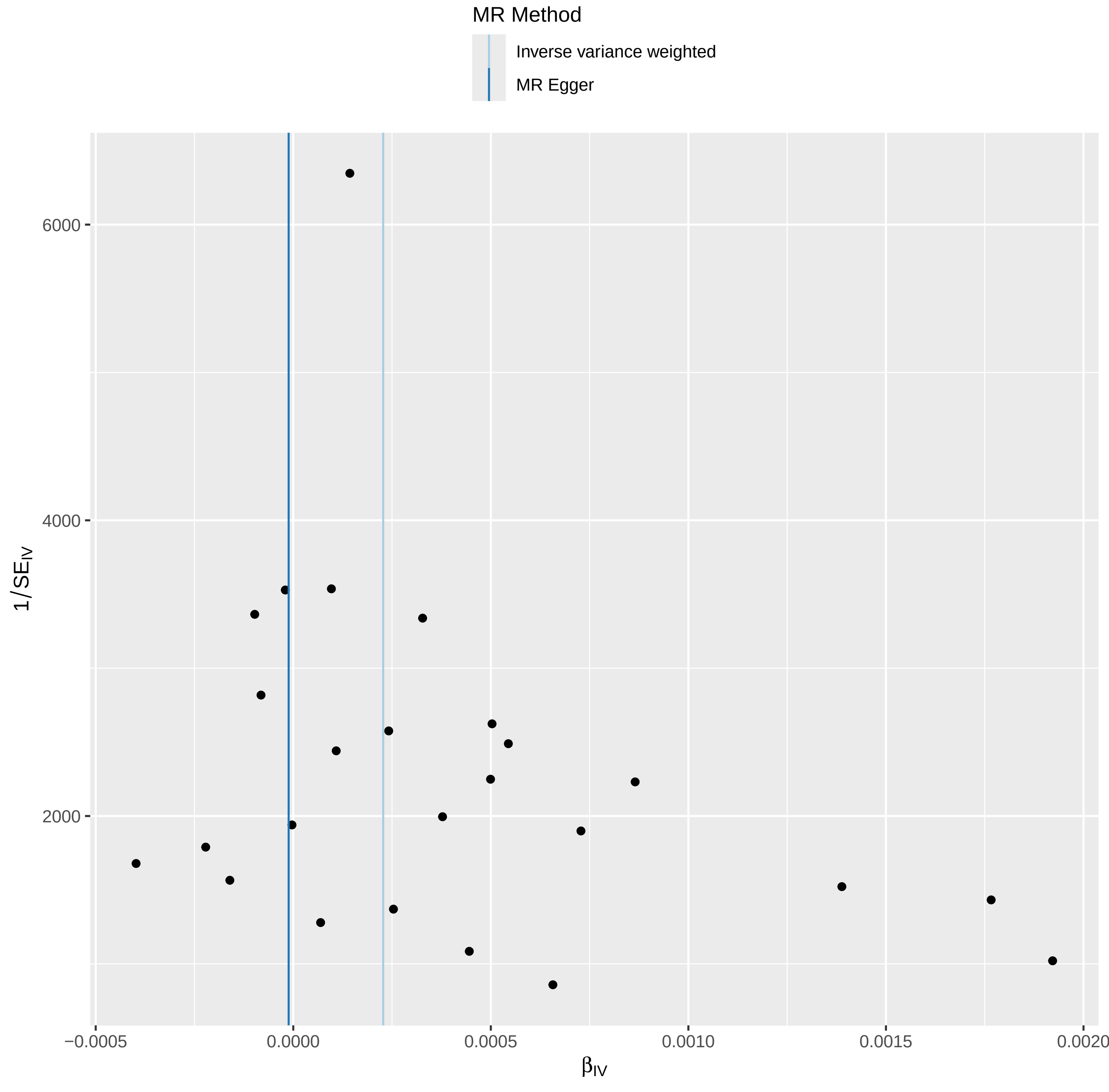

### funnelplot.pdf

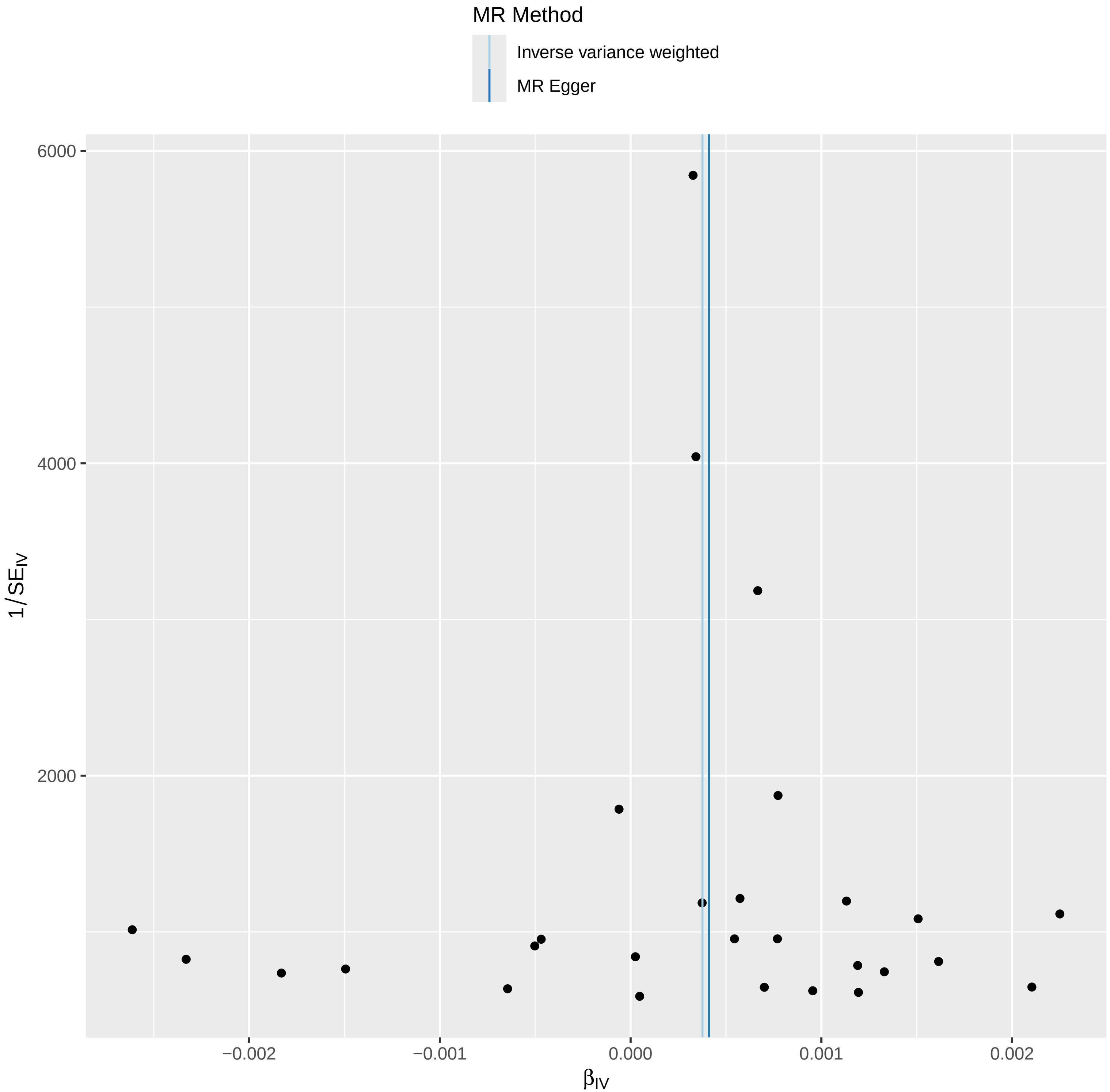

### funnelplot.pdf

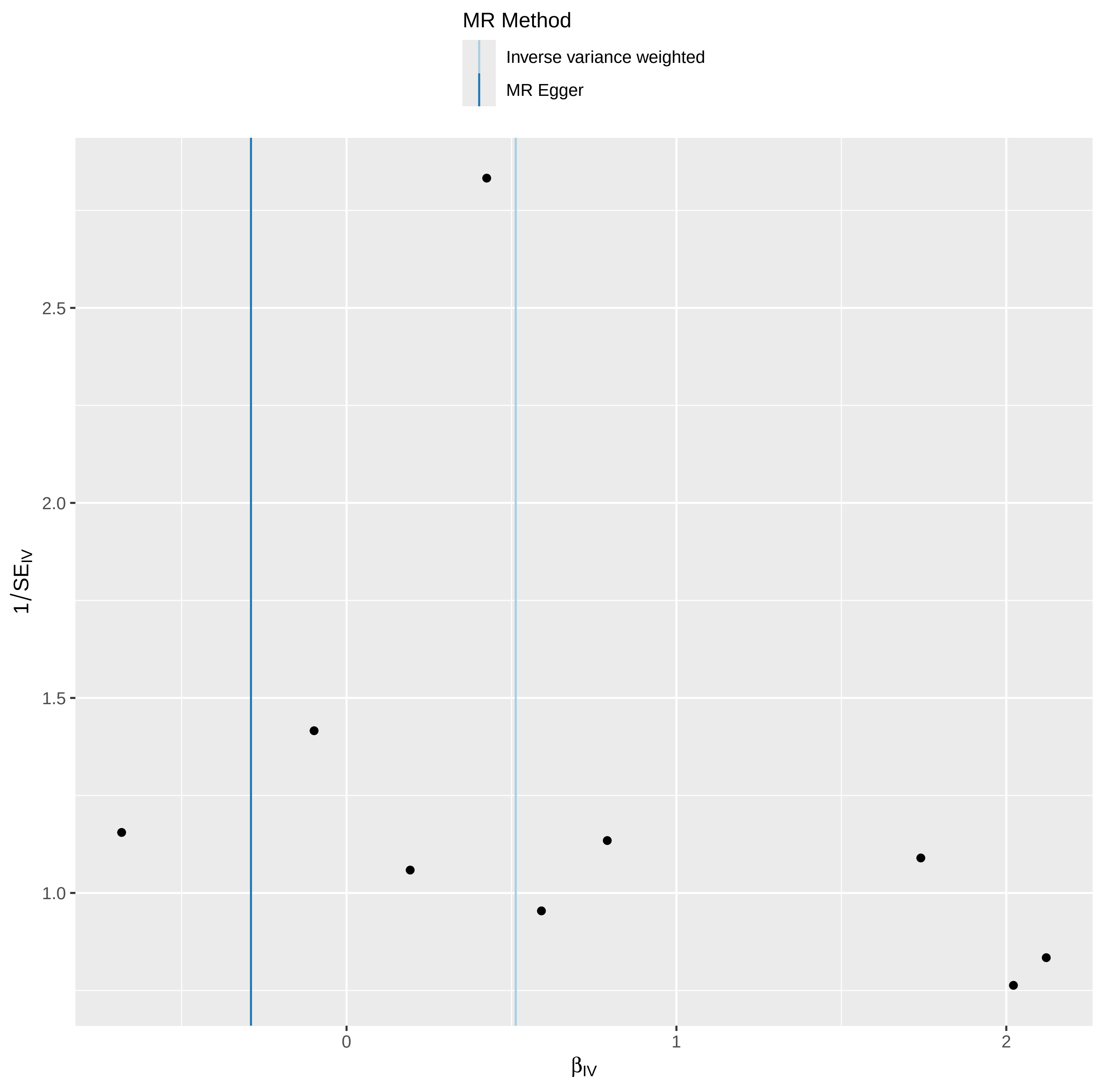

### funnelplot.pdf

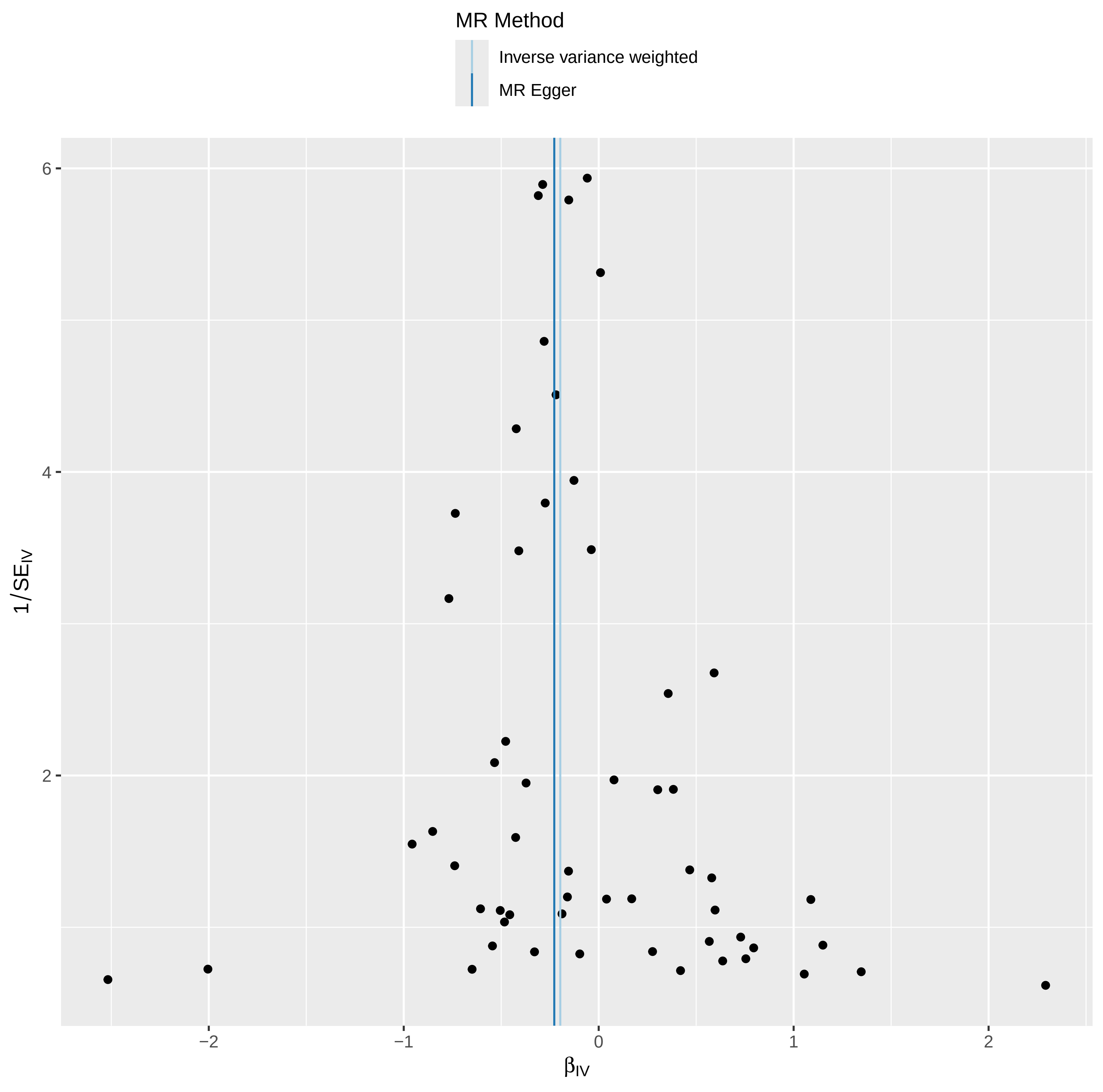

### funnelplot.pdf

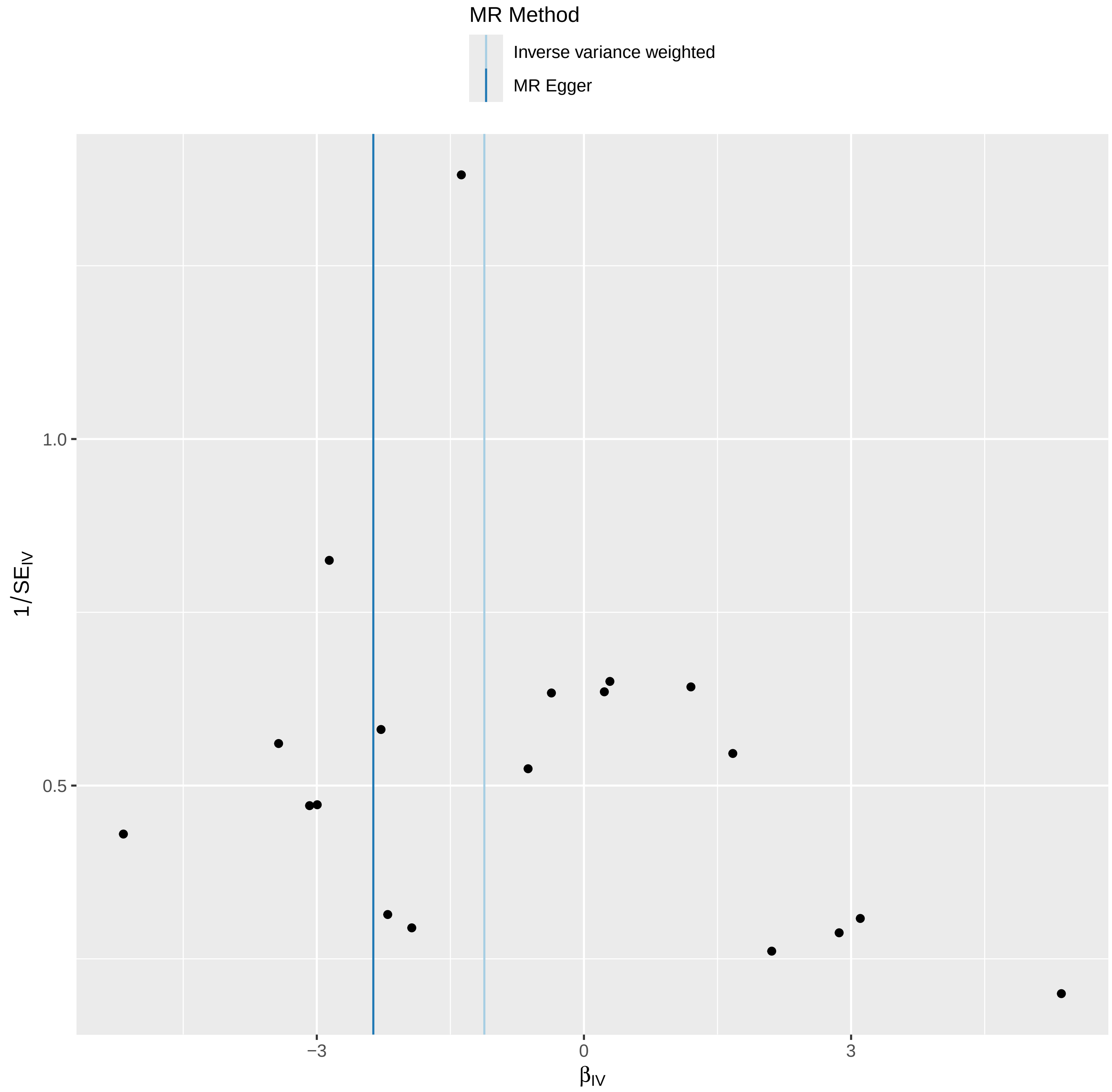

### funnelplot.pdf

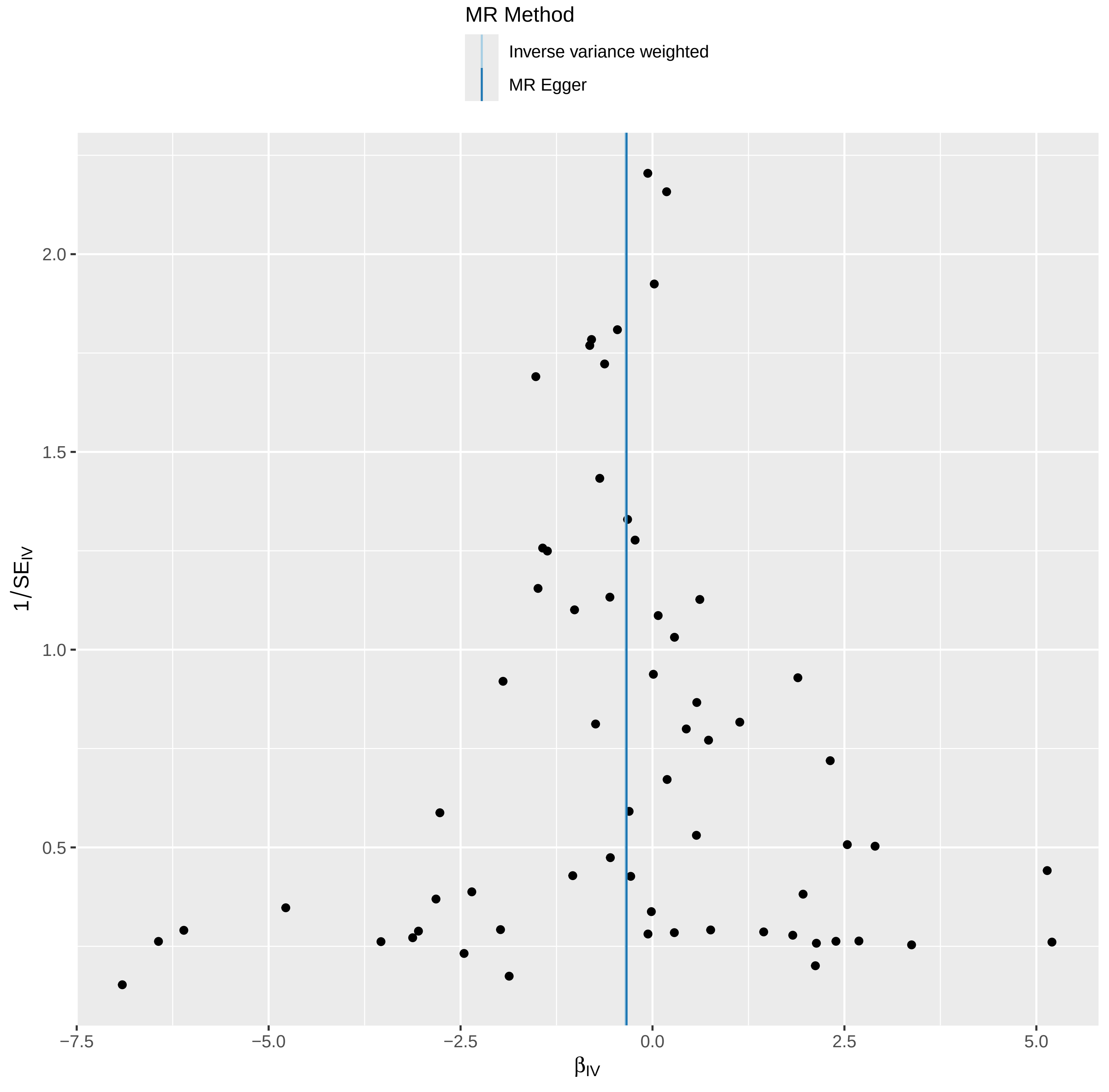

### GO_KEGG_PPI network.jpg

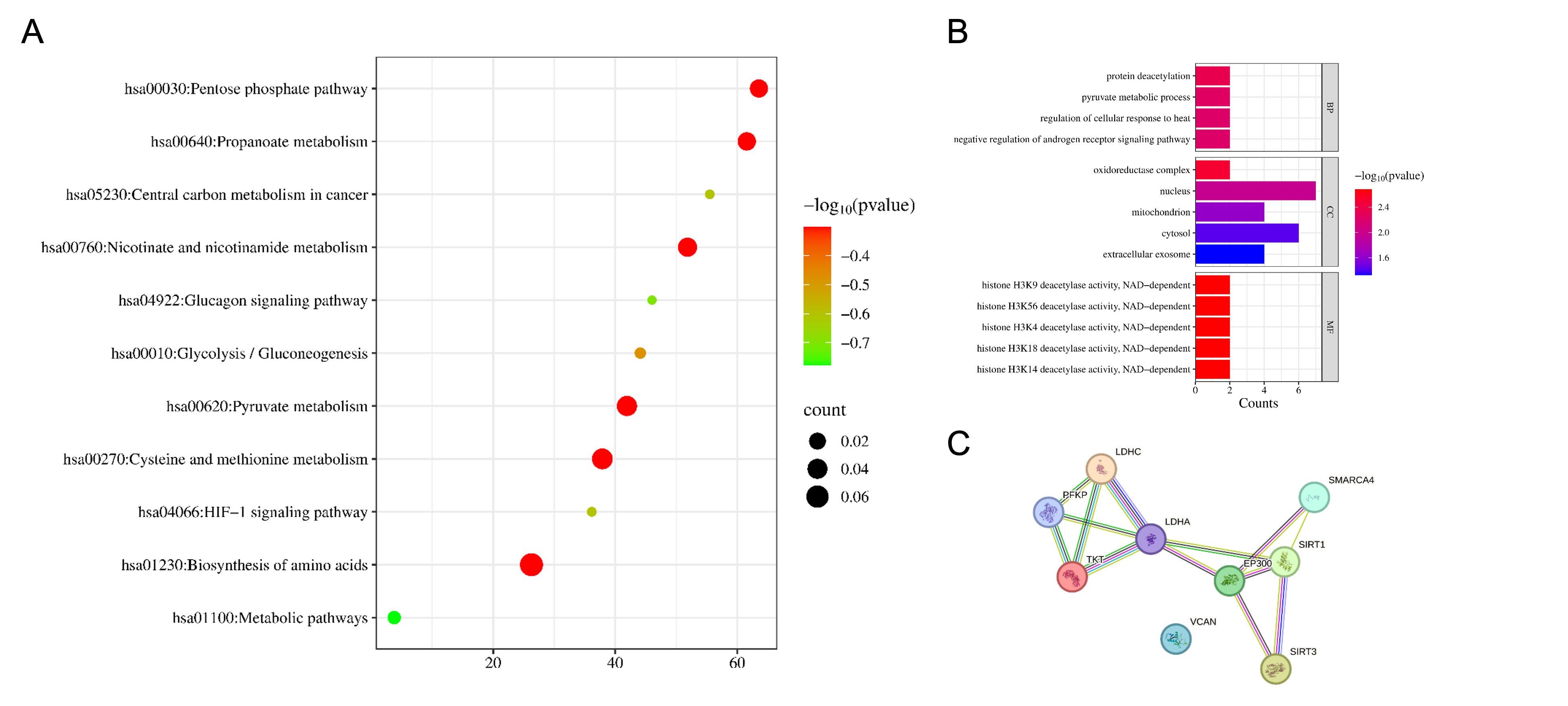

### ML.jpg

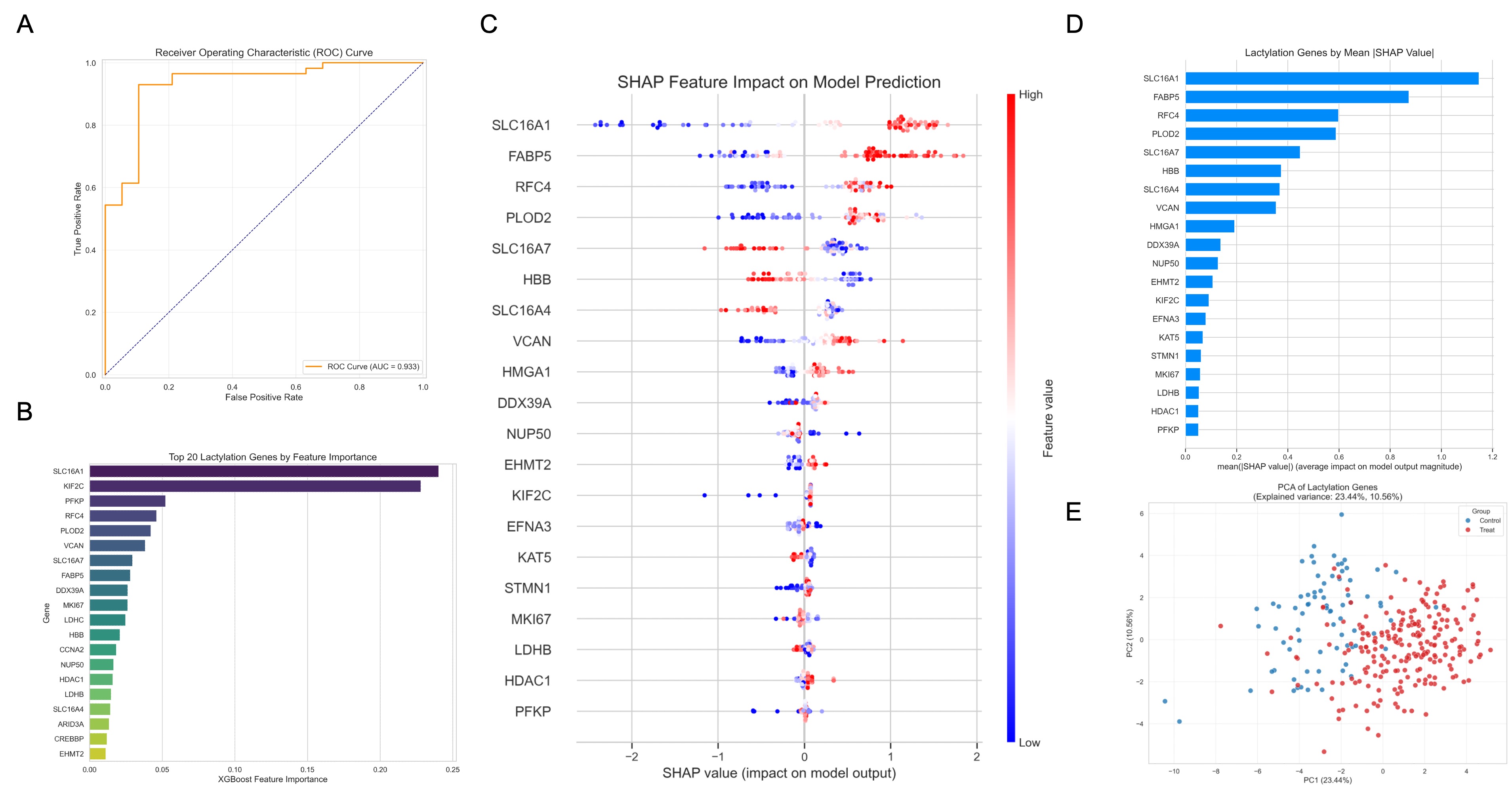

### pQTL_lactylation.pdf

#### Overlap of pQTL and Lactylation Genes

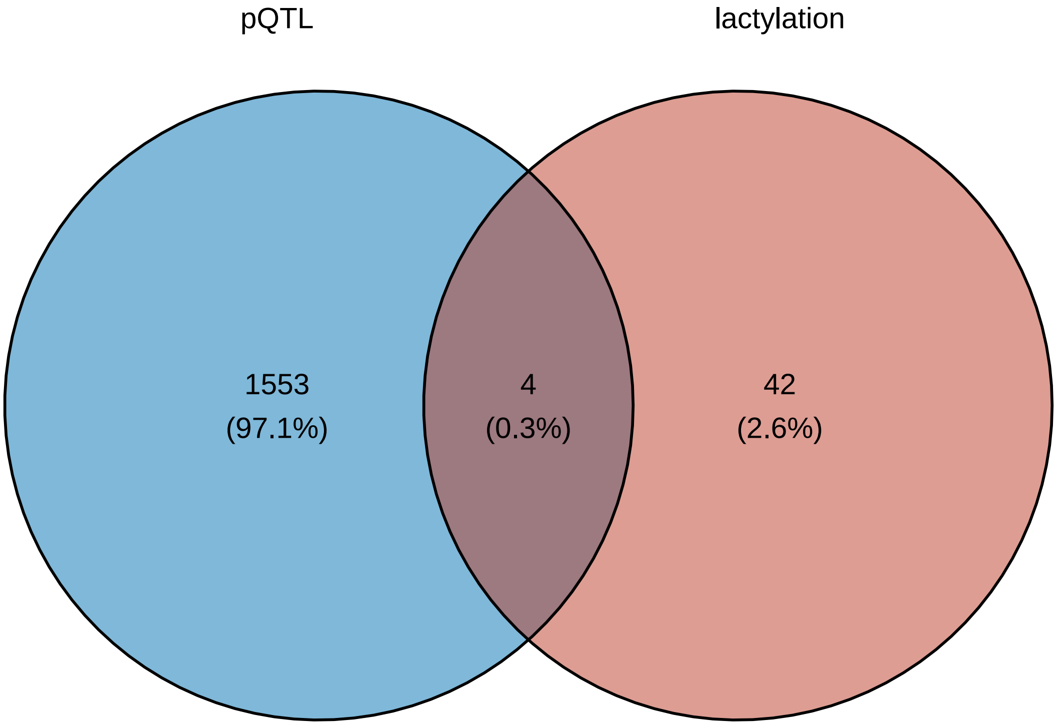

### scatter.pdf

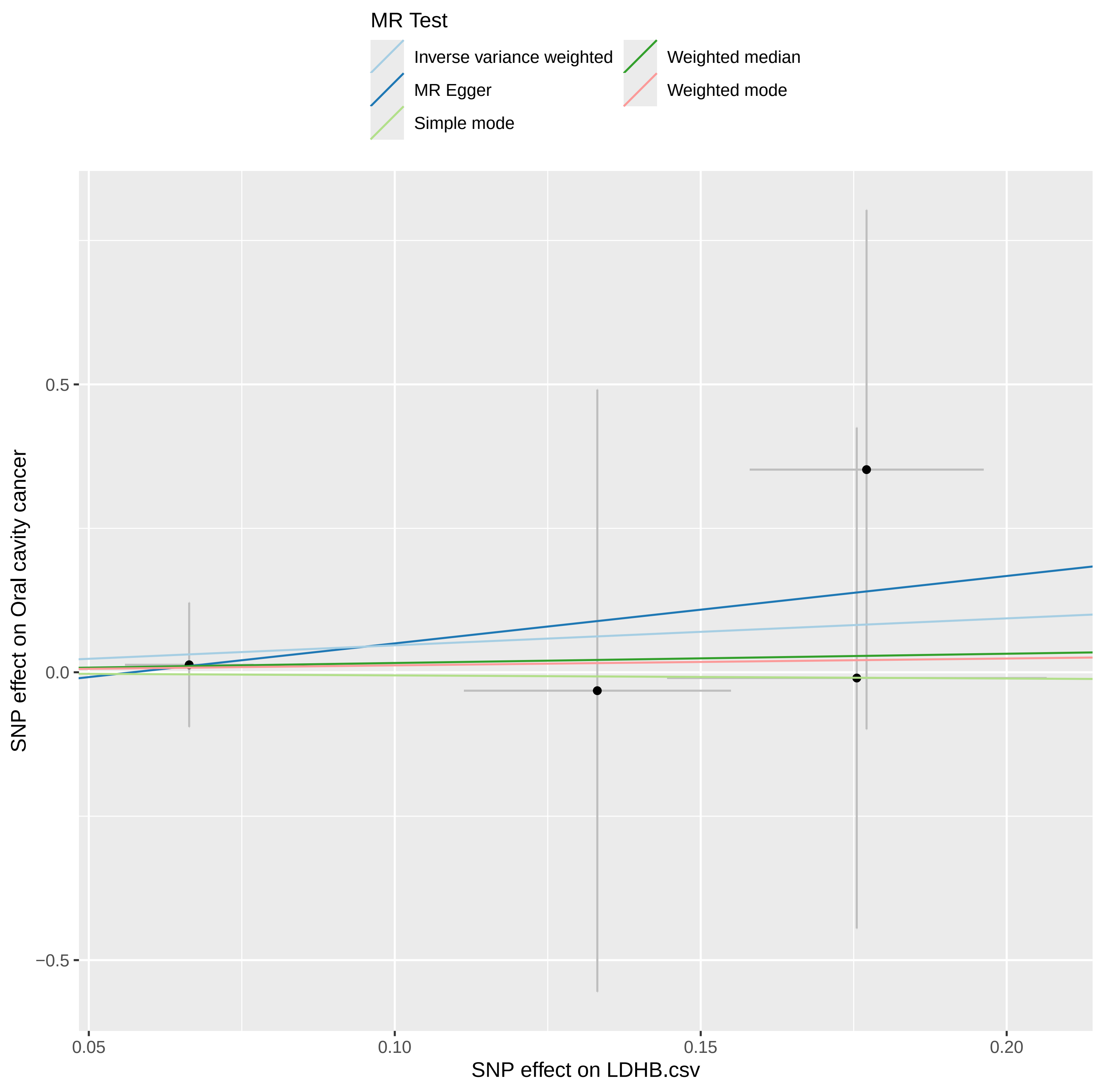

### scatter.pdf

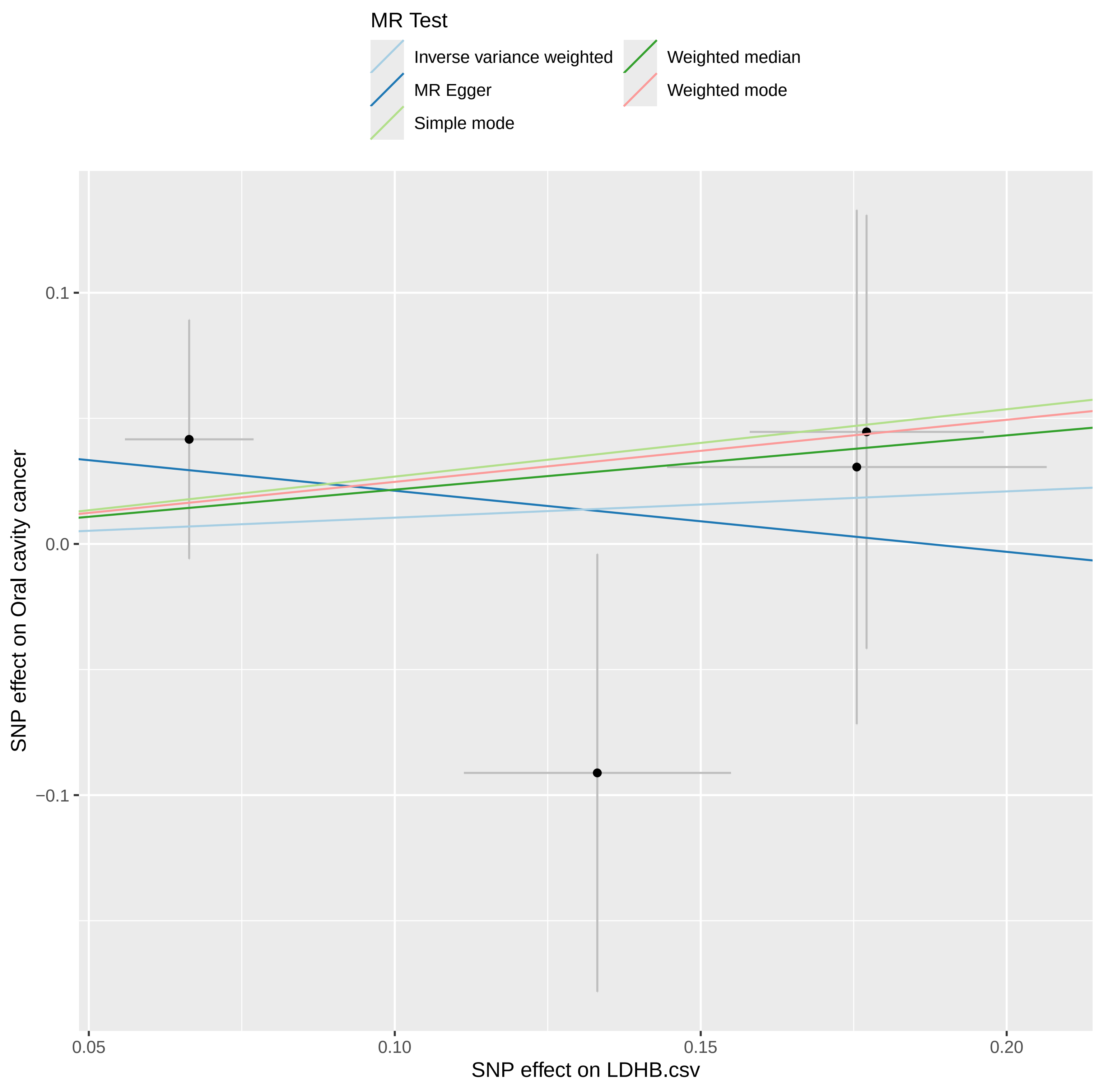

### scatter.pdf

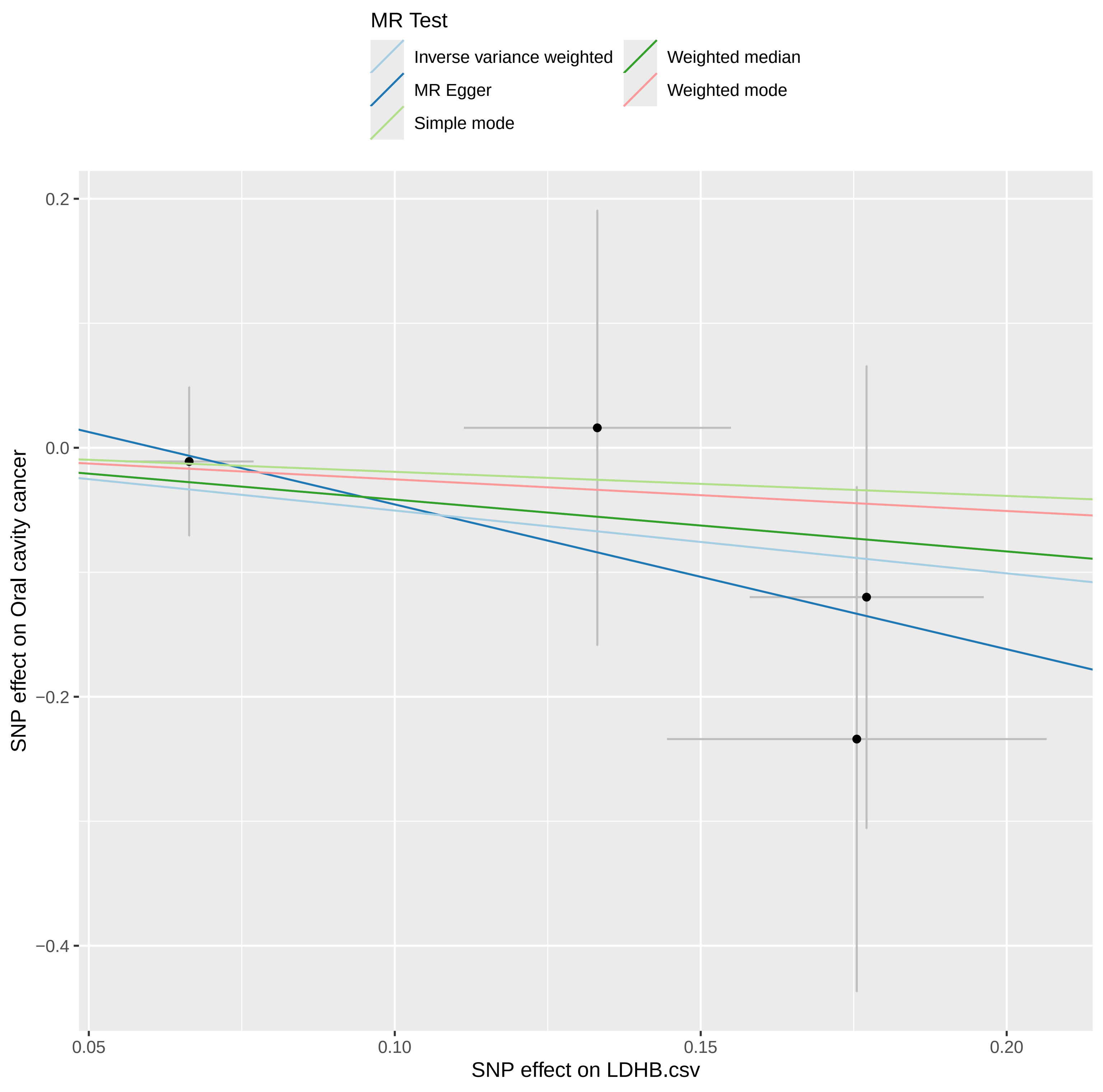

### scatter.pdf

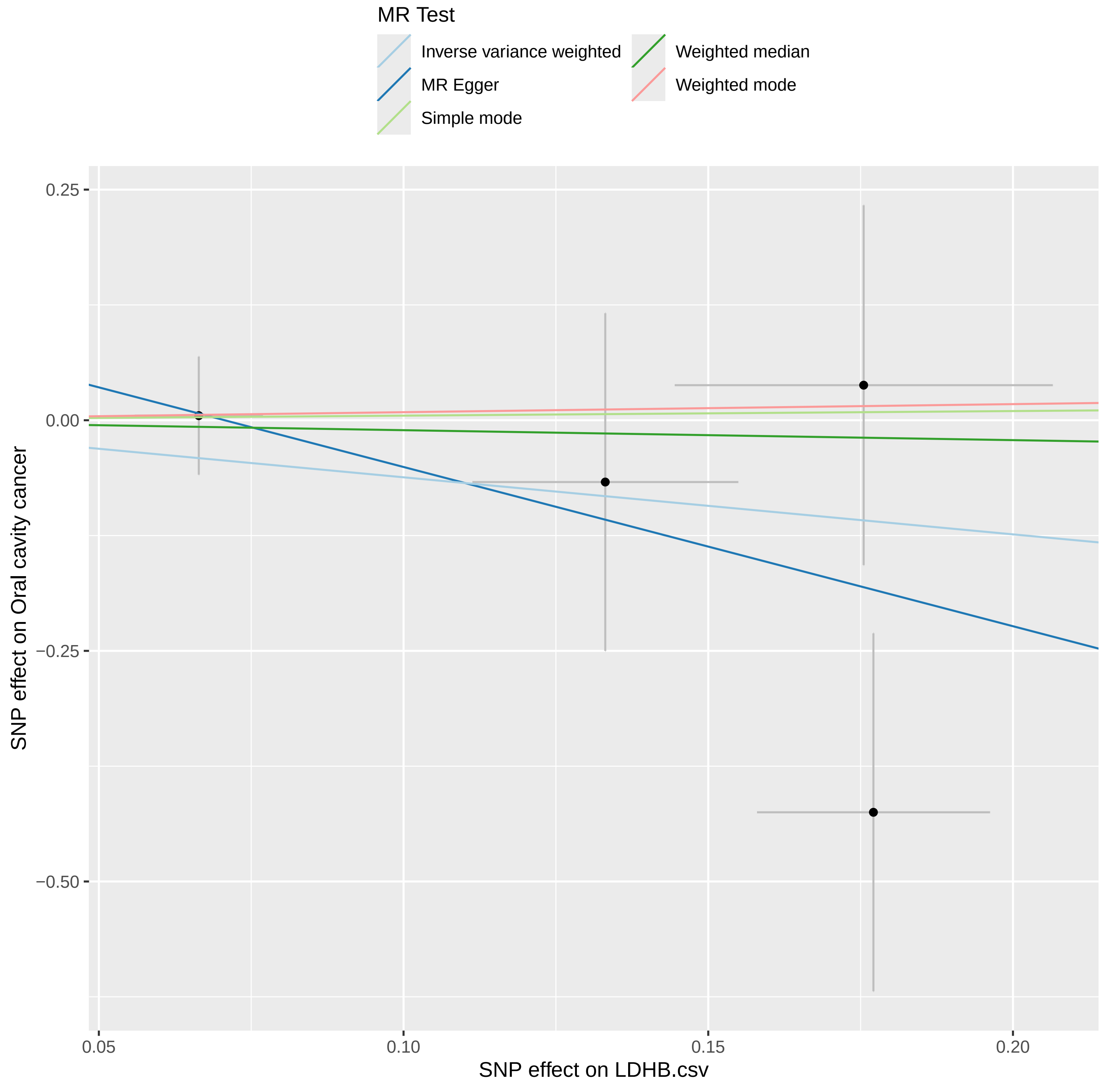

### scatter.pdf

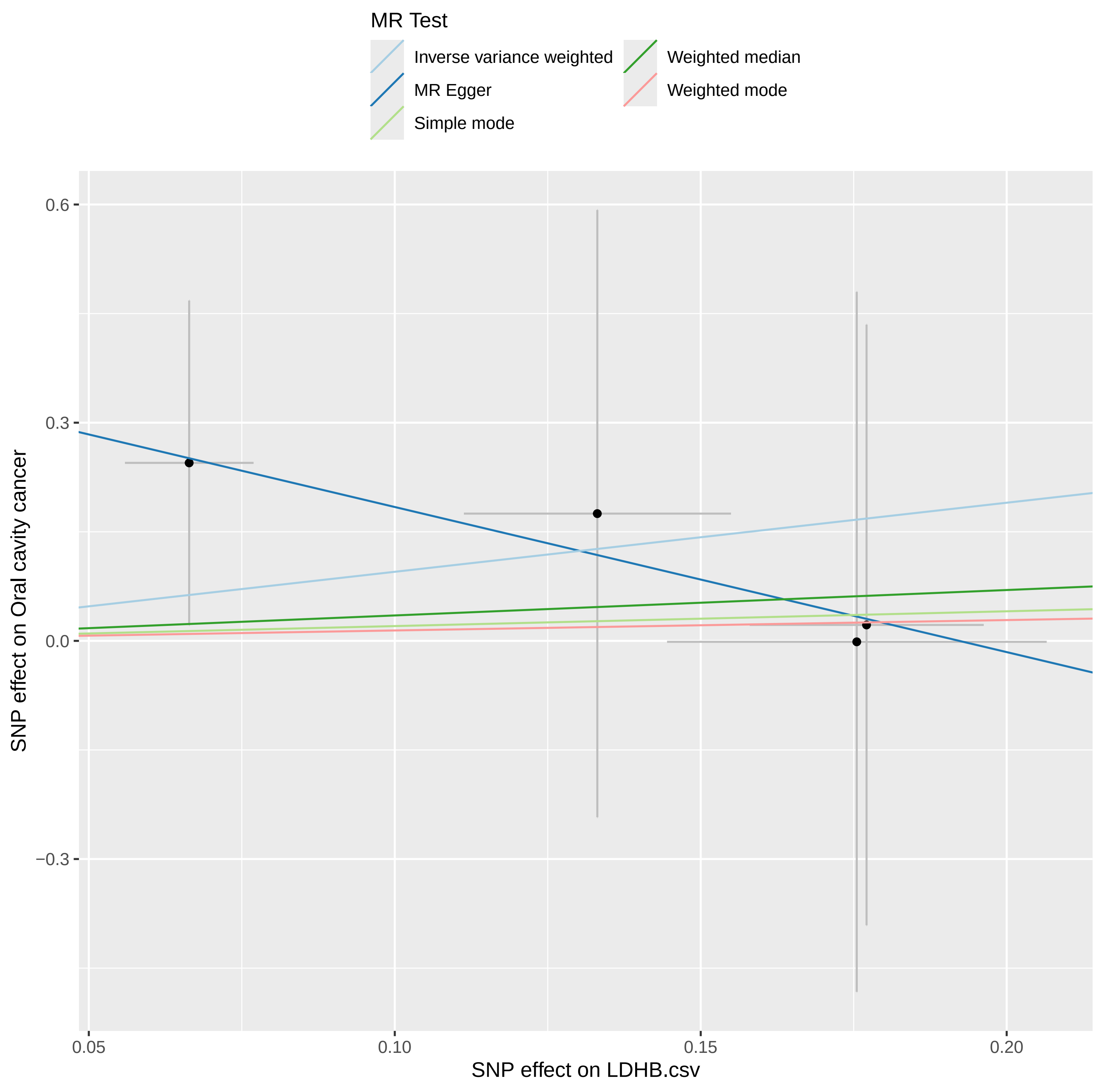

### scatter.pdf

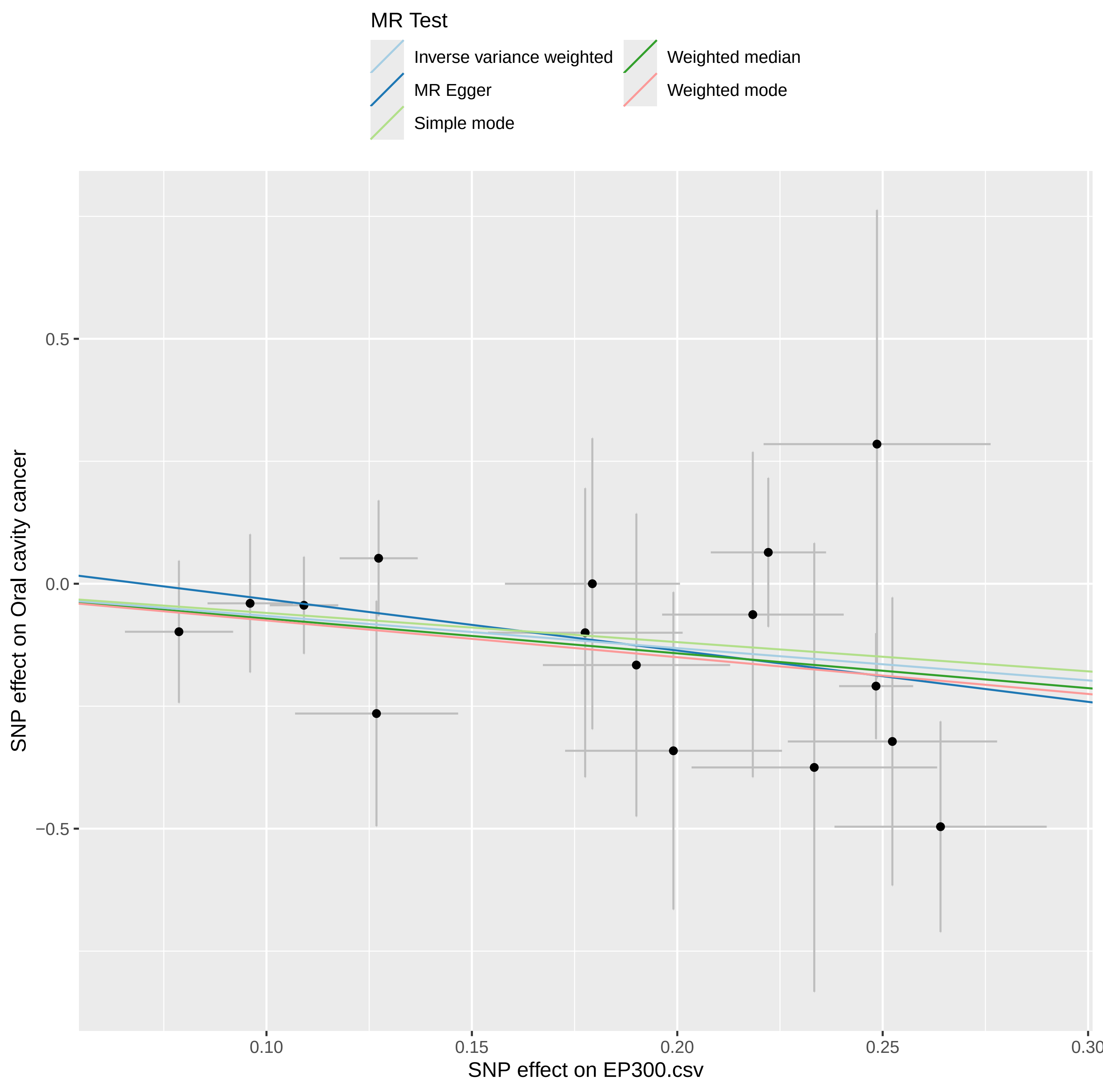

### scatter.pdf

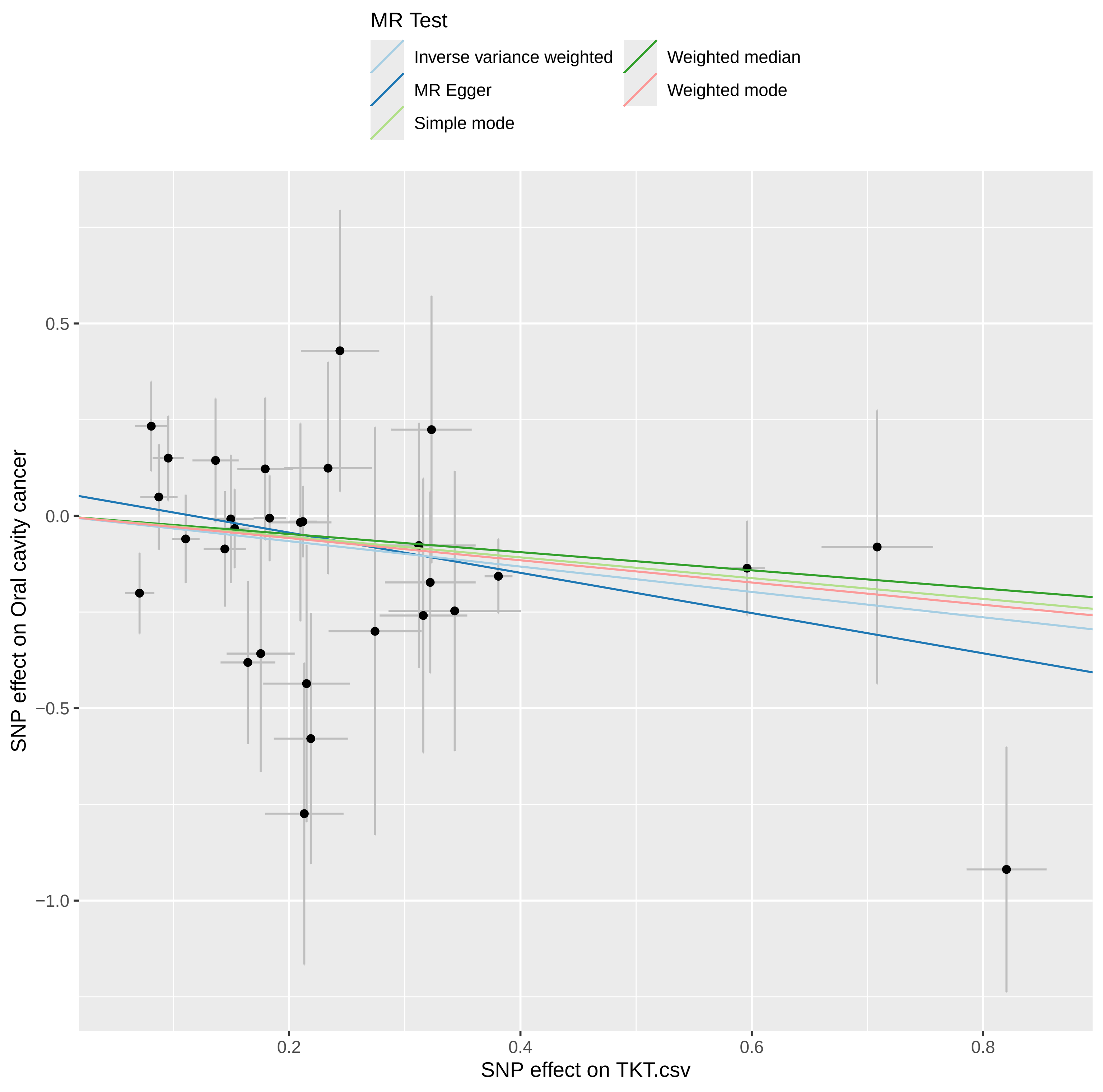

### scatter.pdf

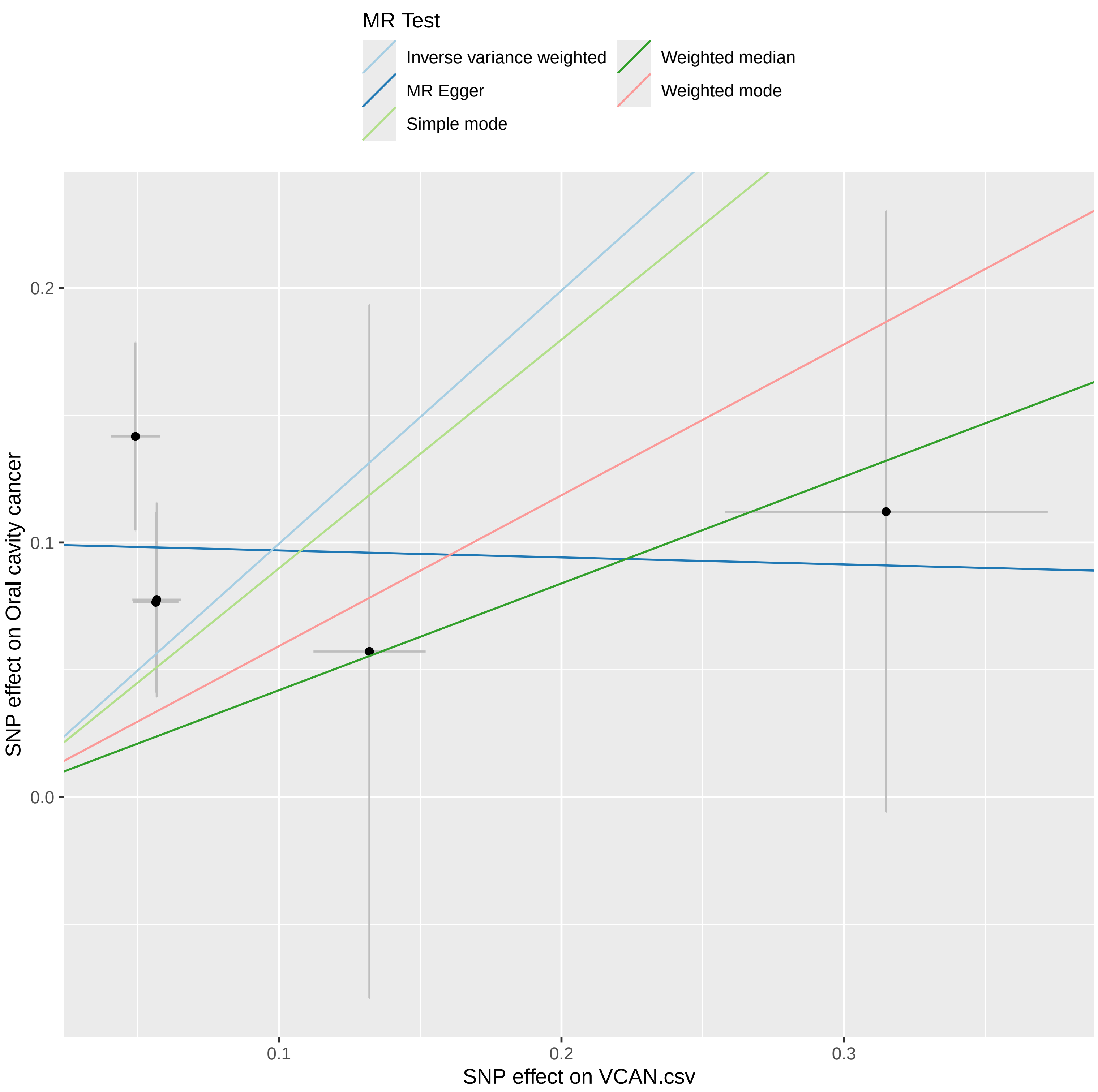

### scatter.pdf

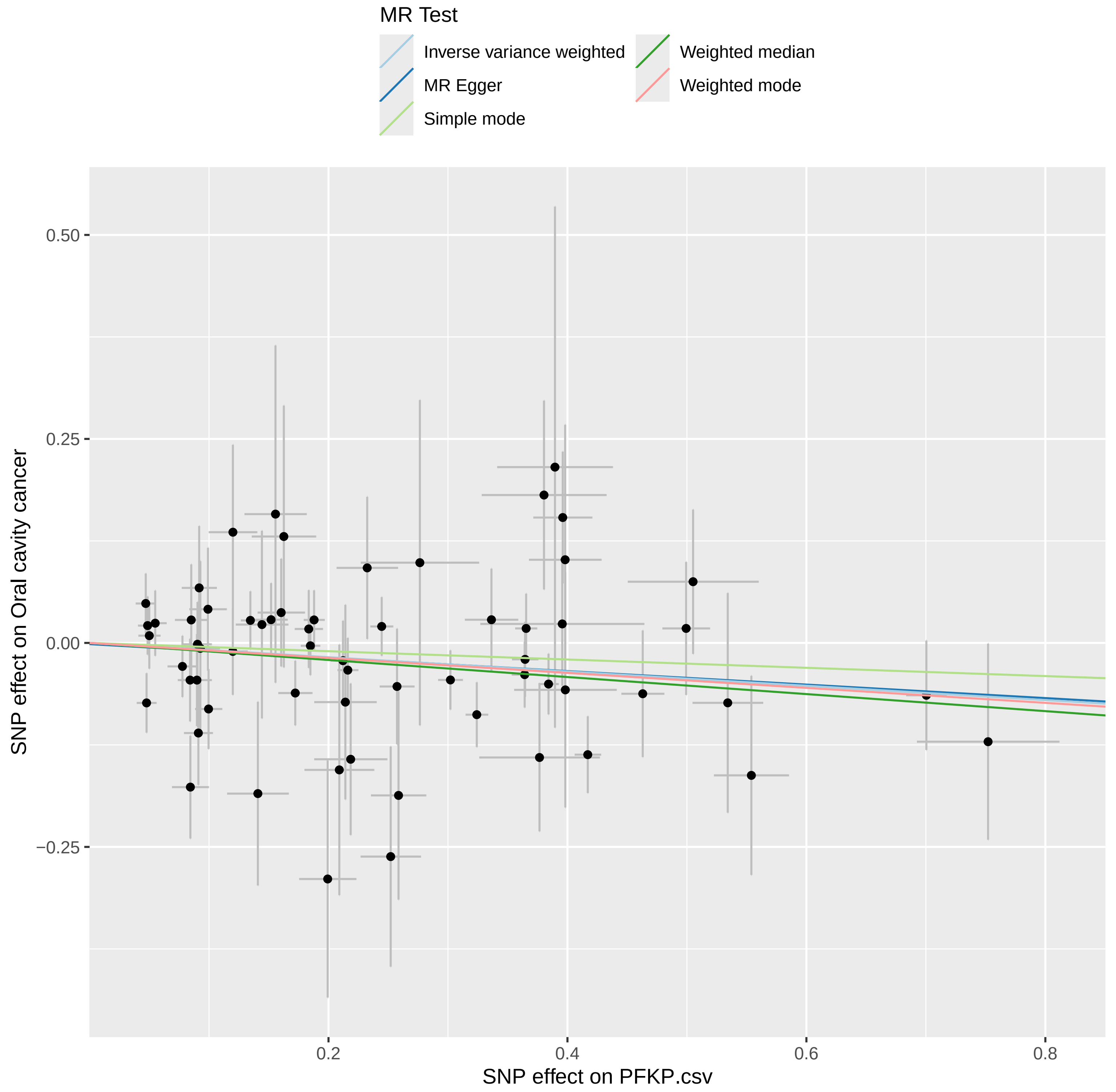

### scatter.pdf

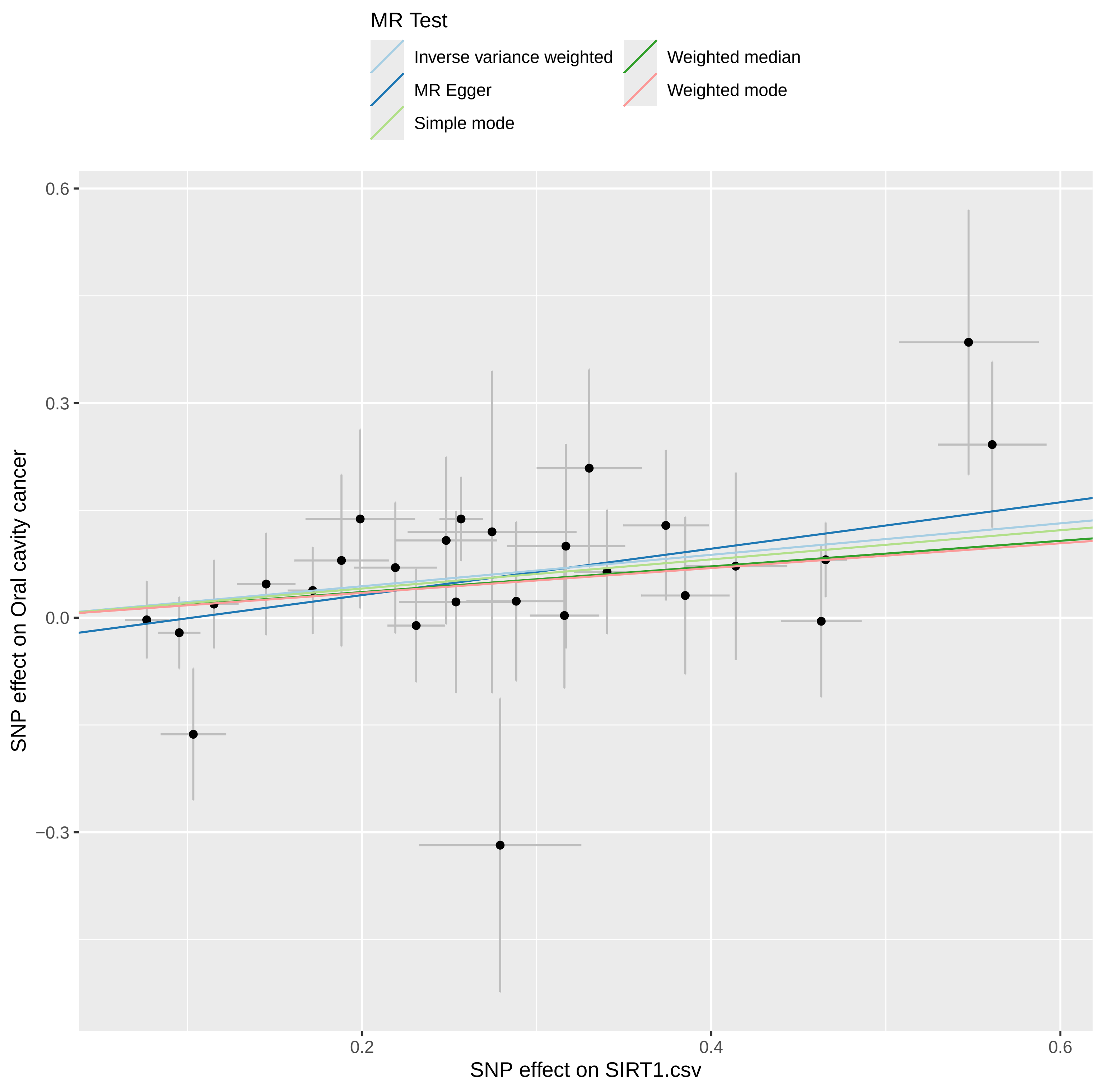

### scatter.pdf

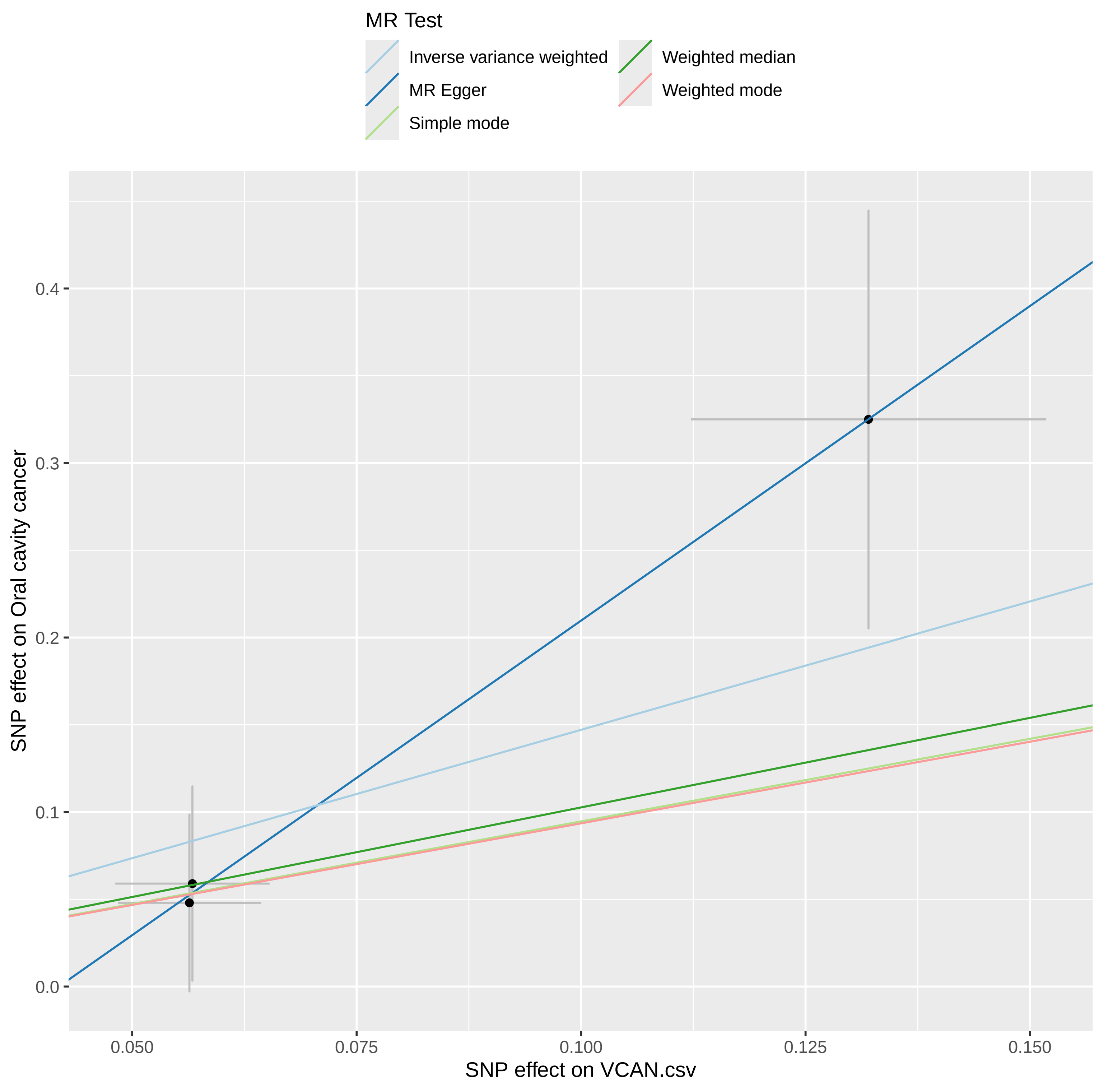

### scatter.pdf

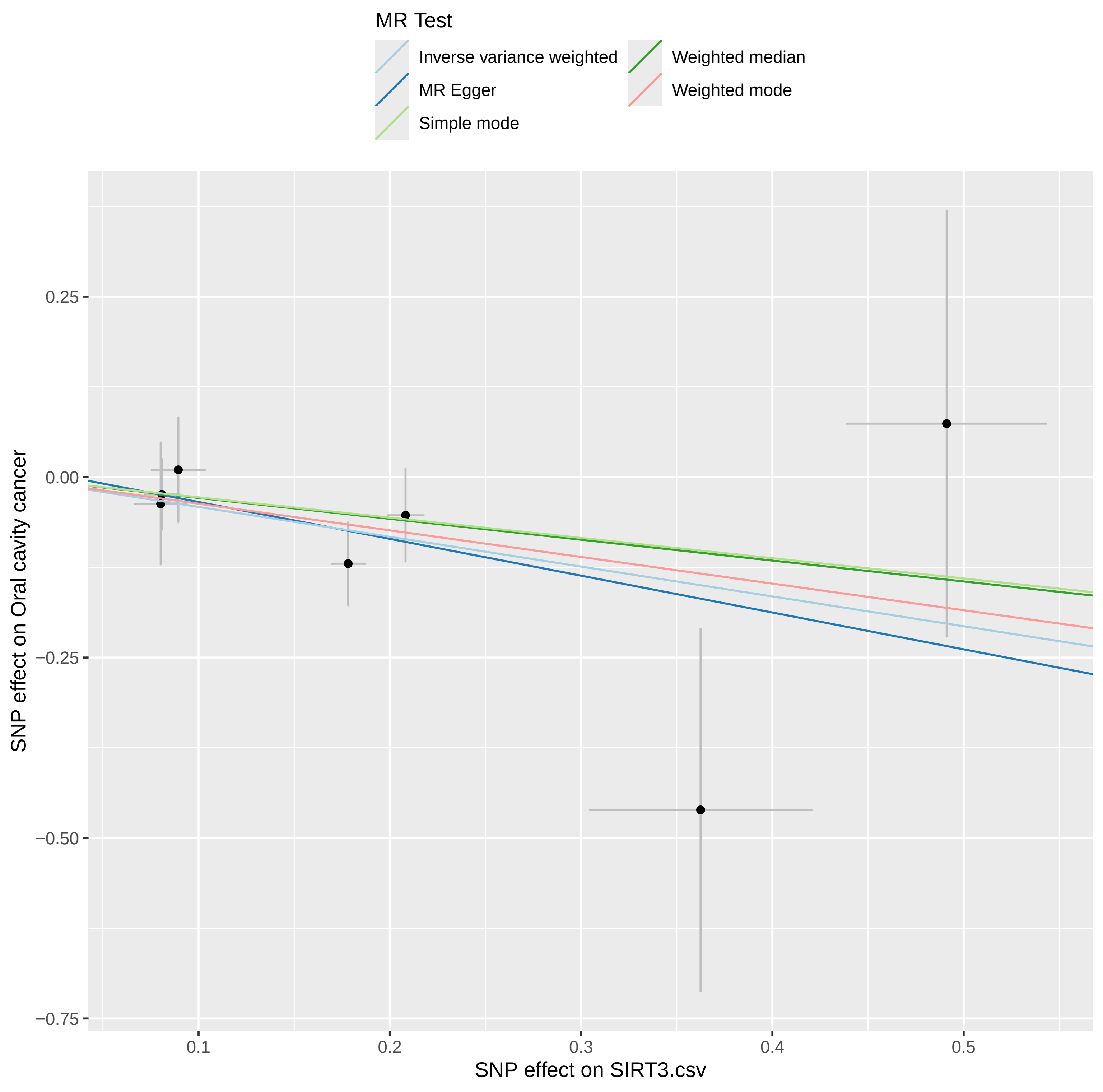

### scatter.pdf

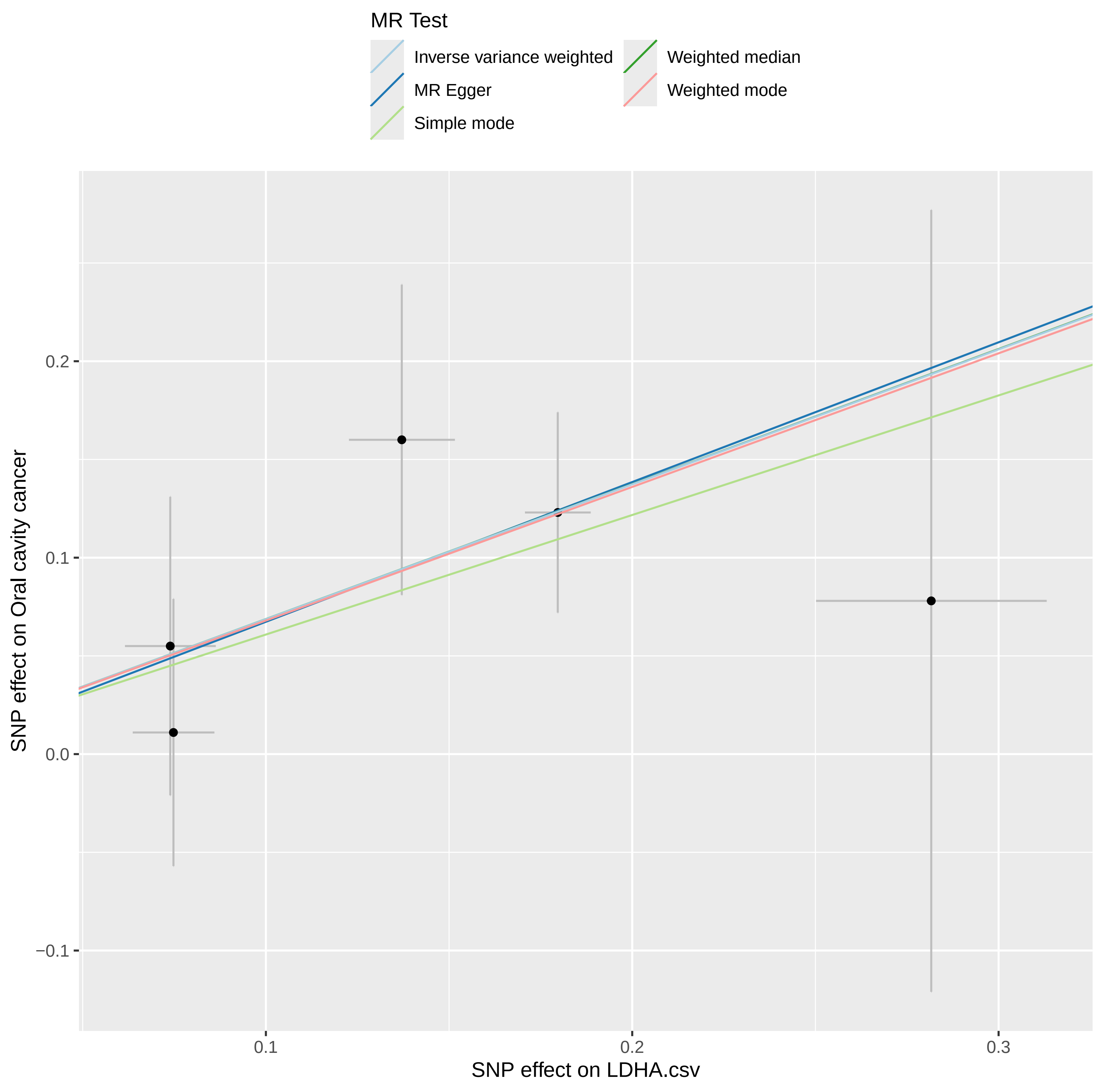

### scatter.pdf

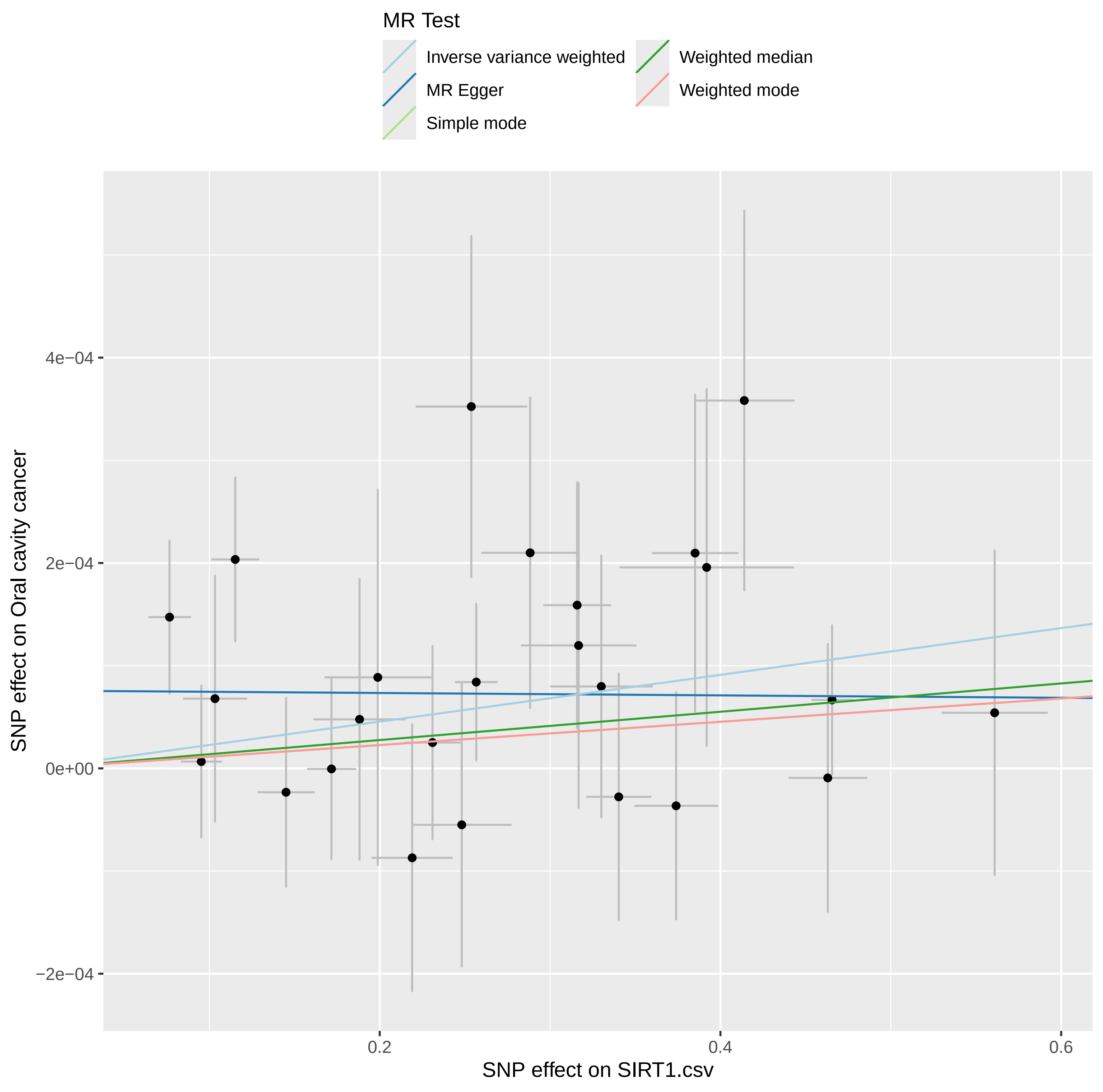

### scatter.pdf

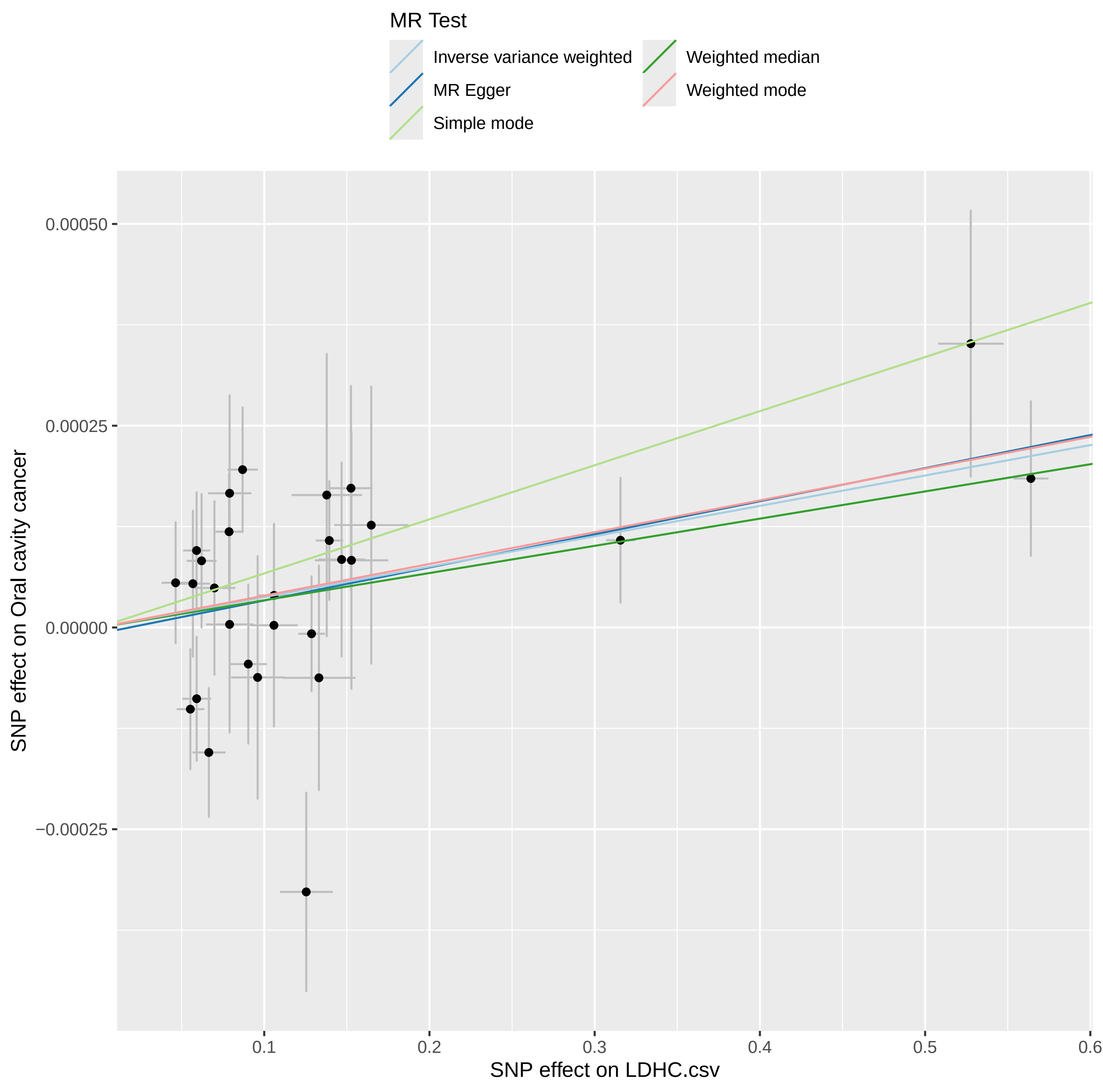

### scatter.pdf

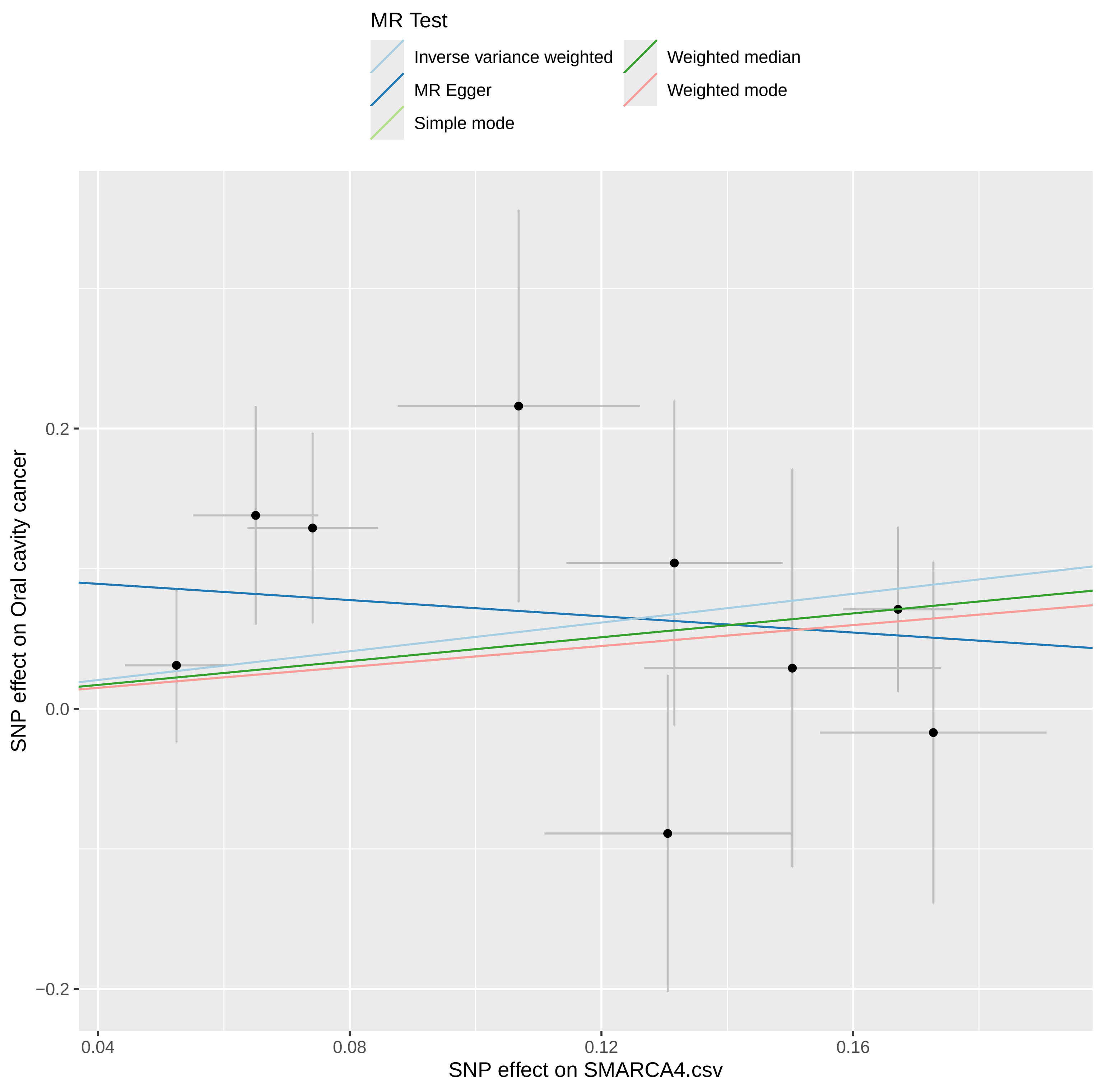

### scatter.pdf

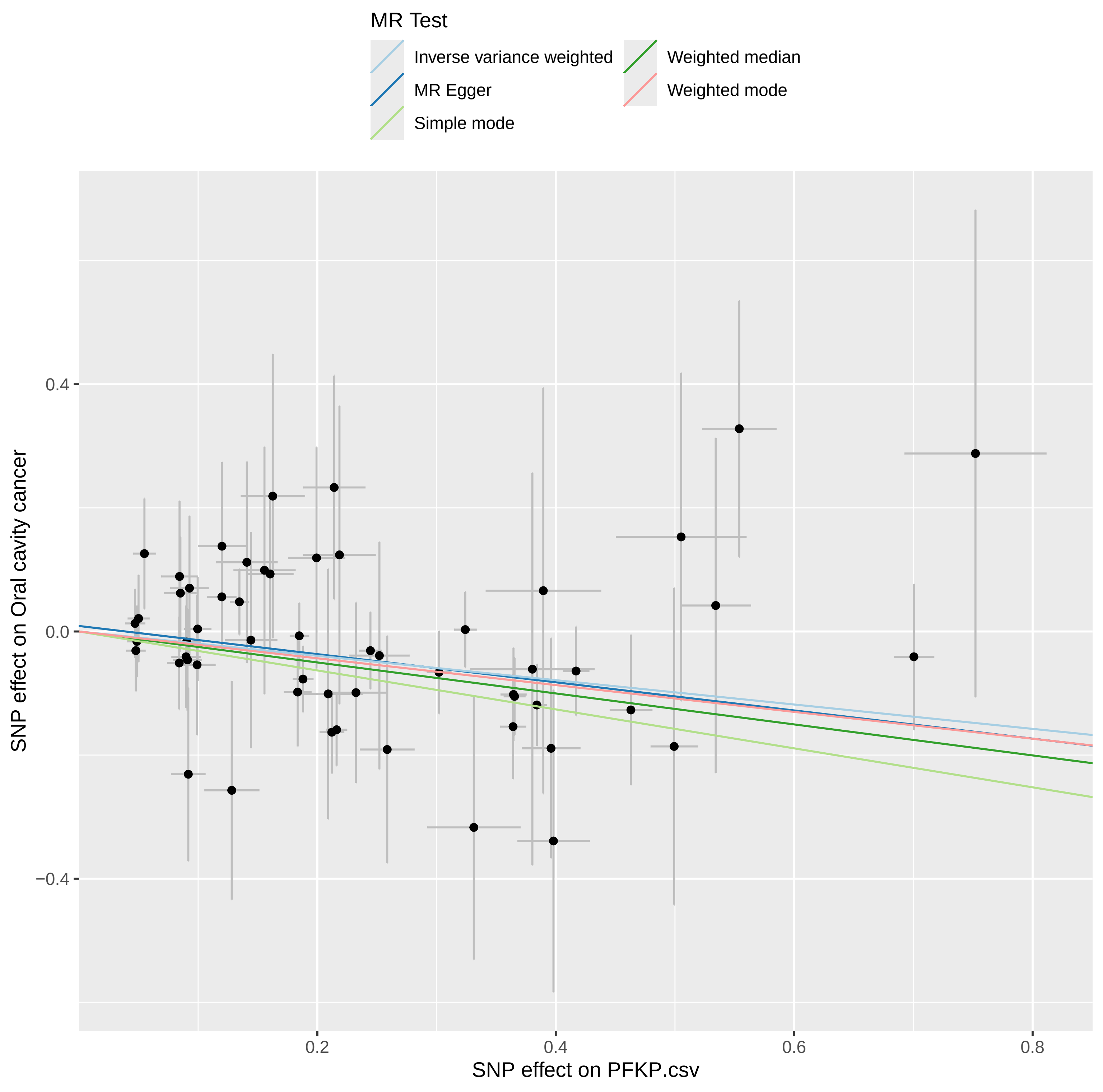

### scatter.pdf

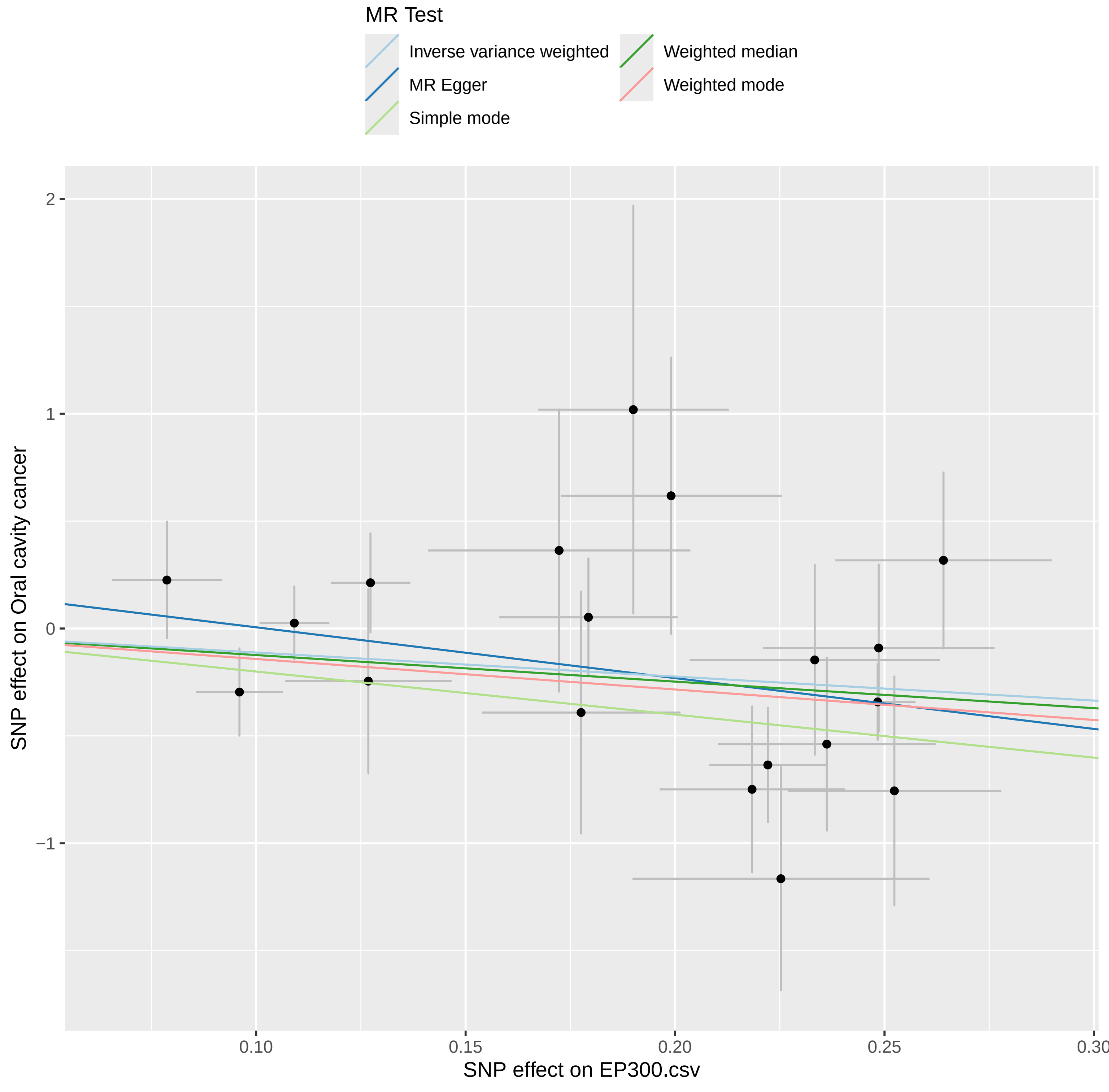

### scatter.pdf

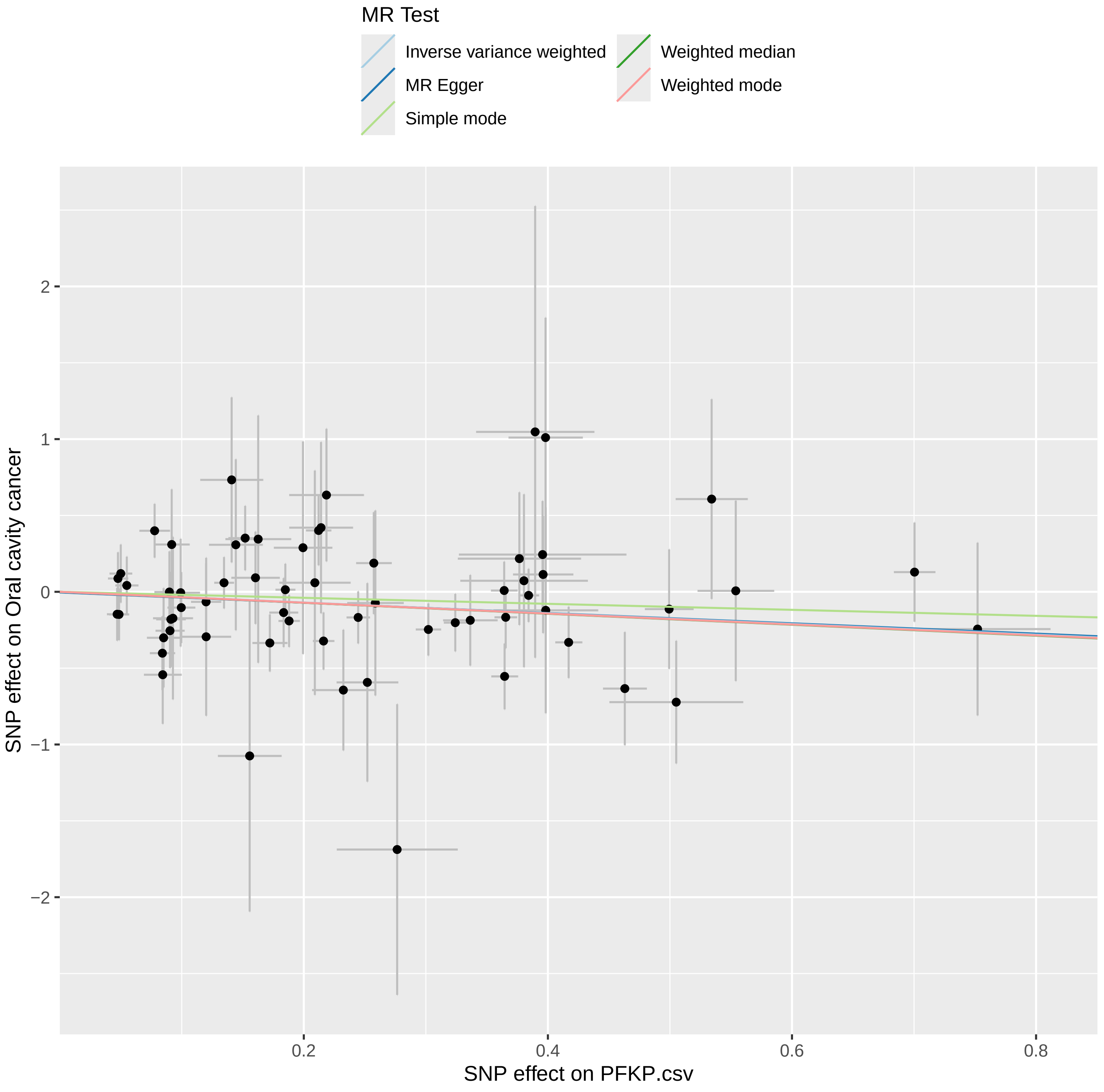

### sensitivity-analysis.pdf

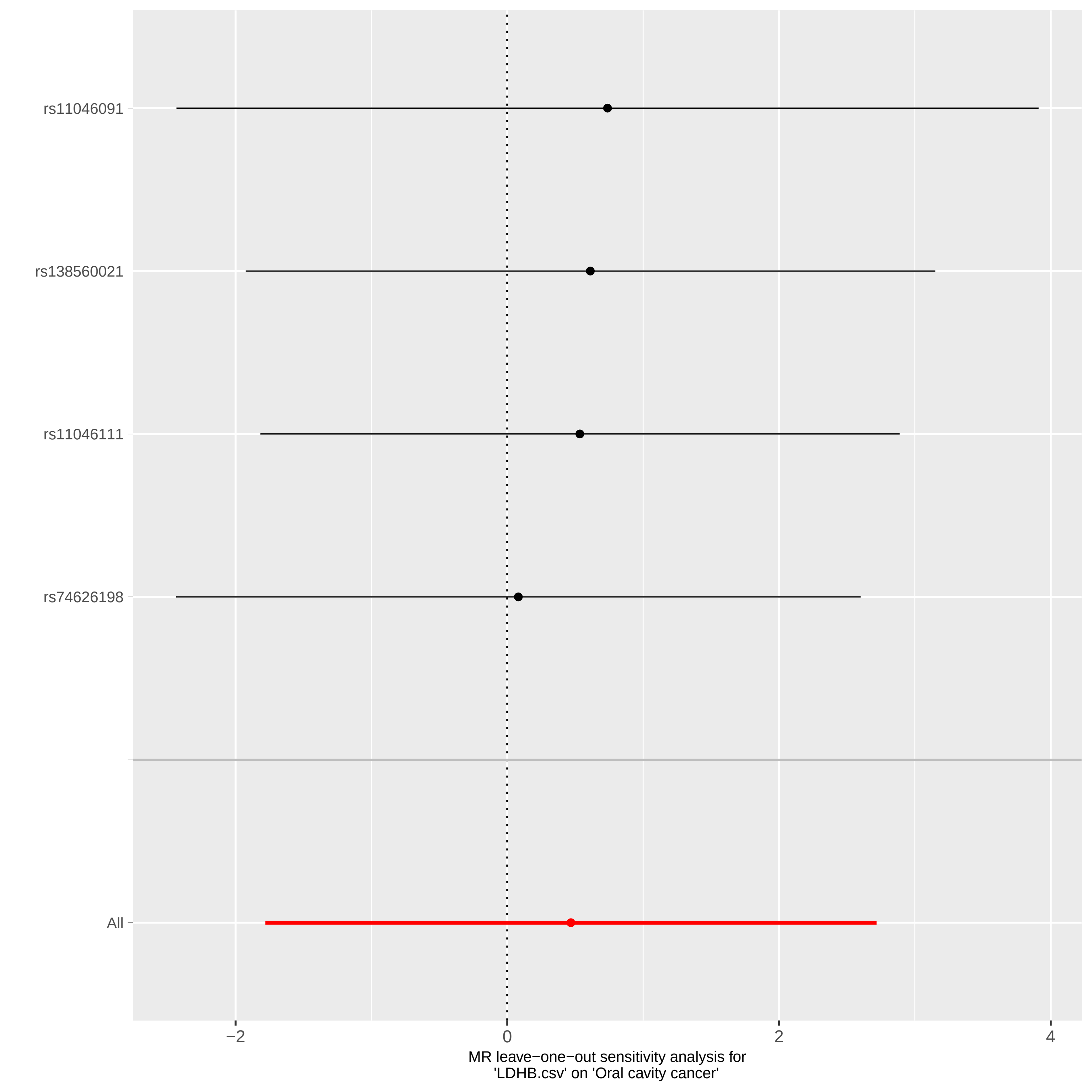

### sensitivity-analysis.pdf

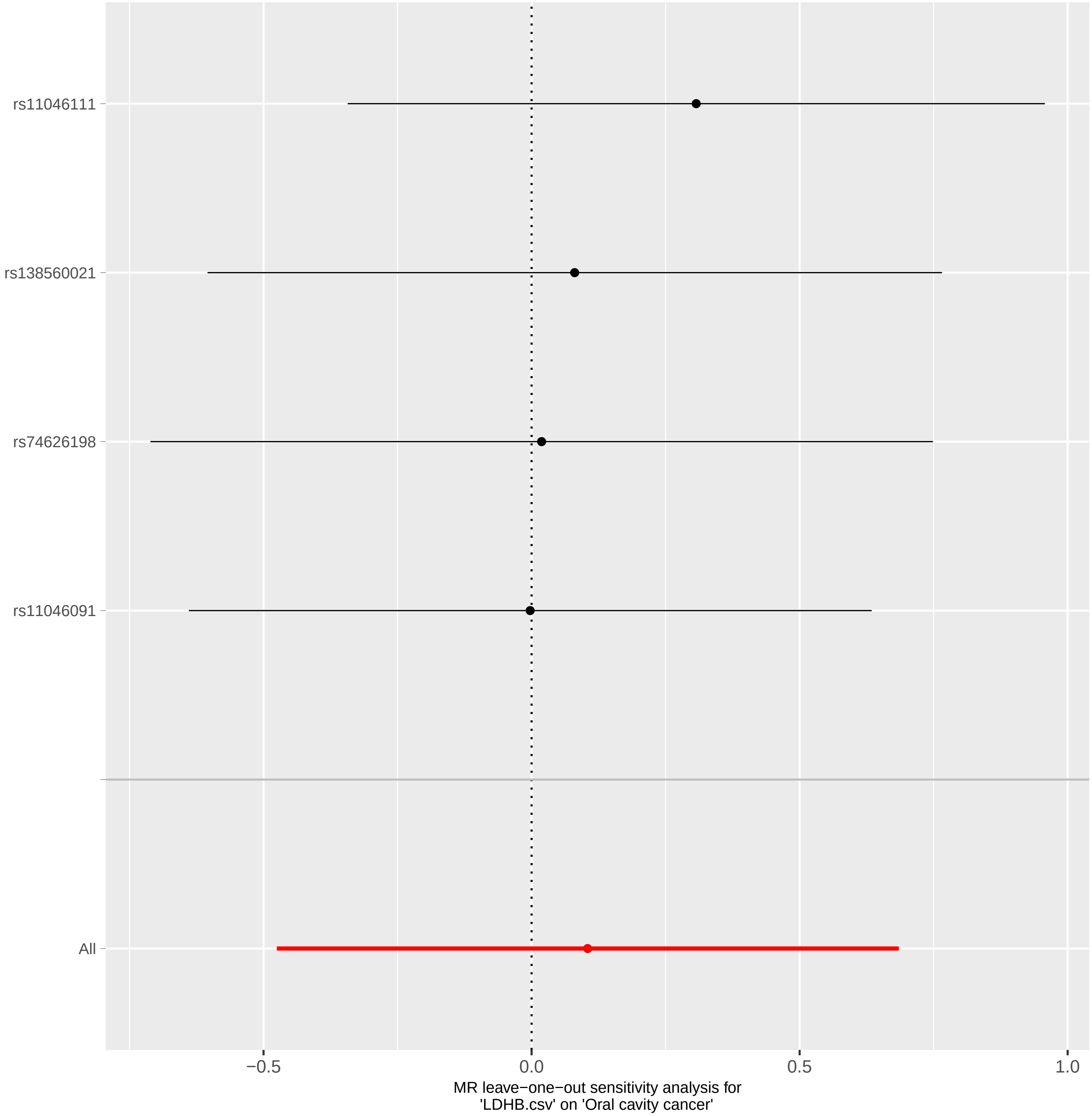

### sensitivity-analysis.pdf

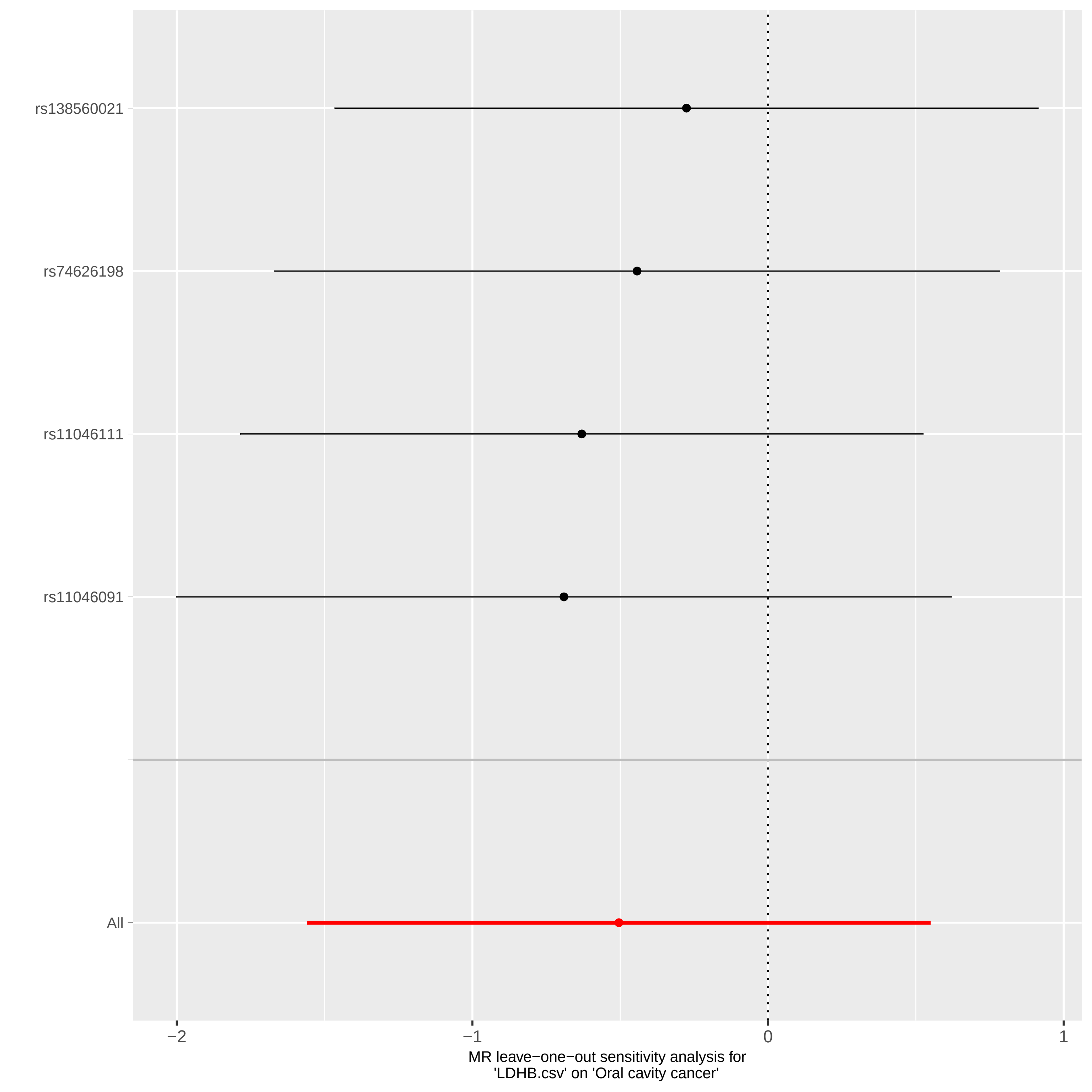

### sensitivity-analysis.pdf

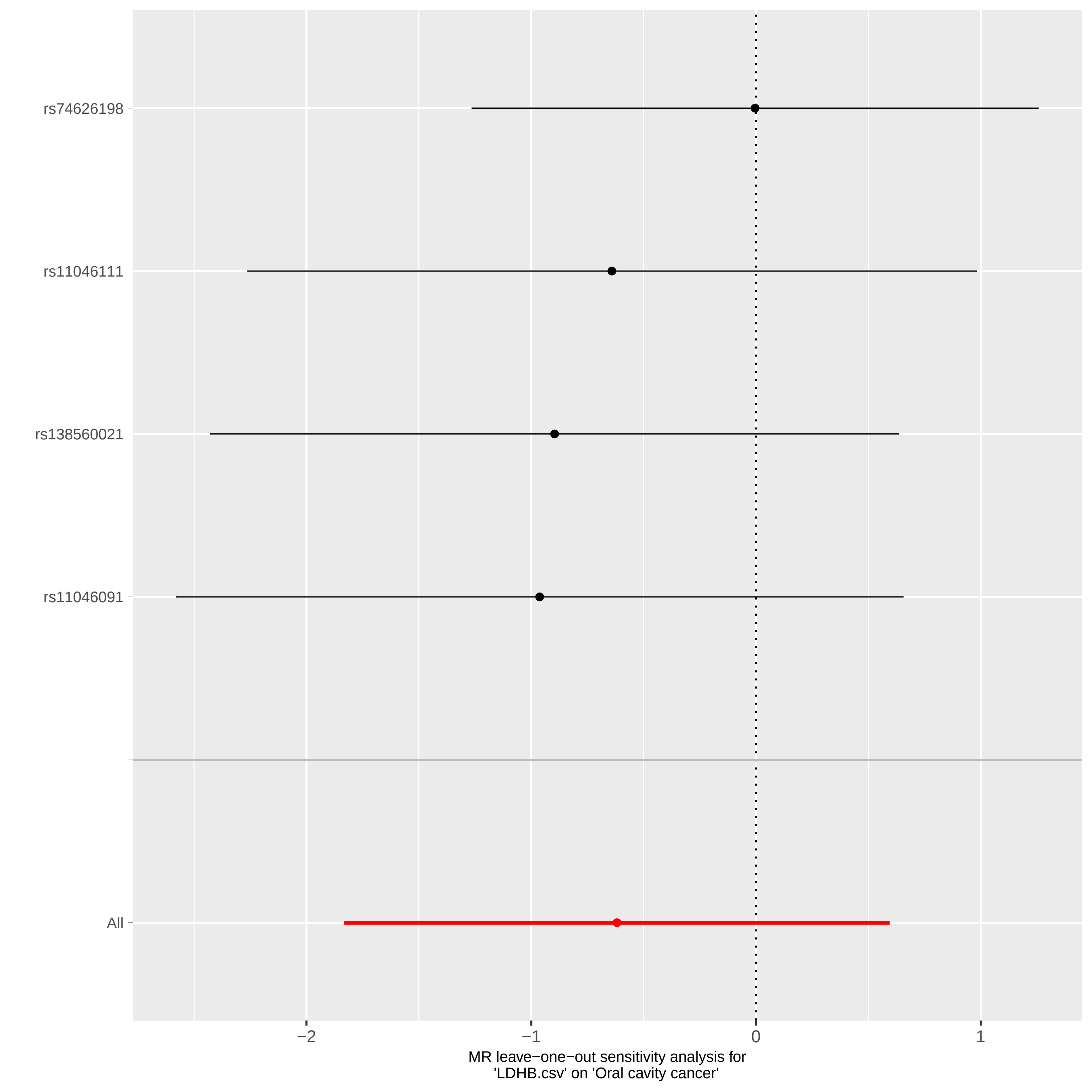

### sensitivity-analysis.pdf

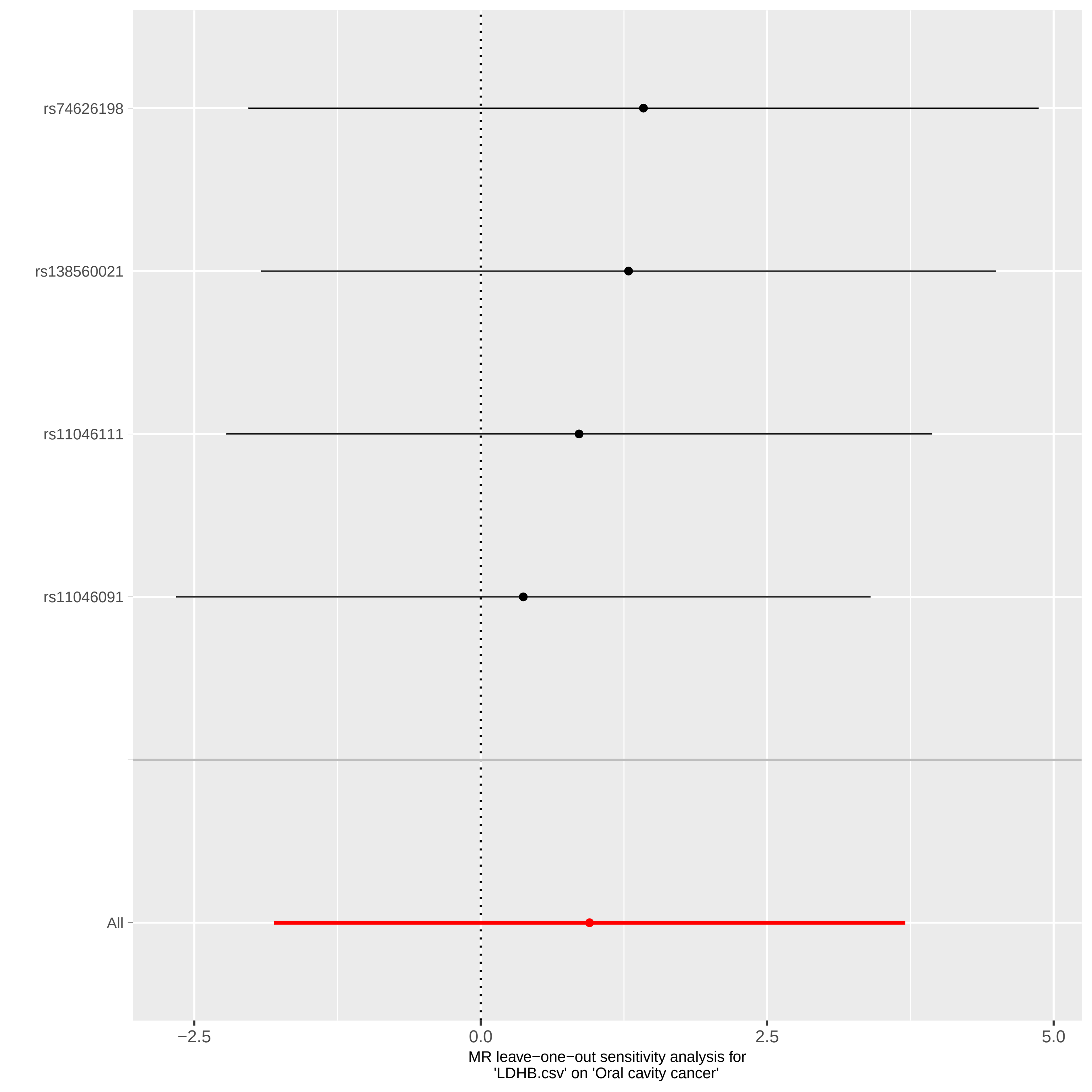

### sensitivity-analysis.pdf

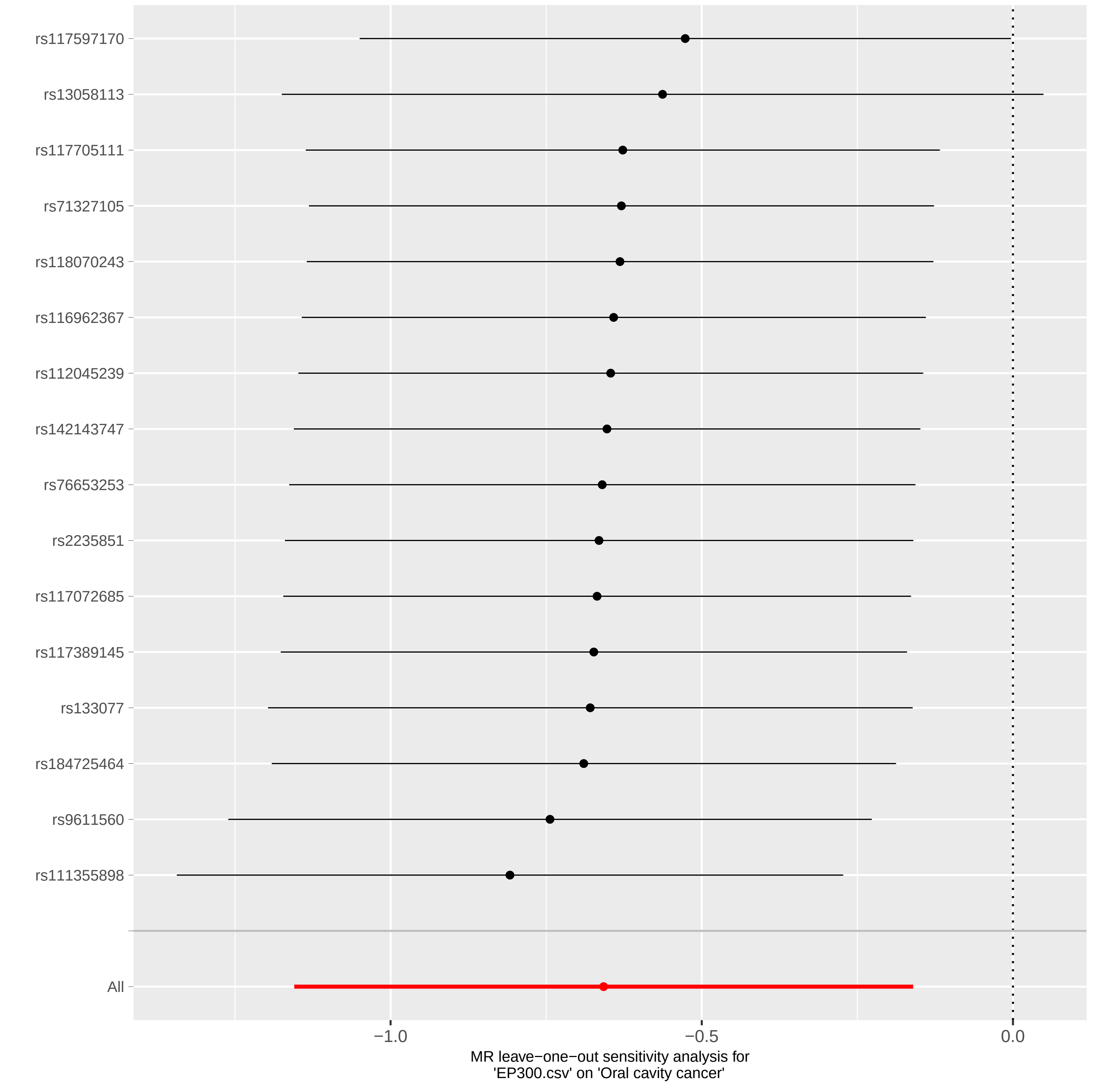

### sensitivity-analysis.pdf

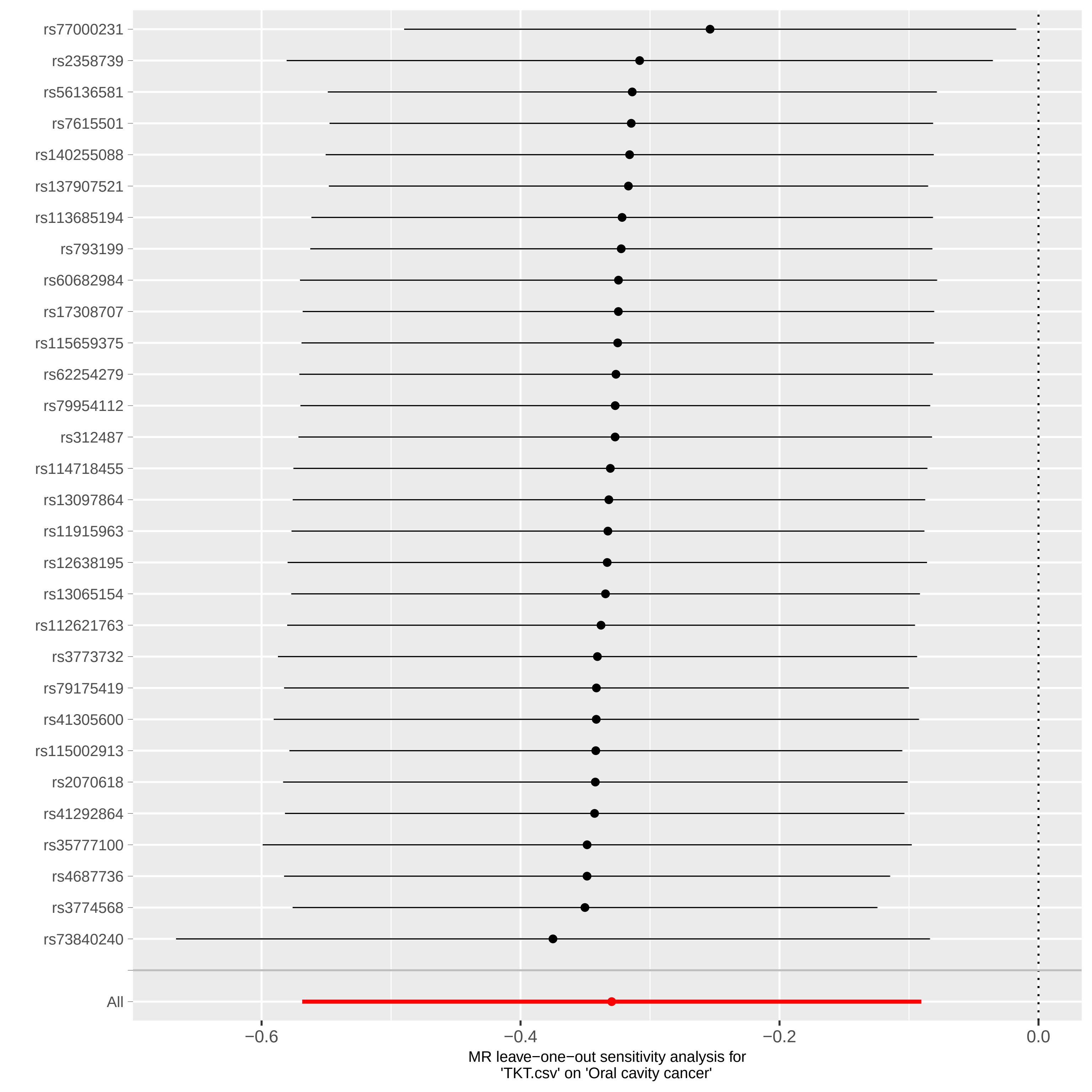

### sensitivity-analysis.pdf

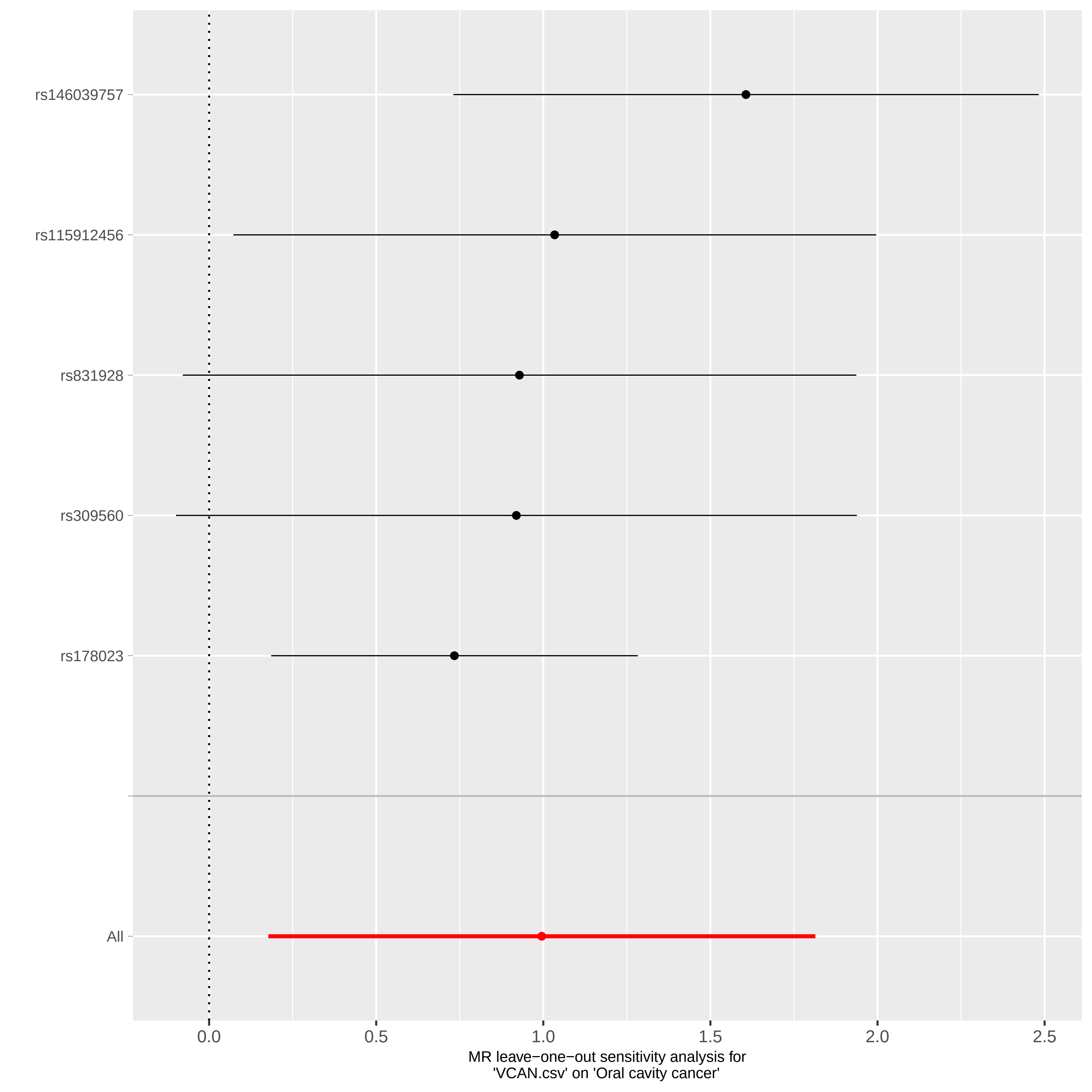

### sensitivity-analysis.pdf

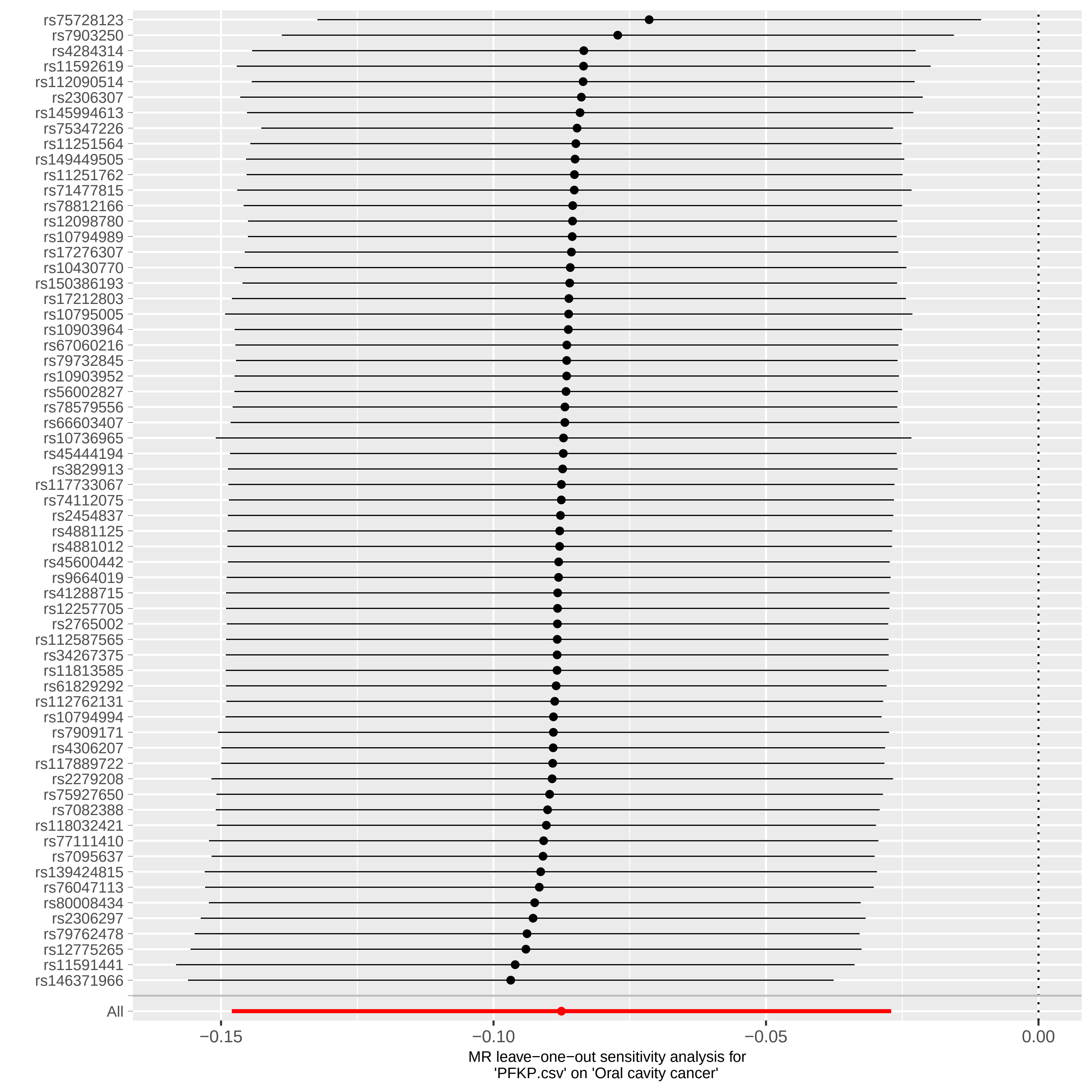

### sensitivity-analysis.pdf

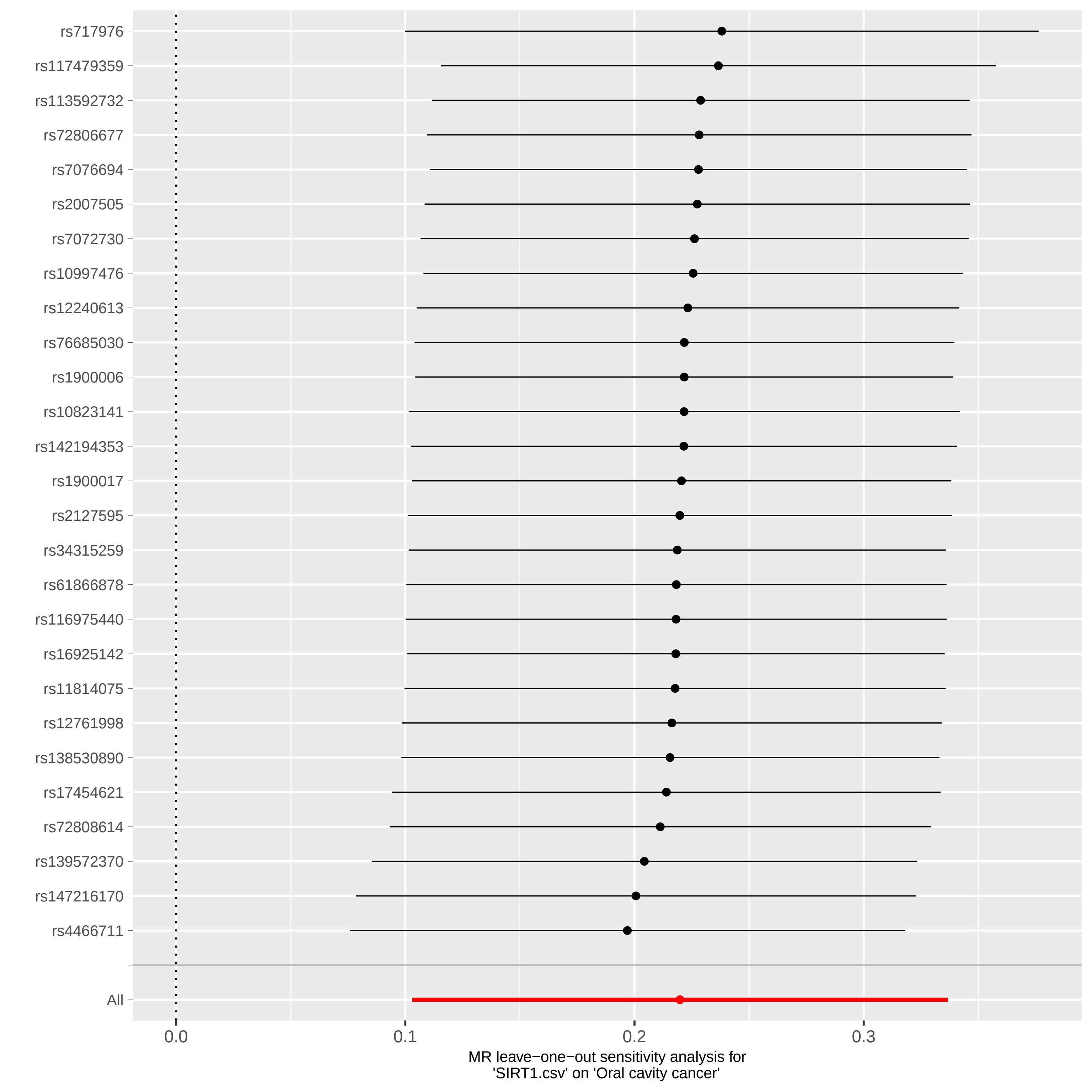

### sensitivity-analysis.pdf

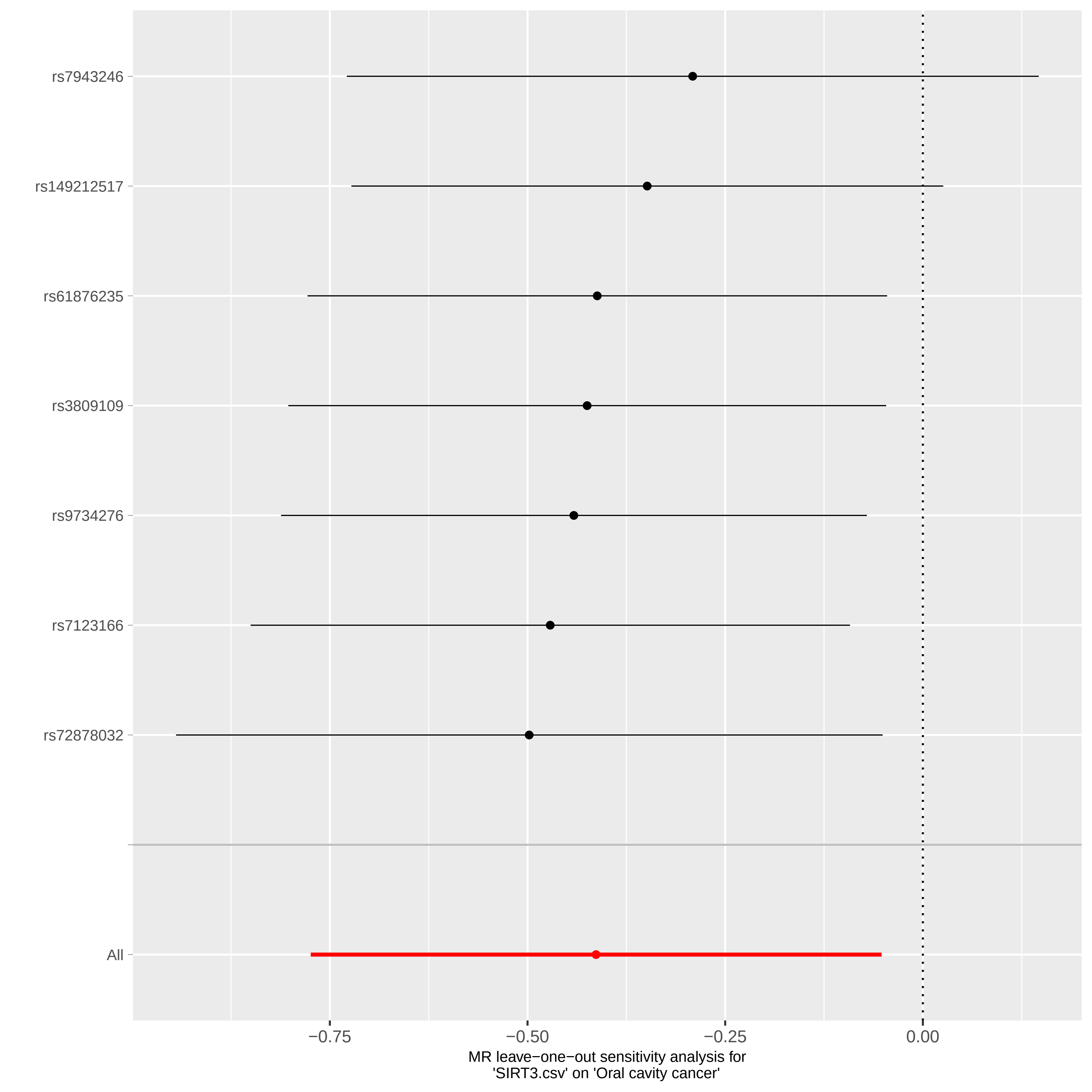

### sensitivity-analysis.pdf

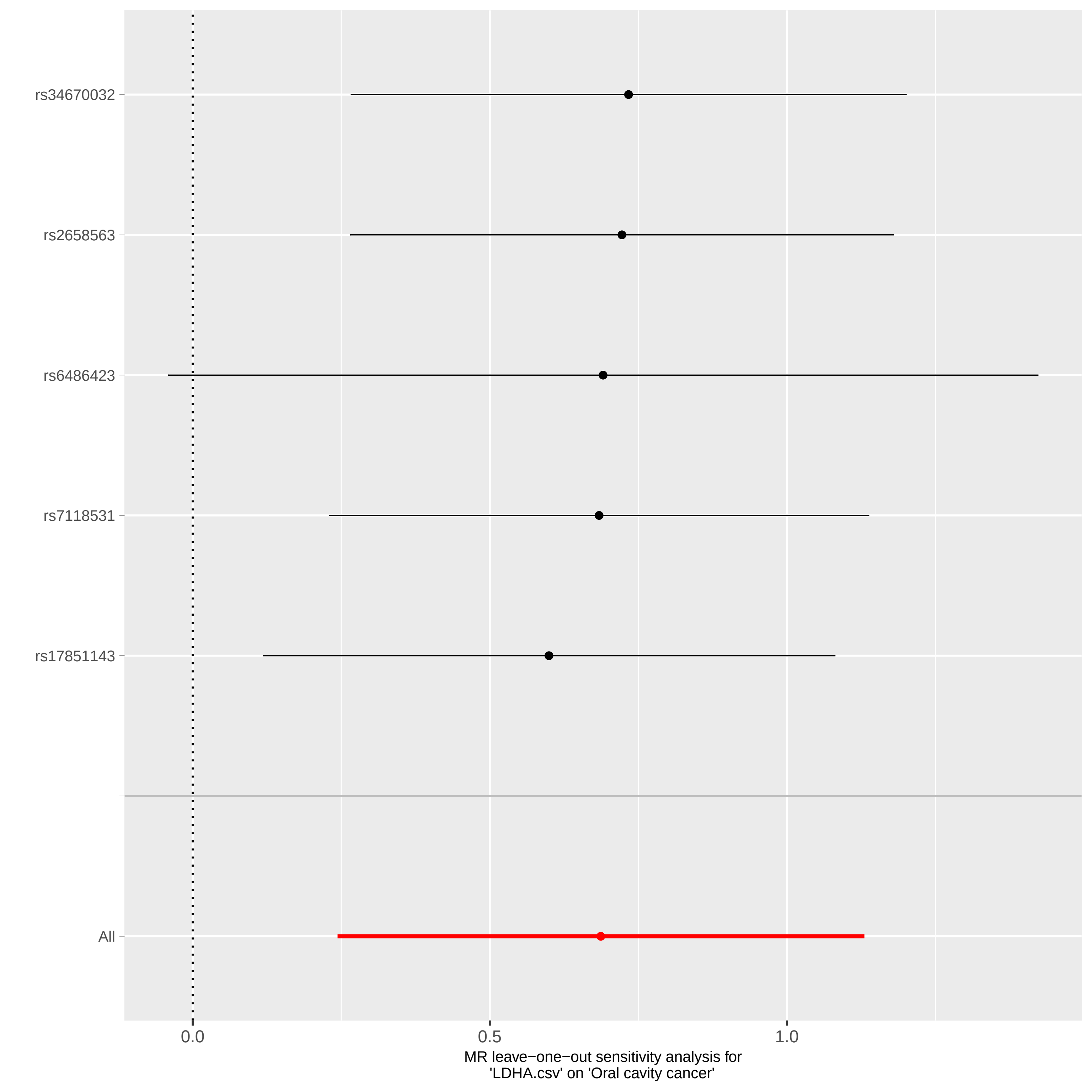

### sensitivity-analysis.pdf

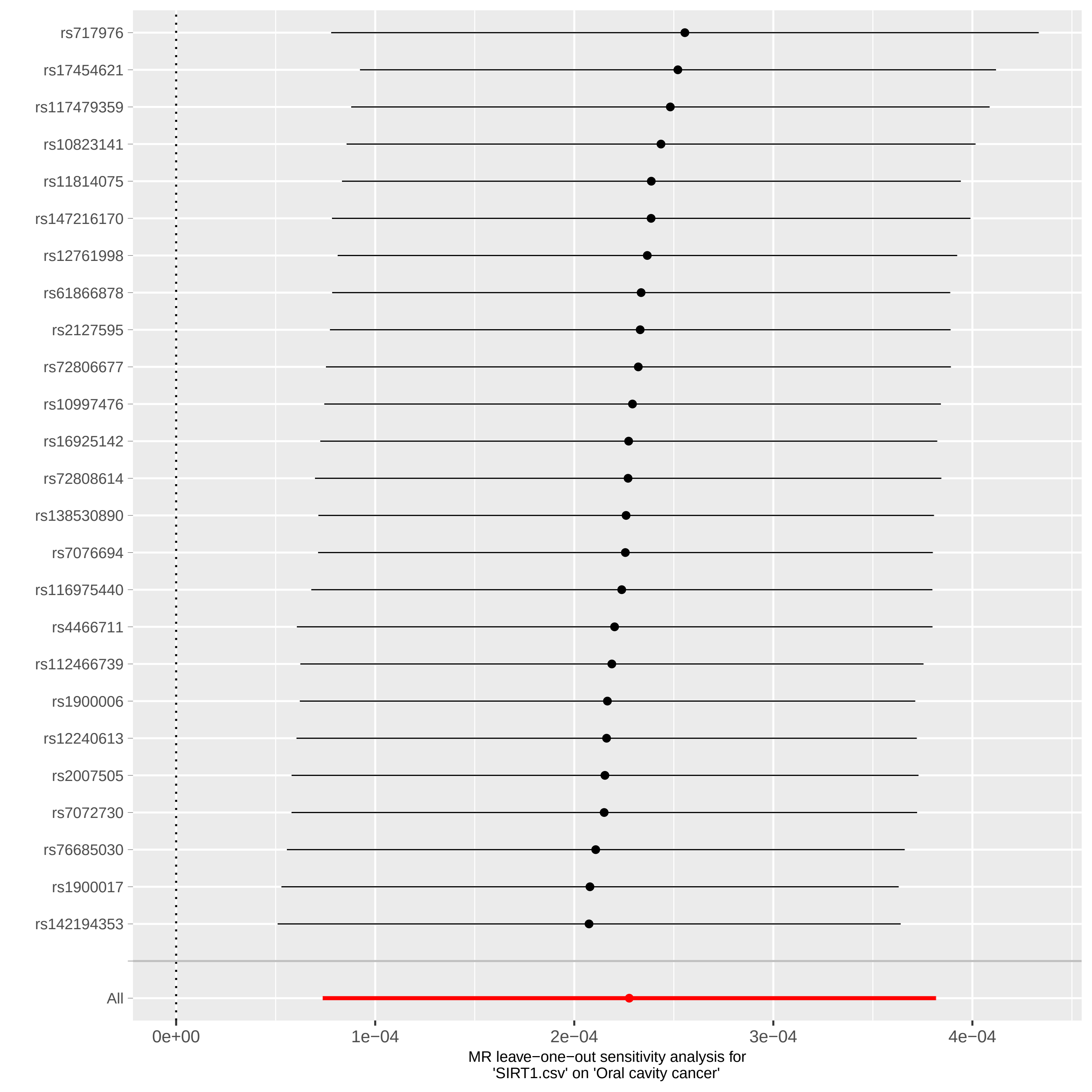

### sensitivity-analysis.pdf

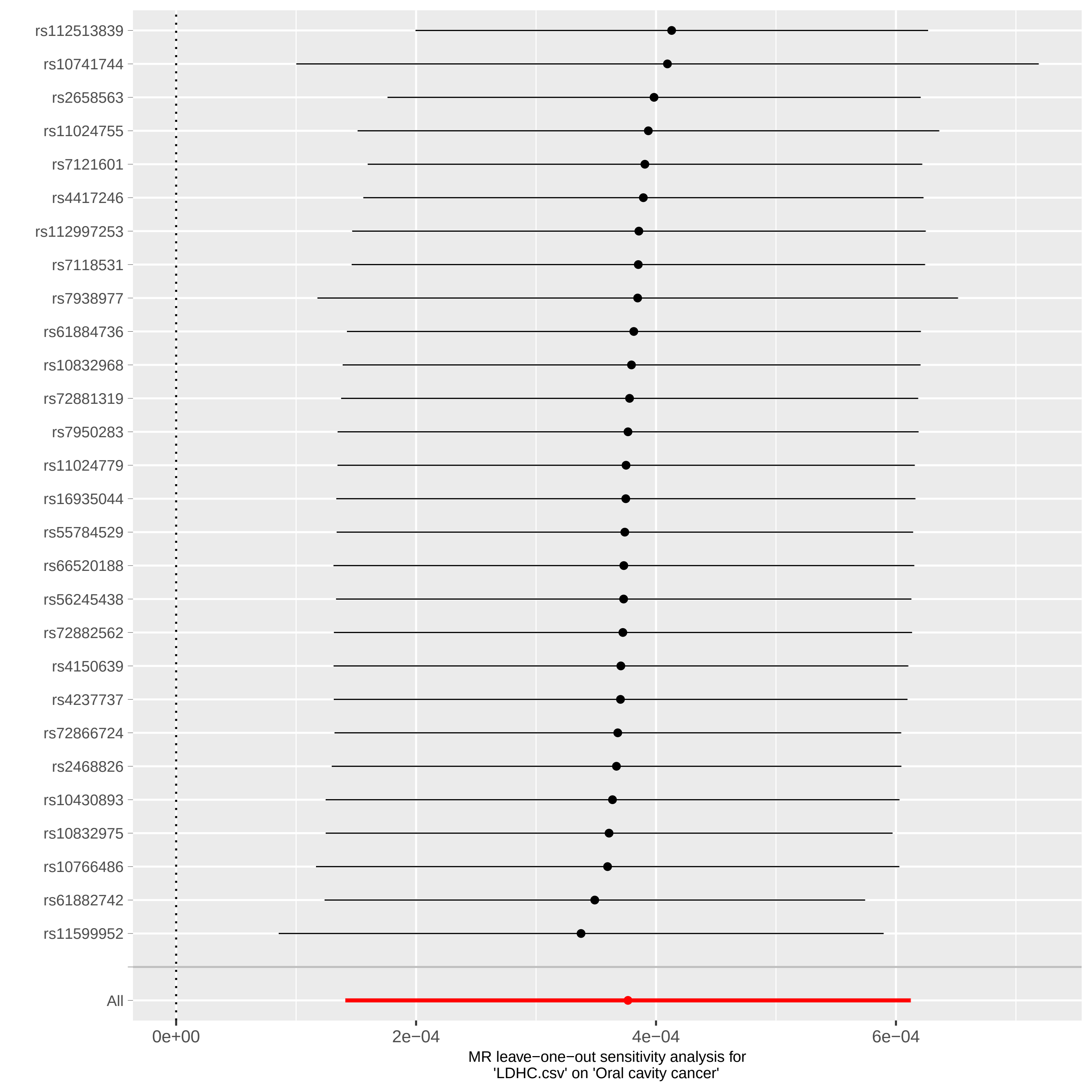

### sensitivity-analysis.pdf

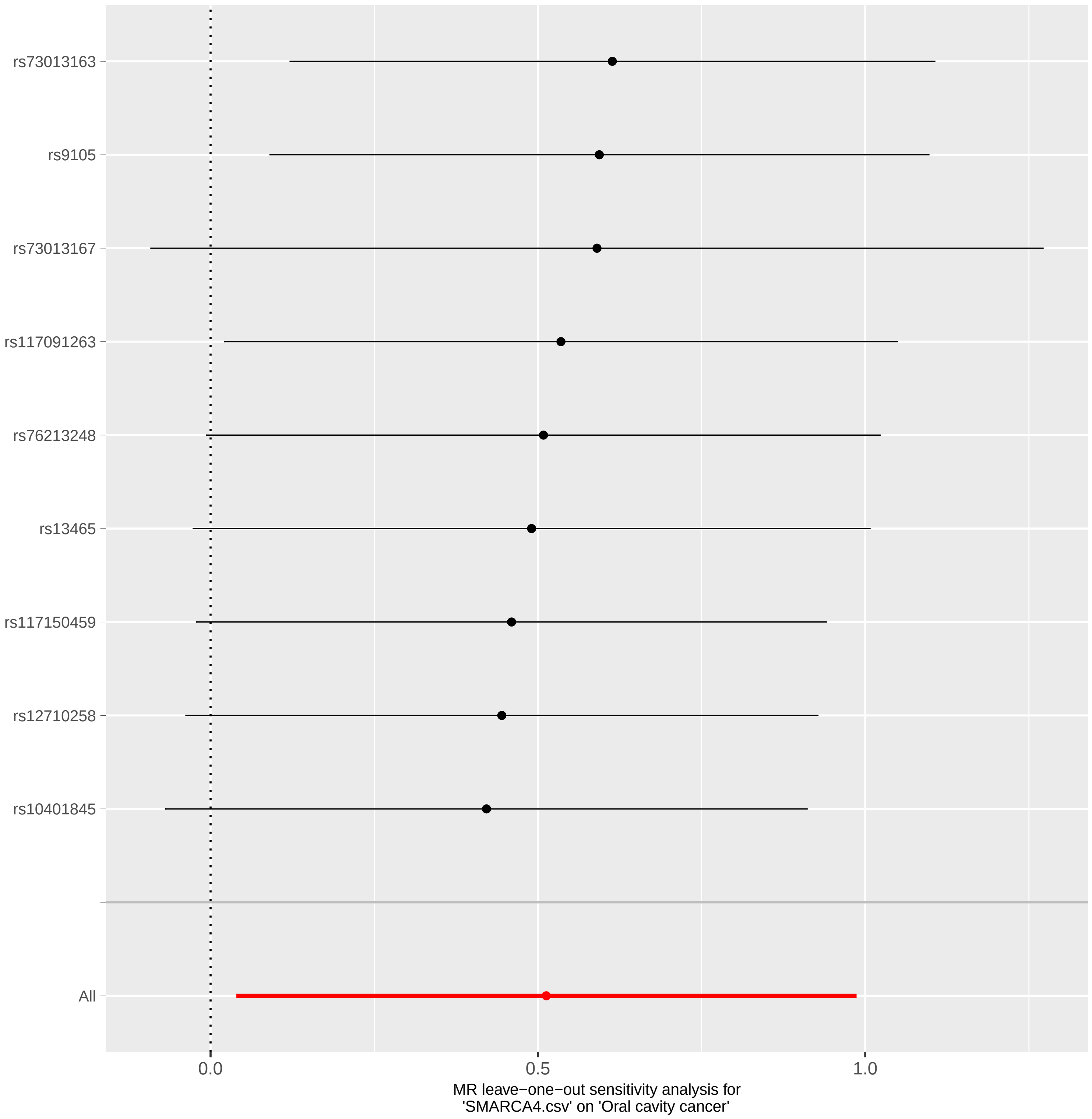

### sensitivity-analysis.pdf

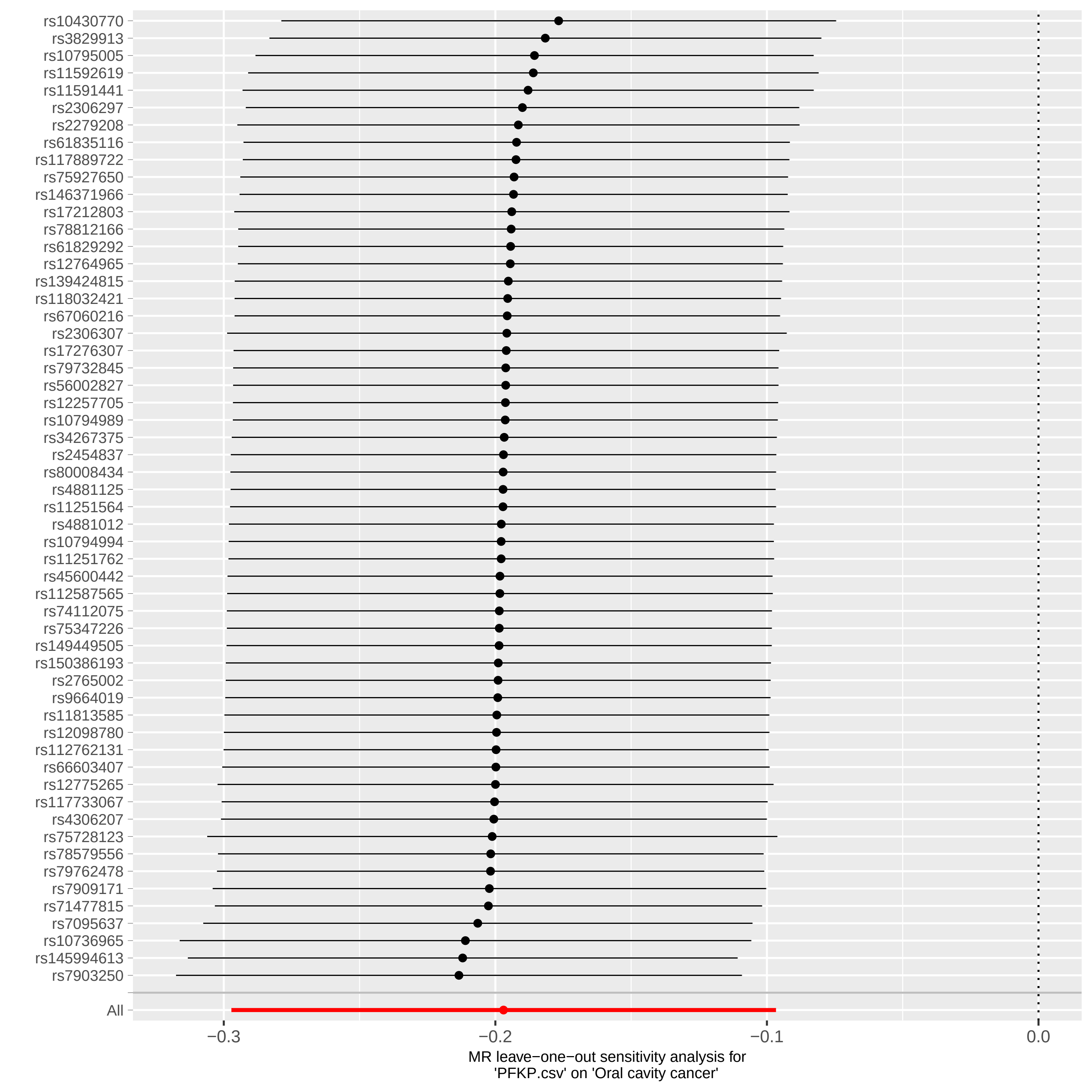

### sensitivity-analysis.pdf

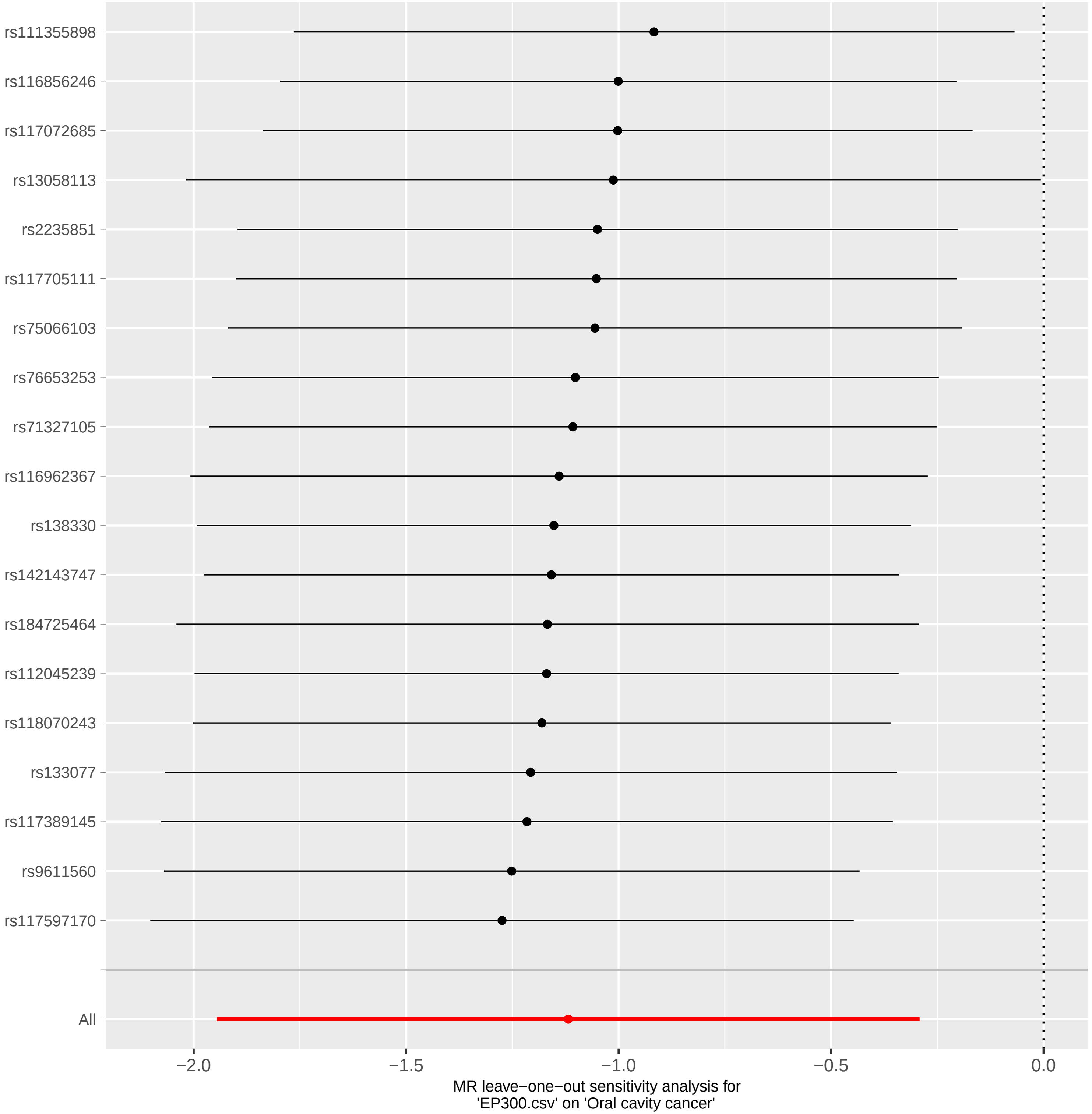

### sensitivity-analysis.pdf

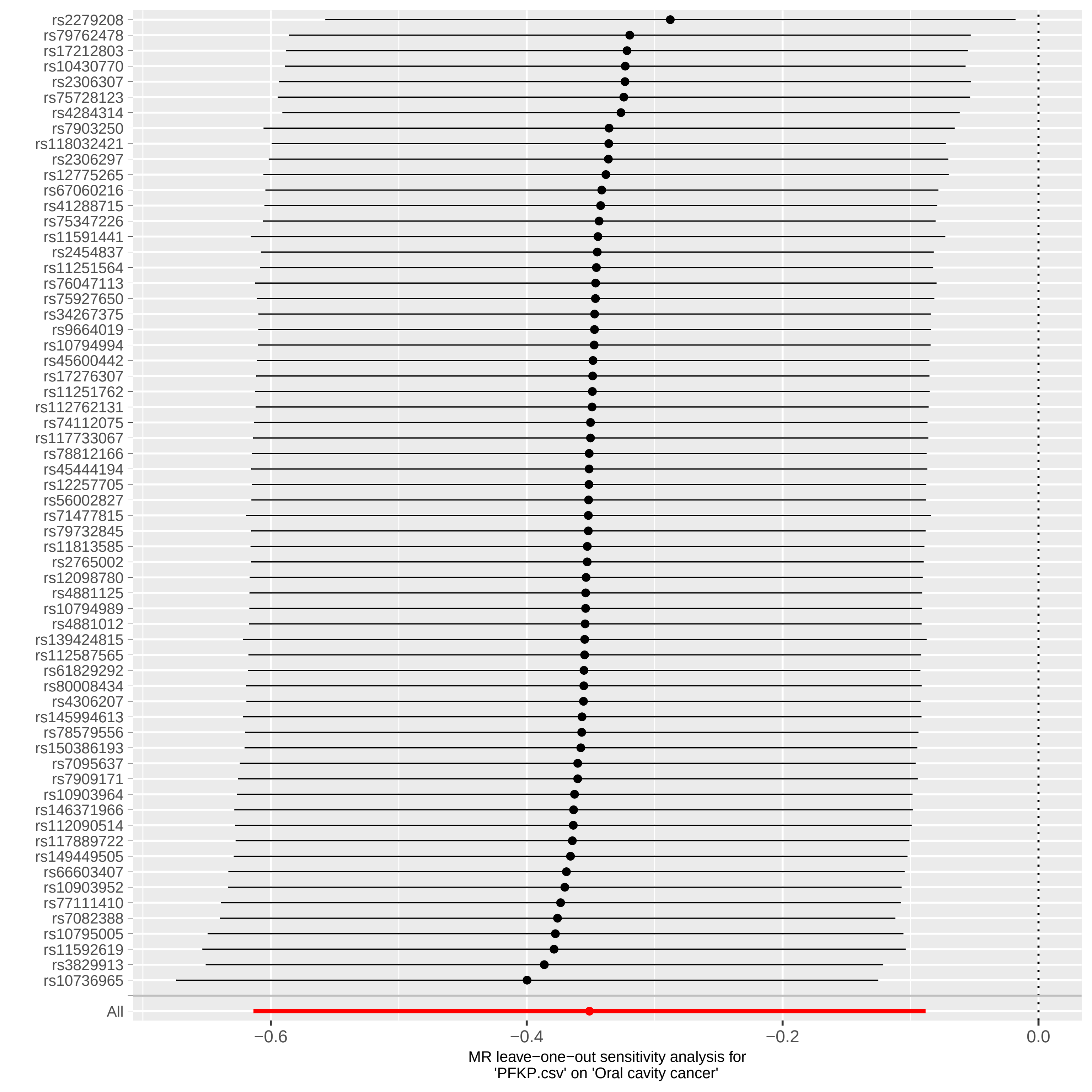
